# Association of clinical outcome assessments of mobility capacity and incident disability in community-dwelling older adults - a systematic review and meta-analysis

**DOI:** 10.1101/2022.03.02.22271795

**Authors:** Tobias Braun, Christian Thiel, Raphael Simon Peter, Carolin Bahns, Gisela Büchele, Kilian Rapp, Clemens Becker, Christian Grüneberg

## Abstract

**IMPORTANCE:** The predictive value of common performance-based outcome assessments of mobility capacity on incident disability in activities of daily living in community-dwelling older adults remains uncertain.

**OBJECTIVE:** To synthesize all available research on the association between mobility capacity and incident disability in non-disabled older adults.

**DATA SOURCES:** MEDLINE, EMBASE and CINAHL databases were searched without any limits or restrictions.

**STUDY SELECTION:** Published reports of longitudinal cohort studies that estimated a direct association between baseline mobility capacity, assessed with a standardized outcome assessment, and subsequent development of disability, including initially non-disabled older adults.

**DATA EXTRACTION AND SYNTHESIS:** Data extraction was completed by independent pairs of reviewers. The risk of bias was assessed using the Quality in Prognosis Studies (QUIPS) tool. Random-effect models were used to explore the objective. The certainty of evidence was assessed using GRADE.

**MAIN OUTCOME AND MEASURES:** The main outcome measures were the pooled relative risks (RR) per one conventional unit per mobility assessment for incident disability in activities of daily living.

**RESULTS:** A total of 40 reports were included, evaluating 85,515 and 78,379 participants at baseline and follow-up, respectively (median mean age: 74.6 years). The median disability rate at follow-up was 12.0% (IQR: 5.4%–23.3%). The overall risk of bias was judged as low, moderate and high in 6 (15%), 6 (15%), and 28 (70%) reports, respectively.

For usual and fast gait speed, the RR per -0.1 m/s was 1.23 (95% CI: 1.18–1.28; 26,638 participants) and 1.28 (95% CI: 1.19–1.38; 8,161 participants), respectively. Each point decrease in Short Physical Performance Battery score increased the risk of incident disability by 30% (RR = 1.30, 95% CI: 1.23– 1.38; 9,183 participants). The RR of incident disability by each second increase in Timed Up and Go test and Chair Rise Test performance was 1.15 (95% CI: 1.09–1.21; 30,426 participants) and 1.07 (95% CI: 1.04–1.10; 9,450 participants), respectively.

**CONCLUSIONS AND RELEVANCE:** Among community-dwelling non-disabled older adults, a poor mobility capacity is a potent modifiable risk factor for incident disability. Mobility impairment should be mandated as a quality indicator of health for older people.

**Key Points:** *QUESTION:* What are the associations between clinical outcome assessments of mobility capacity and incident disability in community-dwelling older adults?

*FINDINGS:* In this systematic review and meta-analysis that included 40 reports and data of 85,515 older adults, the risk ratios of incident disability on activities of daily living were 1.23, 1.30, 1.15, and 1.07 for usual gait speed, Short Physical Performance Battery, Timed Up and Go test, and Chair Rise Test, respectively, per one conventional unit.

*MEANING:* Common assessments of mobility capacity may help identify older people at risk of incident disability and should be routinely established in regular health examinations.

## Background

Activities of daily living (ADL), such as bathing, dressing, and toileting, are essential for older individuals to remain independent.^1^ Disability can be defined on the basis of having difficulty with one or more ADLs or needing (or receiving) help with a functional task, such as transferring, eating, and walking.^2^ ADL disability is associated with increased mortality, institutionalization, greater use of care, and a lower quality of life.^1,3^ Disability is a complex, multidimensional, and dynamic process that can be prevented, delayed, or improved through evidence-based interventions, which are most effective if they are based on personalized risk stratification.^4,5^

Mobility impairment, defined as reduced mobility capacity, is one of the strongest modifiable predictors for incident ADL disability in older adults.^6-9^ Mobility capacity can be used for stratification to start interventions.^10^ There are established methods to quantify mobility capacity such as single-component or multi-component clinical outcome assessments (COA) of walking, balance, transferring, or lower-extremity functioning, including comfortable or fast gait speed, chair rise performance, the Timed Up and Go test (TUG), or the Short Physical Performance Battery (SPPB).^10,11^ These performance-based measures of supervised capacity are relatively simple to perform and accepted by older people.^8,12^

There is solid evidence for a longitudinal association between physical functioning and disability onset and/or progression.^6,7,9,13,14^ In a recent meta-analysis, Wang et al.^7^ reported pooled longitudinal associations between physical performance measures (low/slow vs. high/fast) and ADL dependency for assessments of mobility, including the SPPB (odds ratio (OR): 3.5, 95% CI: 2.5–4.9), TUG (3.4, 1.9– 6.3), gait speed (2.3, 1.6–3.4), and Chair Rise Test (CRT; 1.9, 1.6–2.2). However, these authors did not include all relevant studies into the meta-analyses and they calculated pooled OR, which do not approximate risk ratios (RR) when the outcome is frequent and are sometimes misinterpreted.^15^ Thus, the predictive value of common COAs of mobility capacity on incident ADL disability in community-dwelling older adults remains uncertain. Therefore, the objective of this study was to systematically review the literature on the association between mobility capacity and incident ADL disability in community-dwelling older adults.

## Methods

The systematic review protocol was prospectively registered with PROSPERO (CRD42020160490). Analysis and reporting were informed by recent recommendations^16^ and performed according to the PRISMA 2020 statement.^17^

### Eligibility criteria

Reports were eligible if they met the following criteria: (1) Design: Published reports of longitudinal cohort studies that estimated a direct association between baseline mobility capacity and subsequent development of disability over a follow-up period of 1 to 6 years. (2) Population: Community-dwelling older adults, ≥ 60 years, non-disabled at baseline. (3) Prognostic factor: At least one COA of mobility capacity. (4) Outcome: At least one measure of ADL disability at follow-up, including nursing home admission, use of home care services or care dependency.

We excluded reports (1) not written in English or German, (2) solely focusing on individuals with clinical disorders such as dementia, and (3) from institutionalized settings such as hospitals or nursing homes. A full description of eligibility criteria is given in the eText 1 (Supplement).

### Information sources and search strategy

We searched MEDLINE (OVID), EMBASE (OVID), and CINAHL (EBSCOhost) from inception to February 18, 2021. We used a combination of search terms related to “community-dwelling older adults”, “mobility capacity”, “disability” and “prediction”, informed by published search filters.^16,18-20^ The search strategy was kept broad, reviewed and revised by a senior information specialist, adjusted for each database, and is reported in the Supplement (eTables 1, 2, and 3). We imposed no limits or restrictions on any of the searches.

We also screened reference lists of a convenience sample of 16 related reviews (listed in eTable 4; 1,048 references) from the authors’ private archives and manually screened reference lists of included reports for additional reports.

### Selection process and data extraction

References were stored in EndNote X7.7. After duplicate removal by EndNote, pairs of reviewers (TB, CBa, and a third person) independently screened the titles and abstracts of identified records to remove obviously irrelevant reports (then using an over-inclusive approach at this stage). The level of inter-rater agreement between two reviewers was calculated by means of a kappa statistic.

A full-text review of potentially relevant reports was conducted by one reviewer (TB) and validated by a second reviewer (CBa). If necessary, missing information was requested from the authors. The reasons for full-text exclusion were recorded. At all steps, disagreement between reviewers was resolved by discussion. The methods of data extraction are reported in the eText 2 (Supplement).

### Quality assessment

The risk of bias (RoB) of individual reports was assessed, as recommended,^16^ with the Quality in Prognosis Studies (QUIPS; six domains).^21^ The overall RoB was judged as proposed by Grooten et al.^22^ If all domains were classified as having low RoB, or up to one moderate RoB, the overall RoB was low. If one or more domains were classified as having high RoB, or ≥ 3 moderate RoB, the overall RoB was high. All reports in between were classified as having moderate RoB. The QUIPS tool was completed by one reviewer (TB).

The methodological quality of included reports was assessed using the Newcastle-Ottawa scale (NOS) for cohort studies.^23^ A detailed description of how we applied the NOS in this review is given in the eText 3 in the Supplement. The NOS was asses by one reviewer (TB).

The Grading of Recommendations, Assessment, Development and Evaluations (GRADE) approach for prognosis studies was used to assess the overall certainty of evidence of the meta-analyses as high, moderate, low, or very low.^24^

### Data synthesis and analysis

The primary outcome was any effect measure (eg, hazard ratio (HR), OR, RR) of baseline mobility capacity and incident ADL disability at follow-up for each pair predictor variable and outcome. In the meta-analysis, all results were re-expressed as RRs per unit of the respective mobility outcome assessment (eg, 0.1 m/s for gait speed).

The decision which reports/analyses were eligible for grouped synthesis was based on the following criteria: (a) A minimum of 5 reports for a given standardized mobility assessment. Analyses based on single items of a comprehensive assessment, such as single items of the POMA, were not eligible. (b) In case of different follow-up periods of the same sample, we used the longer period. (c) If results were reported for the total sample and clustered subgroups (eg, by gender), we only used the total sample. (d) If unadjusted and adjusted associations were reported, we used the most fully adjusted estimations.

Associations for different units of measurement (eg, per 1 m/s vs. per 0.1 m/s, walking time over 4 m vs. 6 m vs. 20 feet) were converted to one conventional unit per mobility assessment method (eg, 0.1 m/s for gait speed). Associations reported for ordinal categorical exposures (eg, for thirds, quarters, or by established cut-points) were transferred into continuous measures of association using methods of dose-response meta-analysis.^25^ ORs were converted into RRs using the marginal risk for the outcome reported in the respective study. Finally, the standard error of estimates (if not reported) was calculated from reported confidence intervals or p-values. Combined estimates from random effects meta-analyses were calculated using the inverse variance method and the DerSimonian-Laird estimator for τ^2^ and visualized as forest plots for each mobility assessment method. Statistical analyses were conducted using R (R Foundation for Statistical Computing, Vienna, Austria) version 3.6.3 with the packages ‘dosresmeta’ version 2.0.1 and ‘meta’ version 4.18-2. SPSS version 23.0 (IBM corp., Armonk, NY, USA) was used for descriptive data analysis. We conducted sensitivity meta-analyses restricted to trials with an overall low RoB.

To assess the RoB across studies (small-study effects), we generated funnel plots for meta-analyses including data of at least 10 reports of varying size and used Egger’s test for funnel plot asymmetry.^26^ In case of asymmetry, we reviewed the characteristics of the reports to check for possible reasons, such as publication bias.^26^

## Results

### Study selection

The database search identified 12,038 records (flow chart in eFigure 1 in the Supplement). After removal of duplicates, two pairs of reviewers screened the titles and abstracts of 9,187 records, with a kappa agreement of 0.56 (95% CI: 0.50–0.62, n = 8,777, TB and CBa) and 0.52 (95% CI: 0.37–0.67, n = 410, TB and LH), respectively. Full-text assessment of 318 articles led to the inclusion of 32 reports. The screening of the reference lists of 16 relevant reviews and the included reports led to the full-text assessment of 51 additional reports, of which 8 were included in this review. All 329 reports excluded after full-text assessment are listed in eTables 5 and 6 (Supplement) with reasons for exclusion.

**Figure 1.**
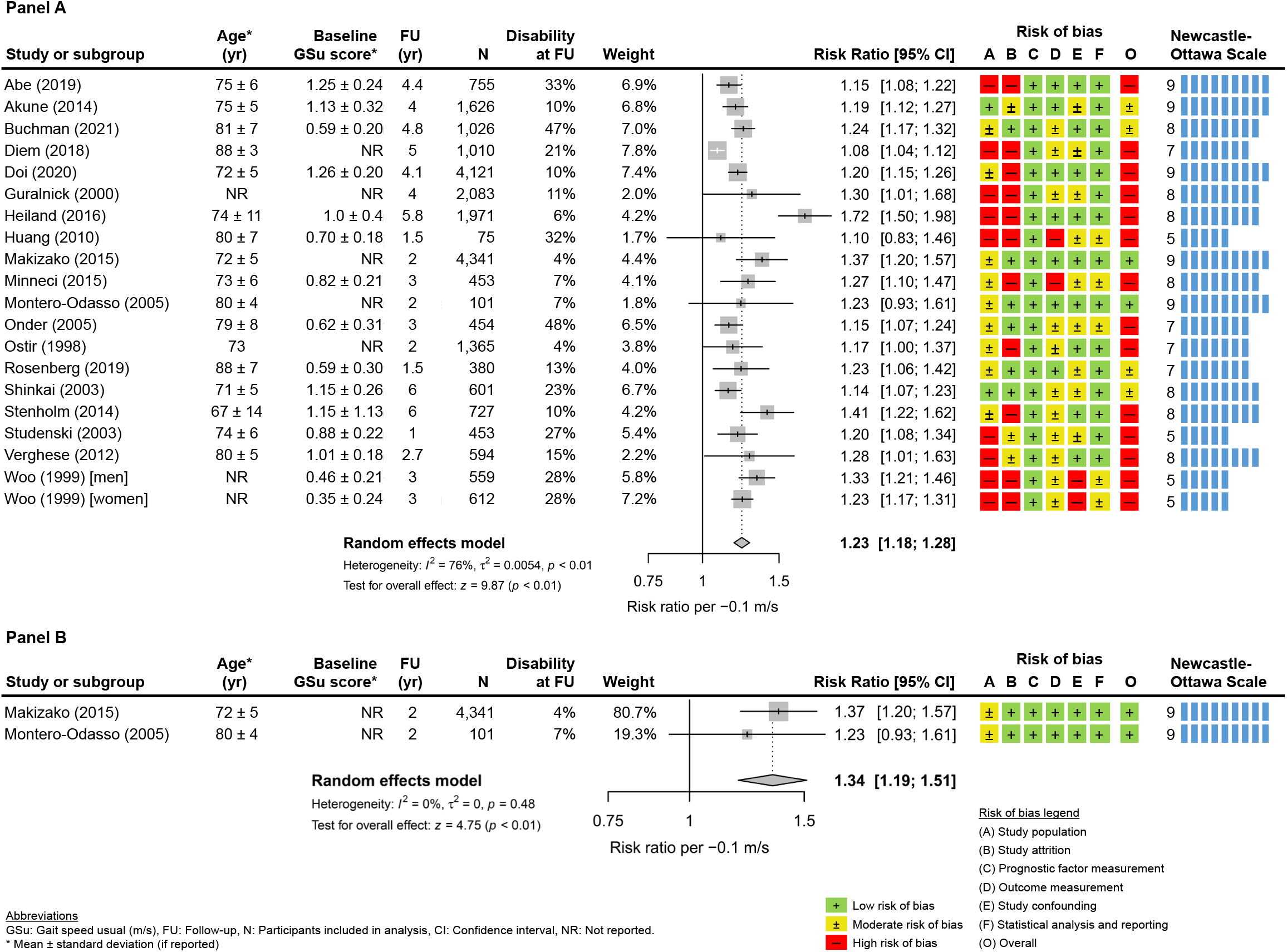
Association of usual gait speed with incident disability (panel A: all studies; panel B: only studies with an overall low risk of bias)

### Study characteristics

We included 40 reports based on 33 different longitudinal cohort studies (key and additional characteristics in eTable 7, eTable 8, and eTable 9 in the Supplement, respectively).^27-66^ Reports came from the US (15, 38%), Japan (10, 25%), Italy (4, 10%), or other single countries (11, 28%). Twenty-four (60%) reports were published between 2011 and 2021. The median follow-up was 3 years (IQR: 2–5; range: 1–6).

The total number of included participants at baseline and at follow-up were 85,515 (median of studies: 921; IQR: 465–1,929; range: 102–25,031) and 78,379 (median: 799; IQR: 453–1,655; range: 75–25,031), respectively. The median completion rate was 93% (IQR: 81%–100%; range: 56%–100%). Two reports included only women.^38,55^ All other reports included women and men, with a median proportion of women of 60% (IQR: 53%–70%; range 44%–100%). The minimal inclusion age of the single studies ranged from 60 years to 85 years, and the median mean age of participants was 74.6 years (IQR: 72.4–79.7; 37 reports).

Seventeen reports (43%) excluded older people with cognitive impairment, as indicated by a low MMSE score or the diagnosis of dementia. The median mean MMSE score at baseline was 26.9 points (IQR: 26.4–27.8; 20 reports).

The most frequent mobility assessment was usual gait speed (23 reports), followed by SPPB (12 reports), CRT (8 reports), fast gait speed (7 reports), and the TUG (7 reports). Balance tests (eg, one-leg standing, functional reach test) were each used in less than 5 reports. As expected, there was high heterogeneity in gait speed assessment conduction, i.e., with respect to distance, counting, or setting (eTable 10 in the Supplement).

Incident ADL disability according to (modified) Katz criteria was used as an outcome in 23 reports (58%), with a median of 15.7% of participants disabled at follow-up (IQR: 8.5%–27.5%). In those reports, ADL disability mainly was assessed by the self-reported inability or difficulty to perform ≥ 1 basic ADL, such as bathing, dressing, eating, toileting, transferring, or walking (overview of disability tasks in the eTable 11 in the Supplement). Incident long term care insurance (LTCI) certification was used in 9 reports (23%; all studies from Japan or Korea), with a median LTCI certification rate at follow-up of 9.6% (IQR: 3.8%–14.3%). Three reports (8%) recorded nursing home admissions/institutionalizations (cumulative event rates: 3.0%, 3.8%, and 16.6%). The median disability rate at follow-up, regardless of the disability outcome, was 12.0% (IQR: 5.4%–23.3%).

### Methodological quality and RoB in studies

The mean NOS score was 7.5 ± 1.6 (range: 3–9) points, with 33 reports (83%) rated with ≥ 7 points (rating per report in eTable 12 in the Supplement).

RoB was relatively low concerning the measurement of the prognostic factor and the statistical analysis and reporting. For the other four categories, RoB was rather moderate or high, especially concerning study population and study attrition. The overall RoB was judged as low, moderate, and high in 6 (15%), 6 (15%), and 28 (70%) reports, respectively (‘RoB summary figure’ eFigure 2 and ‘RoB graph’ in eFigure 3). The certainty of evidence for each prognostic factor is described the eTable 13.

**Figure 2:**
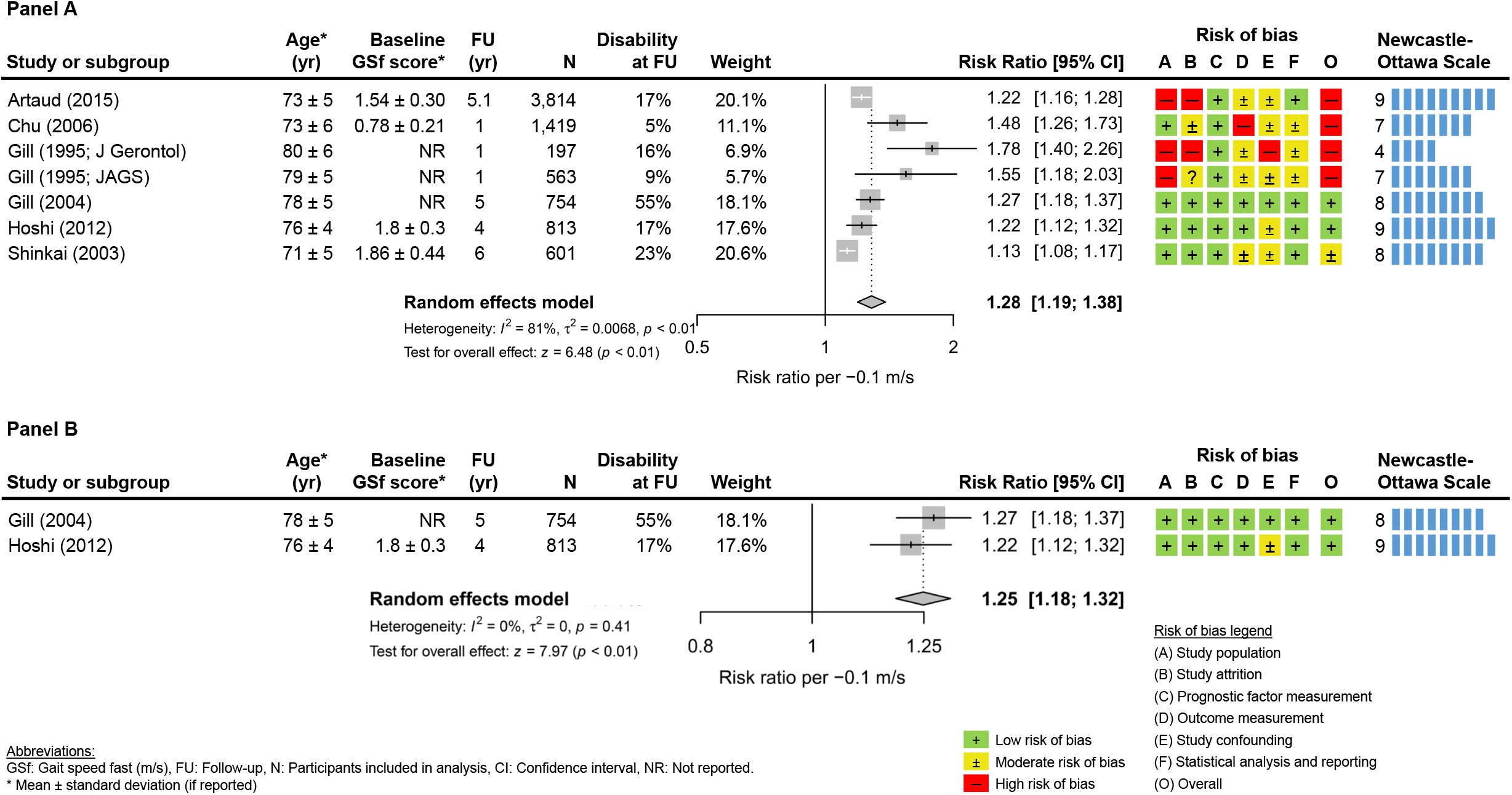
Association of fast gait speed with incident disability (panel A: all studies; panel B: only studies with an overall low risk of bias)

**Figure 3:**
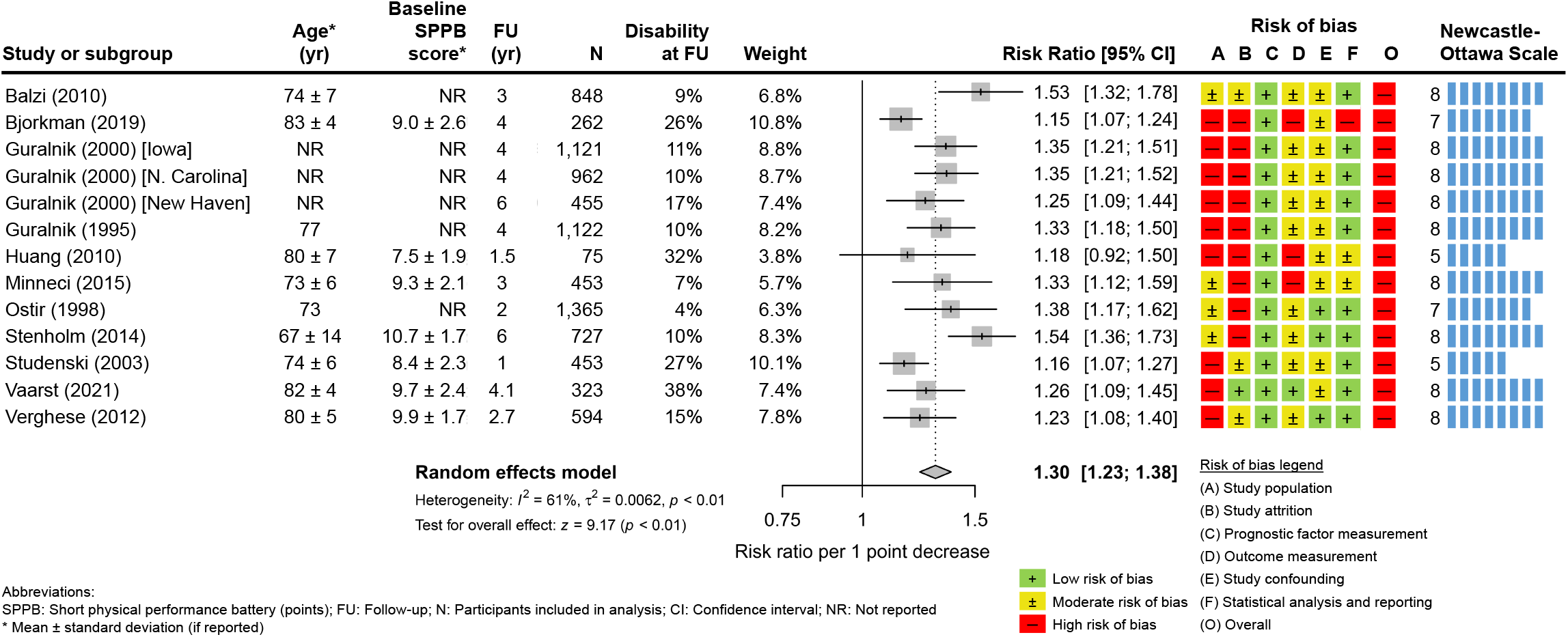
Association of Short Physical Performance Battery with incident disability

### Results of single studies

In total, the included reports presented 161 single effect estimations between baseline mobility capacity and ADL disability at follow-up (eTable 14 in the Supplement). Unless otherwise specified, all data were derived from the primary reference of each included report.

### Results of syntheses

#### Gait speed (usual)

Slower baseline gait speed at usual pace was significantly associated with disability at follow-up (RR per -0.1 m/s = 1.23, 95% CI: 1.18–1.28, *I*^2^ = 76%, 19 reports, 26,638 participants, Figure 1 panel A, moderate certainty of evidence).^27,28,33,38,39,45,46,48,50,52,53,55-57,59-61,64,65^ Egger’s test (t = 2.76, P = 0.01) indicated small-study effects for this analysis, which we attributed to true heterogeneity between reports after visual inspection of the funnel plot and a review of the study characteristics (eFigure 4 in the Supplement).

The association was even higher when only reports with an overall low RoB were included in the meta-analysis (RR per -0.1 m/s = 1.34, 95% CI: 1.19–1.51, I^2^ = 0%, 2 reports, 4,442 participants, Figure 1 panel B).^50,53^

#### Gait speed (fast)

For fast gait speed, the RR for incident disability at follow-up per -0,1 m/s was 1.28 (95% CI: 1.19– 1.38, I^2^ = 81%, 7 reports, 8,161 participants, Figure 2 panel A, low certainty of evidence).^29,35,41-43,47,59^ Based on reports with an overall low RoB, the RR did not change considerably (1.25, 95% CI: 1.18– 1.32, I^2^ = 0%, 2 reports, 1,567 participants, Figure 2 panel B).^43,47^

#### Short Physical Performance Battery

Each point decrease in SPPB score increased the risk of incident disability by 30% (RR per 1-point decrease = 1.30, 95% CI: 1.23–1.38, I^2^ = 61%, 11 reports, 9,183 participants, Figure 3, moderate certainty of evidence).^30,32,44,45,48,52,56,60,61,63,64^. All reports were judged to have an overall high RoB. There was no evidence of small-study effects (funnel plot in eFigure 5 in the Supplement; Egger test: t = 1.84, P = 0.09).

**Figure 4:**
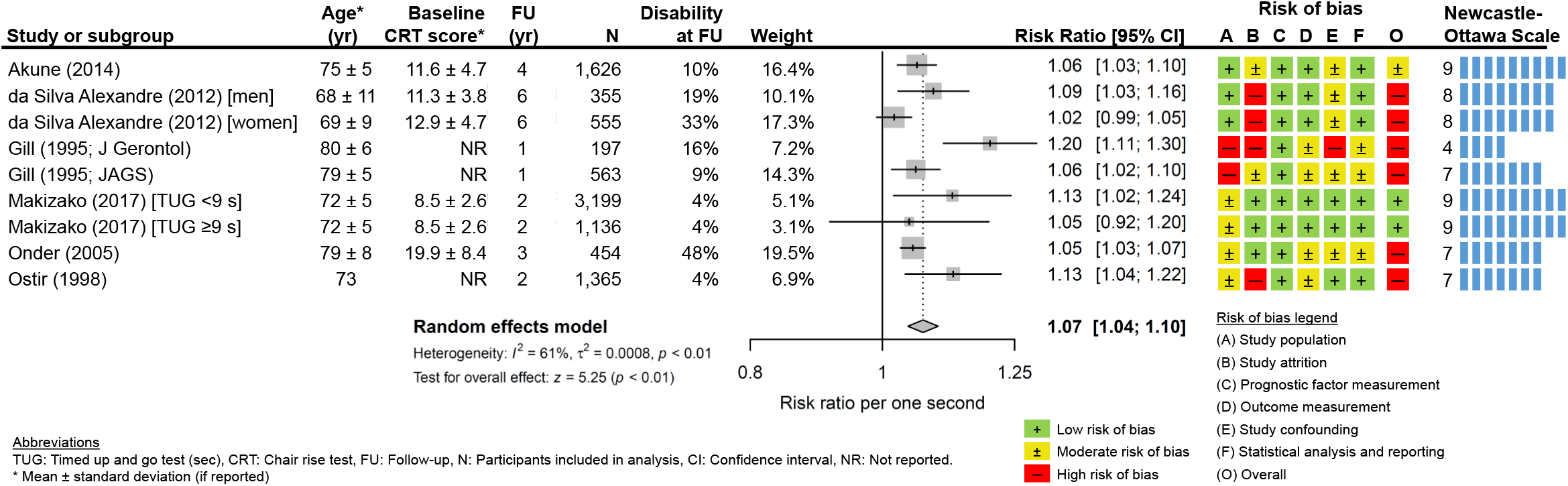
Association of Chair Rise Test with incident disability

**Figure 5:**
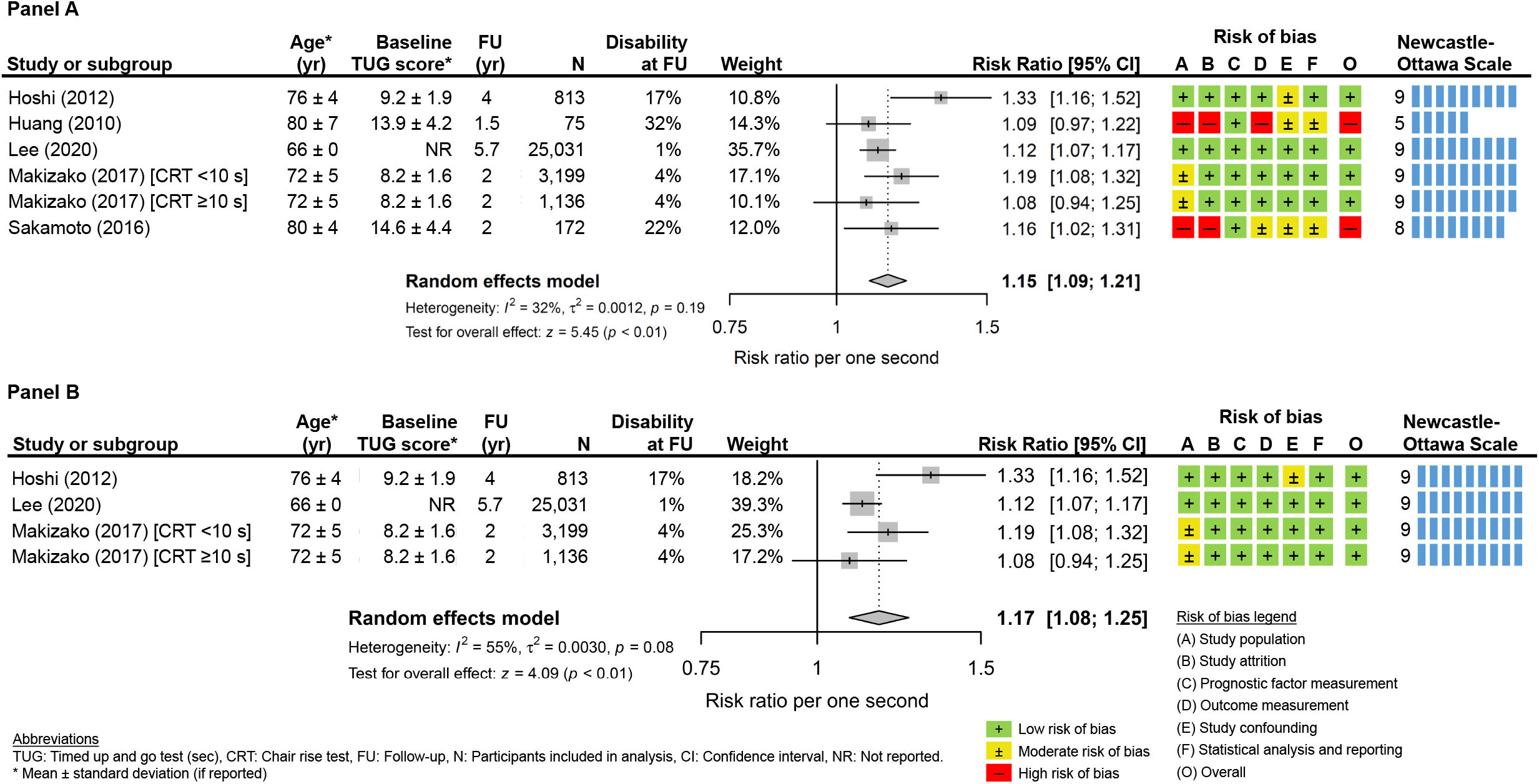
Association of Timed Up and Go test with incident disability (panel A: all studies; panel B: only studies with an overall low risk of bias)

#### Chair Rise Test

There was a statistically significant association between baseline Chair Rise Test performance and disability at follow-up (RR per 1-second increase = 1.07, 95% CI: 1.04–1.10, I^2^ = 61%, 7 reports, 9,450 participants, Figure 4, moderate certainty of evidence).^28,36,41,42,51,55,56^

#### Timed Up and Go test

The RR per 1-second increase in TUG performance (poorer performance) was 1.15 (95% CI: 1.09– 1.21, I^2^ = 32%, 5 reports, 30,426 participants, Figure 5 panel A, high certainty of evidence).^47-49,51,58^ This association remained stable when only reports with an overall low RoB were included in the analysis (RR = 1.17, 95% CI: 1.08–1.25, I^2^ = 55%, 3 reports, 30,179 participants, Figure 5 panel B).47,49,51

## Discussion

In this systematic review and meta-analysis of 40 reports, including more than 85,000 community-dwelling older adults, we found that mobility capacity was significantly associated with incident ADL disability. The pooled relative risk per one conventional unit was calculated for four common, feasible, standardized, and performance-based outcome assessments, including gait speed at usual and fast pace, SPPB, TUG, and CRT. The results are based on a heterogeneous sample of studies with an overall relatively high risk of bias.

Each 0.1 m/s decrease in usual and fast gait speed increased the risk of subsequent disability by 23% and 28%, respectively. Our results are in agreement with the findings of a large meta-analysis of individual participant data from 7 independent cohort studies,^13^ including 27,220 community-dwelling older adults. The age-adjusted relative risk reduction per 0.1 m/s greater speed at usual pace for bathing or dressing dependence over 3 years was 32% in men and 26% in women. In another meta-analysis, Wang et al.^7^ reported a pooled OR of 1.64 per 0.1 m/s lower speed for ADL disability (2 reports). The pooled OR for older people with “low vs. high” gait speed was 2.33 (6 reports).^7^ These associations differ from our findings since they are expressed in ORs, data on usual and fast speed was pooled, and the associations for different units of measurement were not converted to one conventional unit, as in the present analysis.^7^

For the SPPB, our meta-analysis of 11 reports suggests that each 1-point decrease increased the risk of incident disability by 30%. Our analysis extends the findings of two previous reviews reporting on the predictive ability of the SPPB for ADL disability.^7,67^ In the most recent review, the OR per 1-point decrease for incident disability was 1.12 (7 reports).^7^ In a prospective cohort study, the SPPB was associated with different disability subtypes, with adjusted HRs ranging from 1.10 for transient disability to 1.35 for long-term disability.^68^

Based on 7 reports, our findings suggest that each additional second needed to complete the CRT increases the risk of ADL disability by 7%. A previous review, which was based on 3 reports and dichotomized CRT results, also found a strong association (slow vs. fast, OR = 1.90).^7^

For TUG we found a pooled RR of 1.15 per 1-second increase in test completion that remained stable in the sensitivity analysis excluding those reports with a high risk of bias. A systematic review and a recent cohort study of 1,084,875 older adults found that slower TUG speed is associated with incident ADL disability in older adults.^7,69^

In essence, our analyses show that all included assessments of mobility capacity have a predictive value for ADL disability. Strong associations have also been reported between mobility and mortality by others.^69-72^

Assessments of mobility capacity should be implemented in regular health and/or physical evaluations of older adults to identify people at risk of disability. The general practitioner and other qualified health professionals may perform such checks in various clinical or community settings.^51,71^ Although we included only assessments considered feasible in such circumstances, feasibility varies between instruments. For example, the TUG cannot be completed by a significant number of community-dwelling older adults due to physical and/or cognitive abilities, and insufficient space can limit the testing in a patient’s home environment.^73^ Approximately 20% of community-dwelling older adults are unable to complete 5 chair rises.^74^ Gait speed assessment is quick and feasible, but not trivial. The testing protocol has a relevant impact on the recorded gait speed, especially the starting modalities (standing vs. walking), walking pace (usual vs. fast), distance and setting.^75,76^ For example, compared with real-world walking, patients consistently walk faster during ‘normal’ walking in (supervised) laboratory settings.^77^ In agreement with other reviews,^7,70^ we observed large heterogeneity in test protocols. In clinical care and research, a standardized test protocol is essential for obtaining a reliable and valid assessment of gait speed or other mobility parameters.^75^

Walking-related digital mobility outcomes or unsupervised assessment of mobility with mobile health technologies, such as real-world walking speed recorded with wearable sensors or chair rise activities recorded with domestic-integrated devices, may overcome the limitations of conventional (supervised) COAs, such as differences between gait speed test protocols.^78,79^ Moreover, digital (unsupervised) assessments of mobility appear to be more accurate and sensitive to change, and thus may emerge as complementary assessments to support risk stratification and monitoring of physical capacity of older people.^78^ Methods for the non-supervised assessment of gait speed and other digital gait parameters in an older individual’s usual environment are under development.^80,81^ The SPPB’s use for non-supervised assessment is doubtful.^82^

For clinical implementation, evidence of predictive validity for disability of each assessment should be considered. Some of the included studies performed direct comparisons between COA of mobility capacity. For example, Minneci et al.^52^ compared the prognostic value of usual gait speed and the SPPB, and reported non-significant differences between AUC values for incident disability (0.73 versus 0.71). The direct comparison by Makizako et al.^51^ showed that the predictive value of the TUG (HR = 2.24, 95% CI: 1.42–3.53) is better than of the CRT (HR = 1.88, 95% CI: 1.11–3.20).

COAs of mobility capacity have also been used in composite prediction models, which might be more accurate than single parameters alone. For example, Jonkman et al.^83^ included gait speed and CRT in a clinical prediction model for older people of 65–75 years that showed an AUC of 0.72 for the onset of functional decline. Another clinical prediction model that included CRT and balance performance in midlife (at 53 years) showed good performance (AUC = 0.74) for late-life disability, indicating that mobility capacity may improve risk stratification already at younger ages.^84^

## Limitations

This study has some limitations that are inherent to meta-analyses, including heterogeneity in populations and variable endpoint definitions across the included reports. For example, gait speed was assessed differently across the studies and ADL disability was measured in more than 10 ways. The generalizability of our findings is limited since people with cognitive impairment and people who deceased were excluded from some included studies. We could analyze data only from authors who replied to our request and the results might be influenced by reporting bias since trial registers were not searched and conference abstracts were not included. We did, however, perform an extensive screening of reference lists and related reviews.

## Conclusions

Our findings suggest that the SPPB, gait speed at usual or fast pace, the TUG, and the CRT identify people at risk of incident disability. Mobility impairment is a major and modifiable risk factor for the loss of autonomy, impaired quality of life, and mortality. It should be mandated as a quality indicator of health for older people.

## Data Availability

All data produced in the present work are contained in the manuscript.

## Author Contributions

Dr Braun and Dr Peter had full access to all of the data in the study and take responsibility for the integrity of the data and the accuracy of the data analysis.

Concept and design: Braun, Thiel, Becker, Grüneberg.

Acquisition of data: Braun, Bahns.

Analysis of data: Braun, Peter, Büchele.

Interpretation of data: Braun, Thiel, Peter, Bahns, Büchele, Rapp, Becker, Grüneberg.

Drafting of the manuscript: Braun.

Critical revision of the manuscript for important intellectual content: Braun, Thiel, Peter, Bahns, Büchele, Rapp, Becker, Grüneberg.

## Additional Contributions

Lisa Happe, Katja Ehrenbrusthoff, Elena Cramer, Kathrin Stiller, and Jérôme Camerlynck (HS Gesundheit Bochum) contributed to the data extraction. Gabriele Meyer (HS Gesundheit Bochum) contributed to the development of the search strategy.

