## Supplementary material for "Association of clinical outcome assessments of mobility capacity and incident disability in community-dwelling older adults - a systematic review and meta-analysis"

**Supplementary Online Content**

eTable 1: MEDLINE search strategy via OVID

eTable 2: EMBASE search strategy via OVID

eTable 3: CINAHL search strategy via EBSCO

eTable 4: Lists of the 16 published reviews from the authors' private archives

eTable 5: Reports identified through database searching and excluded after full-text assessment (n = 286)

eTable 6: Reports identified through other sources and excluded after full-text assessment (n = 43)

eTable 7: Key characteristics of the included reports (n = 40)

eTable 8: Additional characteristics of the included reports (n = 40)

eTable 9: Overview of cohort studies

eTable 10: Detailed description of gait speed assessment conduction in the included reports

eTable 11: Types of ADL components measured in the included reports using incident ADL disability as an outcome (n = 23)

eTable 12: Quality assessment of included reports using the Newcastle–Ottawa Scale (NOS)

eTable 13: Grading of Recommendations, Assessment, Development and Evaluations (GRADE) for each risk factor

eTable 14: Associations between baseline mobility capacity and disability at follow-up

eFigure 1: Flow chart

eFigure 2: Risk of bias summary

eFigure 3: Risk of bias graph

eFigure 4: Funnel plot for reports on usual gait speed

eFigure 5: Funnel plot for reports on the Short Physical Performance Battery (SPPB)

eText 1: Eligibility criteria

eText 2: Data extraction

eText 3: Additional information on the use of the Newcastle-Ottawa scale (NOS) in this review

### eTable 1: MEDLINE search strategy via OVID

| Search | Row | Query |
|---|---|---|
| Population | 1 | exp aged/ |
| | 2 | (aged or aging or ageing or elder* or frail* or geriatric* or old*).ti,ab. |
| | 3 | independent-living/ or homes for the aged/ or self care/ or homebound persons/ |
| | 4 | (independent-living or community-dwelling or community-living or "home for the aged" or self-care or home-bound).ti,ab. |
| | 5 | 3 or 4 |
| | 6 | 2 and 5 |
| | 7 | 1 or 6 |
| Mobility assessment | 8 | exp walking/ or gait/ or postural balance/ or physical functional performance/ or (walking or gait or "body equilibrium" or performance or ambulation or balance or "physical function*" or "lower body function*" or "lower-extremity function*" or mobility).ti,ab. |
| | 9 | walk test/ or gait analysis/ or "outcome assessment (Health Care)"/ or geriatric assessment/ or treatment outcome/ or ("walk test" or "gait analysis" or "physical performance" or "outcome assessment" or "functional assessment" or "geriatric assessment" or "treatment outcome" or "10-meter walk" or "ten-meter walk" or "10-MWT" or "6-meter walk" or "six-meter walk" or "6-MWT" or "chair-ris*" or "chair-stand" or "sit-to-stand" or "outcome measure*" or "performance test" or "performance measure*" or "performance-based functional measure*" or SPPB or "timed-up-and-go" or "timed-get-up-and-go" or TUG or "stair-climb* power").ti,ab. |
| | 10 | 8 and 9 |
| Disability | 11 | "Activities of daily living"/ or mobility limitation/ or nursing homes/ or (ADL or "daily activit*" or "daily life" or "daily living" or IADL or "Barthel-Index" or "Berg Balance Scale" or disability or disabilities or disablement or "functional independence" or "functional status" or "low-function*" or LLFDI or "lower-extremity limitation*" or "inability to walk" or "task difficult*" or "care need" or "need for help" or "nursing home*").ti,ab. or ((functional or mobility or physical or walking or balance) adj2 (impairment* or difficult* or limitation* or decline or capa* or abilit* or performance)).ti,ab. |
| Prediction | 12 | Validat$.ti,ab. or Predict$.ti. or Rule$.ti,ab. or (Predict$ and (Outcome$ or Risk$ or Model$)).ti,ab. or ((History or Variable$ or Criteria or Scor$ or Characteristic$ or Finding$ or Factor$) and (Predict$ or Model$ or Decision$ or Identif$ or Prognos$)).ti,ab. or (Decision$.ti,ab. and ((Model$ or Clinical$).ti,ab. or Logistic Models/)) or (Prognostic and (History or Variable$ or Criteria or Scor$ or Characteristic$ or Finding$ or Factor$ or Model$)).ti,ab. |
| | 13 | "Stratification".ti,ab. or "ROC Curve".sh. or "Discrimination".ti,ab. or "Discriminate".ti,ab. or "c-statistic".ti,ab. or "c statistic".ti,ab. or "Area under the curve".ti,ab. or "AUC".ti,ab. or "Calibration".ti,ab. or "Indices".ti,ab. or "Algorithm".ti,ab. or "Multivariable".ti,ab. |
| | 14 | 12 or 13 |
| | 15 | 7 and 10 and 11 and 14 |
| | 16 | 15 not (infant/ or child/ or young adult/ or middle aged/) |
| | 17 | (controlled clinical trial or meta analysis or randomized controlled trial or "review" or "systematic review").pt. |
| | 18 | 16 not 17 |
| | 19 | exp animals/ not humans.sh. |
| | 20 | 18 not 19 |

#### eTable 2: EMBASE search strategy via OVID

| Search | Row | Query |
|---|---|---|
| Population | 1 | exp aged/ |
| | 2 | (aged or aging or ageing or elder* or frail* or geriatric* or old*).ti,ab. |
| | 3 | independent living/ or community living/ or community dwelling person/ or home for the aged/ or self care/ or homebound patient/ |
| | 4 | (independent-living or community-living or community-dwelling or "home for the aged" or self-care or home-bound).ti,ab. |
| | 5 | 3 or 4 |
| | 6 | 2 and 5 |
| | 7 | 1 or 6 |
| Mobility assessment | 8 | exp walking/ or gait/ or body equilibrium/ or performance/ or (walking or gait or "body equilibrium" or performance or ambulation or balance or "physical function*" or "lower body function*" or "lower-extremity function*" or mobility).ti,ab. |
| | 9 | walk test/ or physical performance/ or outcome assessment/ or functional assessment/ or geriatric assessment/ or treatment outcome/ or ("walk test" or "gait analysis" or "physical performance" or "outcome assessment" or "functional assessment" or "geriatric assessment" or "treatment outcome" or "10-meter walk" or "ten-meter walk" or "10-MWT" or "6-meter walk" or "six-meter walk" or "6-MWT" or "chair-ris*" or "chair-stand" or "sit-to-stand" or "outcome measure*" or "performance test" or "performance measure*" or "performance-based functional measure*" or SPPB or "timed-up-and-go" or "timed-get-up-and-go" or TUG or "stair-climb* power").ti,ab. |
| | 10 | 8 and 9 |
| Disability | 11 | Daily life activity/ or Barthel Index/ or ADL disability/ or walking difficulty/ or balance impairment/ or Berg Balance Scale/ or nursing home/ or (ADL or "daily activit*" or "daily life" or "daily living" or IADL or "Barthel-Index" or "Berg Balance Scale" or disability or disabilities or disablement or "functional independence" or "functional status" or "low-function*" or LLFDI or "lower-extremity limitation*" or "inability to walk" or "task difficult*" or "care need" or "need for help" or "nursing home*").ti,ab. or ((functional or mobility or physical or walking or balance) adj2 (impairment* or difficult* or limitation* or decline or capa* or abilit* or performance)).ti,ab. |
| Prediction | 12 | Validat$.mp. or Predict$.ti. or Rule$.mp. or (Predict$ and (Outcome$ or Risk$ or Model$)).mp. or ((History or Variable$ or Criteria or Scor$ or Characteristic$ or Finding$ or Factor$) and (Predict$ or Model$ or Decision$ or Identif$ or Prognos$)).mp. or (Decision$.mp. and ((Model$ or Clinical$).mp. or statistical model/)) or (Prognostic and (History or Variable$ or Criteria or Scor$ or Characteristic$ or Finding$ or Factor$ or Model$)).mp. |
| | 13 | Stratification.mp. or receiver operating characteristic/ or Discrimination.mp. or Discriminate.mp. or "c-statistic".mp. or "c statistic".mp. or "Area under the curve".mp. or AUC.mp. or Calibration.mp. or Indices.mp. or Algorithm.mp. or Multivariable.mp. |
| | 14 | 12 or 13 |
| | 15 | 7 and 10 and 11 and 14 |
| | 16 | 15 not (infant/ or child/ or young adult/ or middle aged/) |
| | 17 | (controlled clinical trial or meta analysis or randomized controlled trial or "review" or "systematic review").pt. |
| | 18 | 16 not 17 |
| | 19 | (exp animals/ or nonhuman/) not human/ |
| | 20 | 18 not 19 |

### eTable 3: CINAHL search strategy via EBSCO

| Search | Row | Query |
|---|---|---|
| Population | 1 | (MH "Aged+") |
| | 2 | TI ( (aged or aging or ageing or elder* or frail* or pre-frail* or geriatric* or old*) ) OR AB ( (aged or aging or ageing or elder* or frail* or geriatric* or old*) ) |
| | 3 | (MH "Community Living") |
| | 4 | TI ( (independent-living or community-dwelling or community-living or "home for the aged" or self-care or home-bound ) ) OR AB ( (independent-living or community-dwelling or community-living or "home for the aged" or self-care or home-bound ) ) |
| | 5 | S3 OR S4 |
| | 6 | S2 AND S5 |
| | 7 | S1 OR S6 |
| Mobility assessment | 8 | (MH "Walking+") OR (MH "Balance, Postural") OR (MH "Physical Performance") |
| | 9 | TI ( (walking or gait or "body equilibrium" or performance or ambulation or balance or "physical function*" or "lower body function*" or "lower-extremity function*" or mobility) ) OR AB ( (walking or gait or "body equilibrium" or performance or ambulation or balance or "physical function*" or "lower body function*" or "lower-extremity function*" or mobility) ) |
| | 10 | S8 OR S9 |
| | 11 | (MH "Gait Analysis") OR (MH "outcome assessment") OR (MH "geriatric assessment") OR (MH "treatment outcomes") |
| | 12 | TI ( ("walk test" or "gait analysis" or "physical performance " or "outcome assessment" or "functional assessment" or "geriatric assessment" or "treatment outcome" or "10-meter walk" or "ten-meter walk" or "10-MWT" or "6-meter walk" or "six-meter walk" or "6-MWT" or "chair-ris*" or "chair-stand" or "sit-to-stand" or "outcome measure*" or "performance test" or "performance measure*" or "performance-based functional measure*" or SPPB or "timed-up-and-go" or "timed-get-up-and-go" or TUG or "stair-climb* power")) or AB ( ("walk test" or "gait analysis" or "physical performance " or "outcome assessment" or "functional assessment" or "geriatric assessment" or "treatment outcome" or "10-meter walk" or "ten-meter walk" or "10-MWT" or "6-meter walk" or "six-meter walk" or "6-MWT" or "chair-ris*" or "chair-stand" or "sit-to-stand" or "outcome measure*" or "performance test" or "performance measure*" or "performance-based functional measure*" or SPPB or "timed-up-and-go" or "timed-get-up-and-go" or TUG or "stair-climb* power")) |
| | 13 | S11 OR S12 |
| | 14 | S10 AND S13 |
| Disability | 15 | (MH "Activities of Daily Living") OR (MH "Barthel Index") OR (MH "Nursing homes") |
| | 16 | TI ( (ADL or "daily activit*" or "daily life" or "daily living" or IADL or "Barthel-Index" or "Berg Balance Scale" or disability or disabilities or disablement or "functional independence" or "functional status" or "low-function*" or LLFDI or "lower-extremity limitation*" or "inability to walk" or "task difficult*" or "care need" or "need for help" or "nursing home*") ) OR AB ( (ADL or "daily activit*" or "daily life" or "daily living" or IADL or "Barthel-Index" or "Berg Balance Scale" or disability or disabilities or disablement or "functional independence" or "functional status" or "low-function*" or LLFDI or "lower-extremity limitation*" or "inability to walk" or "task difficult*" or "care need" or "need for help" or "nursing home*") ) |
| | 17 | TI ( (functional or mobility or physical or walking or balance) N2 (impairment* or difficult* or limitation* or decline or capa* or abilit* or performance) ) OR AB ( (functional or mobility or physical or walking or balance) N2 (impairment* or difficult* or limitation* or decline or capa* or abilit* or performance) ) |
| | 18 | S15 OR S16 OR S17 |
| Prediction | 19 | TI ( (Validat$ OR Rule$) ) OR AB ( (Validat$ OR Rule$) ) OR TI Predict$ |

| Search | Row | Query |
|---|---|---|
| | 20 | TI ( (Predict$) AND TI ( (Outcome$ or risk$ or Factor$) ) ) OR AB ( (Predict$) AND AB ( (Outcome$ or risk$ or Factor$) ) ) |
| | 21 | TI ( (History OR Variable$ OR Criteria OR Scor$ OR Characteristic$ OR Finding$ OR Factor$) ) OR AB ( (History OR Variable$ OR Criteria OR Scor$ OR Characteristic$ OR Finding$ OR Factor$) ) |
| | 22 | TI ( (Predict$ OR Model$ OR Decision$ OR Identif$ OR Prognos$) ) OR AB ( (Predict$ OR Model$ OR Decision$ OR Identif$ OR Prognos$) ) |
| | 23 | TI ( (Decision$) AND TI ( ( Model$ OR Clinical$ OR "Logistic Model$") ) ) OR AB ( (Decision$) AND AB ( (Model$ OR Clinical$ OR "Logistic Model$") ) ) |
| | 24 | TI ( (Prognostic) AND TI ( (History OR Variable$ OR Criteria OR Scor$ OR Characteristic$ OR Finding$ OR Factor$ OR Model$) ) ) OR AB ( (Prognostic) AND AB ( (History OR Variable$ OR Criteria OR Scor$ OR Characteristic$ OR Finding$ OR Factor$ OR Model$) ) ) |
| | 25 | S19 OR S20 OR (S21 AND S22 OR S23) OR S24 |
| | 26 | (MH "ROC Curve") OR TI ( (Stratification OR Discrimination OR Discriminate OR c-statistic OR "c statistic" OR "Area under the curve" OR AUC OR Calibration OR Indices OR Algorithm OR Multivariable") ) OR AB ( (Stratification OR Discrimination OR Discriminate OR c-statistic OR "c statistic" OR "Area under the curve" OR AUC OR Calibration OR Indices OR Algorithm OR Multivariable") ) |
| | 27 | S25 OR S26 |
| | 28 | S7 AND S14 AND S18 AND S27 |
| | 29 | (MH "Infant") OR (MH "Child") OR (MH "young adult") OR (MH "middle age") |
| | 30 | S28 NOT S29 |
| | 31 | PT meta analysis or randomized controlled trial or review or systematic review |
| | 32 | S30 NOT S31 |
| | 33 | (MH "Animals+) |
| | 34 | S32 NOT S33 |

**eTable 4: Lists of the 16 published reviews from the authors' private archives focusing on the prognostic value of mobility assessments for health outcomes and/or risk factors for disability in older adults**

| Authors | Year | Review title | References screened (title) | References screened (full-text) |
|---|---|---|---|---|
| Abellan van Kan et al. | 2009 | Gait speed at usual place as a predictor of adverse outcomes in community-dwelling older people – an international academy on nutrition and aging (IANA) task force | 52 | 10 |
| Cavanaugh et al. | 2018 | The predictive validity of physical performance measures in determining markers of preclinical disability in community-dwelling middle-aged and older adults: a systematic review | 38 | 7 |
| Freiberger et al. | 2012 | Performance-based physical function in older community-dwelling persons: a systematic review of instruments | 69 | 5 |
| Gaugler et al. | 2007 | Predicting nursing home admission in the U.S: a meta-analysis | 50 | 16 |
| Gaugler et al. | 2009 | Predictors of nursing home admission for persons with dementia | 50 | 2 |
| Gawel et al. | 2018 | The short physical performance battery as a predictor for long term disability or institutionalization in the community dwelling population aged 65 years old or older | 36 | 6 |
| Koijma | 2017 | Frailty as a predictor of disabilities among community-dwelling older people: a systematic review and meta-analysis | 44 | 3 |
| Luppa et al. | 2009 | Prediction of institutionalization in the elderly. A systematic review | 52 | 16 |
| O'Caoimh et al. | 2015 | Risk prediction in the community: A systematic review of case-finding instruments that predict adverse healthcare outcomes in community-dwelling older adults | 63 | 0 |
| Pamoukdjian et al. | 2015 | Measurement of gait speed in older adults to identify complications associated with frailty: A systematic review | 61 | 4 |
| Peel et al. | 2012 | Gait speed as a measure in geriatric assessment in clinical settings: a systematic review | 29 | 2 |
| Savino et al. | 2014 | Assessment of mobility status and risk of mobility disability in older persons | 128 | 19 |
| Stuck et al. | 1999 | Risk factors for functional status decline in community-living elderly people: a systematic review | 152 | 22 |
| Tas et al. | 2007 | Prognostic factors of disability in older people: a systematic review | 18 | 0 |
| Vermeulen et al. | 2011 | Predicting ADL disability in community-dwelling elderly people using physical frailty indicators: a systematic review | 45 | 13 |
| Wang et al. | 2020 | Muscle mass, strength, and physical performance predicting activities of daily living: a meta-analysis | 161 | 46 |
| | | Sum | 1048 | 171 |

**eTable 5: Reports identified through database searching and excluded after full-text assessment (n = 286)**

| Author(s) | Year | Title | Reason for exclusion |
|---|---|---|---|
| Abellan Van Kan et al. | 2009 | Gait speed at usual pace as a predictor of adverse outcomes in community-dwelling older people an International Academy on Nutrition and Aging (IANA) task force | 1 |
| Adachi et al. | 2019 | Estimation of reduced walking speed using simple measurements of physical and psychophysiological function in community-dwelling elderly people: a cross-sectional and longitudinal study | 6 |
| Albert et al. | 2015 | Declines in mobility and changes in performance in the instrumental activities of daily living among mildly disabled community-dwelling older adults | 6 |
| Amigues et al. | 2013 | Low skeletal muscle mass and risk of functional decline in elderly community-dwelling women: the prospective EPIDOS study | 6 |
| Antonini et al. | 2016 | Impact of functional determinants on 5.5-year mortality in Amazon riparian elderly | 6 |
| Arnau et al. | 2016 | Risk factors for functional decline in a population aged 75 years and older without total dependence: A one-year follow-up | 4 |
| Arve et al. | 2006 | Physical functioning, health and survival: a ten-year follow-up study | 7 |
| Atkinson et al. | 2005 | Predictors of combined cognitive and physical decline | 4 |
| Auais et al. | 2018 | Fear of Falling Predicts Incidence of Functional Disability 2 Years Later: A Perspective From an International Cohort Study | 5 |
| Avila-Funes et al. | 2008 | Frailty among community-dwelling elderly people in France: the three-city study | 5 |
| Avlund et al. | 1995 | Changes in functional ability from ages 70 to 75: A Danish longitudinal study | 5 |
| Ayis et al. | 2006 | Predicting catastrophic decline in mobility among older people | 4 |
| Bachettini et al. | 2019 | Sarcopenia as a mortality predictor in community-dwelling older adults: a comparison of the diagnostic criteria of the European Working Group on Sarcopenia in Older People | 6 |
| Baker et al. | 2018 | Estimation of Skeletal Muscle Mass Relative to Adiposity Improves Prediction of Physical Performance and Incident Disability | 5 |
| Bannerman et al. | 2002 | Anthropometric indices predict physical function and mobility in older Australians: The Australian longitudinal study of ageing | 5 |
| Barbour et al. | 2016 | Trajectories of Lower Extremity Physical Performance: Effects on Fractures and Mortality in Older Women | 6 |
| Batko-Szwaczka et al. | 2020 | Predicting adverse outcomes in healthy aging community-dwelling early-old adults with the timed up and go test | 1 |
| Batsis et al. | 2015 | Impact of obesity on disability, function, and physical activity: Data from the Osteoarthritis Initiative | 5 |
| Bea et al. | 2018 | Body composition and physical function in the Women's Health Initiative Observational Study | 5 |
| Beauchamp et al. | 2015 | How Should Disability Be Measured in Older Adults? An Analysis from the Boston Rehabilitative Impairment Study of the Elderly | 5 |
| Beauchamp et al. | 2014 | What physical attributes underlie self-reported vs. observed ability to walk 400 m in later life? An analysis from the InCHIANTI Study | 6 |
| Beaudart et al. | 2017 | Sarcopenia in community dwelling subjects: The sarcophage study | 9 |
| Beckett et al. | 1996 | Analysis of change in self-reported physical function among older persons in four population studies | 5 |
| Ben-Ezra et al. | 2006 | Predictors of mortality in the old-old in Israel: the Cross-sectional and Longitudinal Aging Study | 6 |

| Author(s) | Year | Title | Reason for exclusion |
|---|---|---|---|
| Best et al. | 2018 | Longitudinal Associations Between Walking Speed and Amount of Self-reported Time Spent Walking Over a 9-Year Period in Older Women and Men | 7 |
| Bianchi et al. | 2016 | The Predictive Value of the EWGSOP Definition of Sarcopenia: Results From the InCHIANTI Study | 5 |
| Bimou et al. | 2021 | Patterns and predictive factors of loss of the independence trajectory among community-dwelling older adults | 1 |
| Bjorkman et al. | 2019 | Bioimpedance analysis and physical functioning as mortality indicators among older sarcopenic people | 6 |
| Bjornsbo et al. | 2002 | Changes in physical performance in elderly Europeans. SENECA 1993 - 1999 | 1 |
| Blahak et al. | 2011 | Balance disturbances and falls predict rapid transition to disability in patients with age-related white matter changes - Results of the 6-year follow-up of the LADIS study | 9 |
| Blain et al. | 2010 | Balance and walking speed predict subsequent 8-year mortality independently of current and intermediate events in well-functioning women aged 75 years and older | 7 |
| Boeckxstaens et al. | 2015 | Multimorbidity measures were poor predictors of adverse events in patients aged >=80 years: A prospective cohort study | 5 |
| Botoseneanu et al. | 2016 | Sex Differences in Concomitant Trajectories of Self-Reported Disability and Measured Physical Capacity in Older Adults | 7 |
| Botoseneanu et al. | 2013 | Long-term trajectories of lower extremity function in older adults: estimating gender differences while accounting for potential mortality bias | 6 |
| Brach et al. | 2012 | Use of stance time variability for predicting mobility disability in community-dwelling older persons: a prospective study | 5 |
| Brach et al. | 2004 | The relationship among physical activity, obesity, and physical function in community-dwelling older women | 5 |
| Brown et al. | 2020 | Trajectories of short physical performance battery are strongly associated with future major mobility disability: Results from the life study | 6 |
| Bruhl et al. | 2012 | Validity and internal consistency of mobility scales for healthy older people in Germany | 6 |
| Buatois et al. | 2008 | Five times sit to stand test is a predictor of recurrent falls in healthy community-living subjects aged 65 and older | 6 |
| Buchman et al. | 2019 | Different combinations of mobility metrics derived from a wearable sensor are associated with distinct health outcomes in older adults | 1 |
| Buchman et al. | 2016 | Motor Function Is Associated With Incident Disability in Older African Americans | 5 |
| Buchman et al. | 2011 | Combinations of motor measures more strongly predict adverse health outcomes in old age: The rush memory and aging project, a community-based cohort study | 5 |
| Buchman et al. | 2007 | Physical activity and leg strength predict decline in mobility performance in older persons | 6 |
| Callisaya et al. | 2018 | The Association of Clinic-Based Mobility Tasks and Measures of Community Performance and Risk | 1 |
| Carriere et al. | 2005 | Hierarchical components of physical frailty predicted incidence of dependency in a cohort of elderly women | 7 |
| Cawthon et al. | 2011 | Clustering of strength, physical function, muscle, and adiposity characteristics and risk of disability in older adults | 1 |
| Cesari et al. | 2004 | Inflammatory markers and physical performance in older persons: the InCHIANTI study | 5 |
| Cesari et al. | 2009 | Self-assessed health status, walking speed and mortality in older Mexican-Americans | 6 |
| Cesari et al. | 2008 | Physical function and self-rated health status as predictors of mortality: results from longitudinal analysis in the ilSIRENTE study | 6 |

| Author(s) | Year | Title | Reason for exclusion |
|---|---|---|---|
| Cesari et al. | 2015 | Sarcopenia-related parameters and incident disability in older persons: results from the "invecchiare in Chianti" study | 7 |
| Cesari et al. | 2009 | Added value of physical performance measures in predicting adverse health-related events: results from the Health, Aging And Body Composition Study | 6 |
| Cesari et al. | 2005 | Prognostic value of usual gait speed in well-functioning older people -- results from the health, aging and body composition study | 6 |
| Chang et al. | 2004 | Incidence of loss of ability to walk 400 meters in a functionally limited older population | 6 |
| Chaves et al. | 2000 | Predicting the risk of mobility difficulty in older women with screening nomograms: the Women's Health and Aging Study II | 6 |
| Chen et al. | 2015 | Identifying factors associated with changes in physical functioning in an older population | 4 |
| Chen et al. | 2012 | Predicting Cause-Specific Mortality of Older Men Living in the Veterans Home by Handgrip Strength and Walking Speed: A 3-Year, Prospective Cohort Study in Taiwan | 6 |
| Cheung et al. | 2016 | Evaluation of Cutpoints for Low Lean Mass and Slow Gait Speed in Predicting Death in the National Health and Nutrition Examination Survey 1999-2004 | 6 |
| Chivite et al. | 2013 | Utility of geriatric assessment to predict mortality in the oldest old: The octabaix study 3-year follow-up | 6 |
| Chock et al. | 2019 | Handgrip strength in late life predicts future functional impairment: The kuakini honolulu-asia aging study | 9 |
| Clark et al. | 2015 | Improving the validity of activity of daily living dependency risk assessment | 1 |
| Clark et al. | 1998 | Predictors of mobility and basic ADL difficulty among adults aged 70 years and older | 5 |
| Corsonello et al. | 2012 | Prognostic significance of the short physical performance battery in older patients discharged from acute care hospitals | 2 |
| Costanzo et al. | 2018 | Clusters of functional domains to identify older persons at risk of disability | 5 |
| Covinsky et al. | 2006 | Development and validation of an index to predict activity of daily living dependence in community-dwelling elders | 5 |
| Dalle Carbonare et al. | 2009 | Physical disability and depressive symptomatology in an elderly population: a complex relationship. The Italian Longitudinal Study on Aging (ILSA) | 5 |
| Danilovich et al. | 2015 | Performance measures, hours of caregiving assistance, and risk of adverse care outcomes among older adult users of medicaid home and community-based services | 1 |
| Dapp et al. | 2013 | Correlates of frailty, prediction of functional decline and preventative approaches-selected results from the lausanne cohort 65+ (Lc65+) study (switzerland) (lausanne) and the longitudinal urban cohort ageing study (lucas) (Germany) | 9 |
| Davis et al. | 2015 | Mobility predicts change in older adults' health-related quality of life: evidence from a Vancouver falls prevention prospective cohort study | 6 |
| De Buyser et al. | 2013 | Physical function measurements predict mortality in ambulatory older men | 6 |
| De Buyser et al. | 2016 | Three year functional changes and long-term mortality hazard in community-dwelling older men | 6 |
| De Carvalho Bastone et al. | 2018 | The Use of the Incremental Shuttle Walk Test to Identify Instrumental Activity Daily Living Disability and to Predict Peak Oxygen Consumption in Older Adults: A Methodological Study | 1 |
| de la Torre-Luque et al. | 2020 | Functioning profiles in a nationally representative cohort of Spanish older adults: A latent class study | 5 |

| Author(s) | Year | Title | Reason for exclusion |
|---|---|---|---|
| Delmonico et al. | 2007 | Alternative definitions of sarcopenia, lower extremity performance, and functional impairment with aging in older men and women | 5 |
| den Ouden et al. | 2013 | Identification of high-risk individuals for the development of disability in activities of daily living. A ten-year follow-up study | 7 |
| Denkinger et al. | 2012 | Physical activity and other health-related factors predict health care utilisation in older adults: the ActiFE Ulm study | 5 |
| Deshpande et al. | 2008 | Activity restriction induced by fear of falling and objective and subjective measures of physical function: a prospective cohort study | 5 |
| Deshpande et al. | 2013 | Predicting 3-year incident mobility disability in middle-aged and older adults using physical performance tests | 6 |
| Di Bari et al. | 2006 | Predictive validity of measures of comorbidity in older community dwellers: the Insufficienza Cardiaca negli Anziani Residenti a Dicomano Study | 5 |
| Diekmann et al. | 2020 | Minimizing comprehensive geriatric assessment to identify deterioration of physical performance in a healthy community-dwelling older cohort: longitudinal data of the AEQUIPA Versa study | 6 |
| Diez-Ruiz et al. | 2016 | Factors associated with frailty in primary care: a prospective cohort study | 6 |
| Ding et al. | 2017 | Predictive Validity of Two Physical Frailty Phenotype Specifications Developed for Investigation of Frailty Pathways in Older People | 5 |
| Doi et al. | 2016 | Insulin-Like Growth Factor-1 Related to Disability Among Older Adults | 5 |
| Donoghue et al. | 2017 | Self-Reported Unsteadiness Predicts Fear of Falling, Activity Restriction, Falls, and Disability | 5 |
| Donoghue et al. | 2013 | Is timed up-and-go a better predictor of difficulty in daily activities than gait speed? | 9 |
| Donoghue et al. | 2013 | Using timed up-and-go to identify future difficulty in activities of daily living | 9 |
| Dutta et al. | 2011 | Predictors of extraordinary survival in the Iowa established populations for epidemiologic study of the elderly: cohort follow-up to "extinction" | 6 |
| Eggermont et al. | 2014 | Pain characteristics associated with the onset of disability in older adults: The maintenance of balance, independent living, intellect, and zest in the elderly boston study | 5 |
| Engedal et al. | 1996 | Mortality in the elderly - A 3-year follow-up of an elderly community sample | 6 |
| Ensrud et al. | 2008 | Comparison of 2 frailty indexes for prediction of falls, disability, fractures, and death in older women | 5 |
| Ensrud et al. | 2016 | Effects of Mobility and Cognition on Risk of Mortality in Women in Late Life: A Prospective Study | 6 |
| Fairhall et al. | 2014 | Predicting participation restriction in community-dwelling older men: the Concord Health and Ageing in Men Project | 6 |
| Falconer et al. | 1992 | Self-reported functional status predicts change in level of care in independent living residents of a continuing care retirement community | 5 |
| Fallah et al. | 2011 | Transitions in frailty status in older adults in relation to mobility: a multistate modeling approach employing a deficit count | 6 |
| Fanning et al. | 2020 | Relationships Between Profiles of Physical Activity and Major Mobility Disability in the LIFE Study | 6 |
| Farsijani et al. | 2017 | Even mealtime distribution of protein intake is associated with greater muscle strength, but not with 3-y physical function decline, in free-living older adults: the Quebec longitudinal study on Nutrition as a Determinant of Successful Aging (NuAge study) | 5 |
| Faurot et al.. | 2015 | Using claims data to predict dependency in activities of daily living as a proxy for frailty | 5 |

| Author(s) | Year | Title | Reason for exclusion |
|---|---|---|---|
| Federman et al. | 2010 | Development of and recovery from difficulty with activities of daily living: an analysis of national data | 5 |
| Feng et al. | 2010 | Effect of new disability subtype on 3-year mortality in Chinese older adults | 6 |
| Ferrer et al. | 2015 | Predicting factors of health-related quality of life in octogenarians: a 3-year follow-up longitudinal study | 6 |
| Ferrucci et al. | 2000 | Characteristics of nondisabled older persons who perform poorly in objective tests of lower extremity function | 6 |
| Fong et al. | 2015 | Disaggregating activities of daily living limitations for predicting nursing home admission | 5 |
| Formiga et al. | 2007 | Risk factors for functional decline in nonagenarians: A one-year follow-up - The Nonasantfeliu study | 2 |
| Formiga et al. | 2010 | Decline in the performance of activities of daily living over three years of follow-up in nonagenarians: The NonaSantfeliu study | 4 |
| Formiga et al. | 2016 | Evidence of functional declining and global comorbidity measured at baseline proved to be the strongest predictors for long-term death in elderly community residents aged 85 years: a 5-year follow-up evaluation, the OCTABAIX study | 6 |
| Formiga et al. | 2012 | The challenge of maintaining successful aging at 87 years old: the Octabaix study two-year follow-up | 6 |
| Fortes-Filho et al. | 2020 | Role of Gait Speed, Strength, and Balance in Predicting Adverse Outcomes of Acutely Ill Older Outpatients | 4 |
| Fox et al. | 2015 | Objectively assessed physical activity and lower limb function and prospective associations with mortality and newly diagnosed disease in UK older adults: an OPAL four-year follow-up study | 6 |
| Fragoso et al. | 2008 | Peak expiratory flow as a predictor of subsequent disability and death in community-living older persons | 5 |
| Fried et al. | 2000 | Preclinical mobility disability predicts incident mobility disability in older women | 6 |
| Fujiwara et al. | 2008 | Predictors of improvement or decline in instrumental activities of daily living among community-dwelling older Japanese | 5 |
| Fujiwara et al. | 2006 | Physical and psychological predictors for the onset of certification of long-term care insurance among older adults living independently in a community a 40-month follow-up study. [Japanese] | 8 |
| Gallucci et al. | 2019 | Artificial Neural Networks Help to Better Understand the Interplay between Cognition, Mediterranean Diet, and Physical Performance: Clues from TRELONG Study | 6 |
| Giampaoli et al.. | 1999 | Hand-grip strength predicts incident disability in non-disabled older men | 5 |
| Gill et al. | 2018 | Prognostic Effect of Changes in Physical Function Over Prior Year on Subsequent Mortality and Long-Term Nursing Home Admission | 5 |
| Gill et al. | 2018 | Development and Validation of a Functional Outcome Measure in the National Health and Aging Trends Study | 5 |
| Gill et al. | 1997 | The role of change in physical performance in determining risk for dependence in activities of daily living among nondisabled community-living elderly persons | 5 |
| Gill et al. | 1999 | The combined effects of baseline vulnerability and acute hospital events on the development of functional dependence among community-living older persons | 5 |
| Gill et al. | 2003 | Restricted activity and functional decline among community-living older persons | 5 |
| Gill et al. | 2009 | Risk factors for disability subtypes in older persons | 7 |
| Gill et al. | 2012 | Risk factors and precipitants of long-term disability in community mobility: a cohort study of older persons | 6 |
| Gill et al. | 2020 | Risk factors and precipitants of severe disability among community-living older persons | 7 |
| Gill et al.. | 2010 | Change in disability after hospitalization or restricted activity in older persons | 7 |
| Giuliani et al. | 2008 | Physical performance characteristics of assisted living residents and risk for adverse health outcomes | 2 |
| Gray et al. | 2016 | Rates and predictors of three-year mortality in older people in rural Tanzania | 6 |

| Author(s) | Year | Title | Reason for exclusion |
|---|---|---|---|
| Griebling et al. | 2014 | Re: Physical performance measures as a useful indicator of multiple geriatric syndromes in women aged 75 years and older: Editorial comment | 9 |
| Groessl et al. | 2019 | Physical Activity and Performance Impact Long-term Quality of Life in Older Adults at Risk for Major Mobility Disability | 6 |
| Guet et al. | 2010 | Factors associated with functional status in community-dwelling Hispanic elders, in east little Havana, FL | 4 |
| Guralnik et al. | 1994 | A short physical performance battery assessing lower extremity function: association with self-reported disability and prediction of mortality and nursing home admission | 1 |
| Guralnik et al. | 2001 | Progressive versus catastrophic loss of the ability to walk: implications for the prevention of mobility loss | 5 |
| Hamalainen et al. | 2006 | Predictive value of health-related fitness tests for self-reported mobility difficulties among high-functioning elderly men and women | 6 |
| Hermsen et al. | 2014 | Trajectories of physical functioning and their prognostic indicators: a prospective cohort study in older adults with joint pain and comorbidity | 5 |
| Hibbs et al. | 2012 | Curved path walking predicts 12-month physical function in older adults without mobility limited physical function | 9 |
| Hirani et al. | 2015 | Sarcopenia Is Associated With Incident Disability, Institutionalization, and Mortality in Community-Dwelling Older Men: The Concord Health and Ageing in Men Project | 5 |
| Hirsch et al. | 2012 | Predicting late-life disability and death by the rate of decline in physical performance measures | 7 |
| Hirvensalo et al. | 2000 | Mobility difficulties and physical activity as predictors of mortality and loss of independence in the community-living older population | 7 |
| Ho et al. | 1997 | Predictors of mobility decline: The Hong Kong old-old study | 6 |
| Hoeymans et al. | 1996 | Measuring functional status: cross-sectional and longitudinal associations between performance and self-report (Zutphen Elderly Study 1990-1993) | 4 |
| Hong et al. | 2018 | Low peak jump power is associated with elevated odds of dysmobility syndrome in community-dwelling elderly individuals: the Korean Urban Rural Elderly (KURE) study | 6 |
| Houles et al. | 2010 | Gait speed at usual pace as a predictor of adverse outcomes in community-dwelling older people. [French] | 8 |
| Hsu et al. | 2017 | Predicting emerging care-need with simple functional indicators: Findings from a national cohort study in Taiwan | 5 |
| Hsu et al. | 2019 | Predictors of developing a new need for long-term care of older adults aged >=70 years: Results from a population-based cohort study in Taiwan | 5 |
| Huang et al. | 2007 | Using physical performance measures to predict the onset of basic ADL disabilities in community-dwelling older adults...Combined Sections Meeting 2008: section on geriatrics poster and platform presentations. February 6-9, 2008, Nashville, TN | 9 |
| Idland et al. | 2013 | Physical performance as long-term predictor of onset of activities of daily living (ADL) disability: a 9-year longitudinal study among community-dwelling older women | 7 |
| Idland et al. | 2013 | Predictors of mobility in community-dwelling women aged 85 and older | 7 |
| Inzitari et al. | 2006 | Risk and predictors of motor-performance decline in a normally functioning population-based sample of elderly subjects: the Italian Longitudinal Study on Aging | 5 |

| Author(s) | Year | Title | Reason for exclusion |
|---|---|---|---|
| Ishizaki et al. | 2011 | Declines in physical performance by sex and age among nondisabled community-dwelling older Japanese during a 6-year period | 6 |
| Jang et al. | 2018 | Comparisons of predictive values of sarcopenia with different muscle mass indices in Korean rural older adults: a longitudinal analysis of the Aging Study of PyeongChang Rural Area | 6 |
| Jonkman et al. | 2019 | Development of a clinical prediction model for the onset of functional decline in people aged 65-75 years: pooled analysis of four European cohort studies | 1 |
| Keevil et al. | 2018 | Physical capability predicts mortality in late mid-life as well as in old age: Findings from a large British cohort study | 6 |
| Kim et al. | 2016 | Muscle strength: A better index of low physical performance than muscle mass in older adults | 5 |
| Kim et al. | 2013 | Prediction of severe, persistent activity-of-daily-living disability in older adults | 5 |
| Kim et al. | 2014 | Sarcopenia: an independent predictor of mortality in community-dwelling older Korean men | 6 |
| Kim et al. | 2020 | Longitudinal trajectory of disability in community-dwelling older adults: An observational cohort study in South Korea | 1 |
| Kita et al. | 2018 | Associations of the step-up test and lower limb dysfunction: A post-hoc analysis of a prospective cohort study | 6 |
| Kobayashi et al. | 2019 | Predictors of locomotive syndrome in community-living people: A prospective five-year longitudinal study | 6 |
| Kurichi et al. | 2017 | Predicting 3-year mortality and admission to acute-care hospitals, skilled nursing facilities, and long-term care facilities in Medicare beneficiaries | 5 |
| Kuroda et al. | 2018 | Risk factor for incident functional disability and the effect of a preventive exercise program: A 4-year prospective cohort study of older survivors from the great east Japan earthquake and nuclear disaster | 5 |
| Laddu et al. | 2018 | 36-Item Short Form Survey (SF-36) Versus Gait Speed As Predictor of Preclinical Mobility Disability in Older Women: The Women's Health Initiative | 6 |
| Lam et al. | 2020 | Cumulative and Incremental Value of Sarcopenia Components on Predicting Adverse Outcomes | 4 |
| Landi et al.. | 2016 | Impact of physical function impairment and multimorbidity on mortality among community-living older persons with sarcopaenia: results from the ilSIRENTE prospective cohort study | 6 |
| Laukkanen et al. | 2000 | Health and functional capacity as predictors of community dwelling among elderly people | 6 |
| Laukkanen et al. | 1995 | Muscle strength and mobility as predictors of survival in 75-84-year-old people | 6 |
| Lee et al. | 2005 | Association of cognitive status with functional limitation and disability in older adults | 5 |
| Legrand et al. | 2014 | Muscle strength and physical performance as predictors of mortality, hospitalization, and disability in the oldest old | 4 |
| Leinonen et al. | 2001 | Predictors of decline in self-assessments of health among older people - A 5-year longitudinal study | 6 |
| Leskinen et al. | 2013 | Prevalence, predictors and covariates of functional status impairment among Finnish Second World War veterans during 1992-2004 | 7 |
| Lin et al. | 2004 | Psychometric comparisons of the timed up and go, one-leg stand, functional reach, and Tinetti balance measures in community-dwelling older people | 4 |
| Lino et al. | 2019 | Association between visual problems, insufficient emotional support and urinary incontinence with disability in elderly people living in a poor district in Rio de Janeiro, Brazil: A six-year follow-up study | 6 |
| Lorquet et al. | 2019 | Three-year adverse health consequences of sarcopenia in community-dwelling older adults according to five diagnosis definitions | 6 |

| Author(s) | Year | Title | Reason for exclusion |
|---|---|---|---|
| Lv et al. | 2018 | Association of Body Mass Index With Disability in Activities of Daily Living Among Chinese Adults 80 Years of Age or Older | 5 |
| Lyons et al. | 2016 | Slow Gait Speed and Risk of Long-Term Nursing Home Residence in Older Women, Adjusting for Competing Risk of Mortality: Results from the Study of Osteoporotic Fractures | 7 |
| Manty et al. | 2007 | Construct and predictive validity of a self-reported measure of preclinical mobility limitation | 6 |
| Marsh et al. | 2011 | Muscle strength and BMI as predictors of major mobility disability in the Lifestyle Interventions and Independence for Elders pilot (LIFE-P) | 5 |
| Mathieson et al. | 2002 | Maintaining functional independence in elderly adults: the roles of health status and financial resources in predicting home modifications and use of mobility equipment | 6 |
| McAuley et al. | 2007 | Effects of change in physical activity on physical function limitations in older women: Mediating roles of physical function performance and self-efficacy | 6 |
| McDougall et al. | 2019 | Predictors of instrumental activities of daily living in community-dwelling older adults | 5 |
| Mello et al. | 2014 | Physical performance measures in the prediction of long-term outcomes after hospitalization in older patients: Preliminary results from the cape horn study | 9 |
| Melzer et al. | 2003 | The predictive validity for mortality of the index of mobility-related limitation--results from the EPESE study | 6 |
| Menant et al. | 2017 | Strength measures are better than muscle mass measures in predicting health-related outcomes in older people: time to abandon the term sarcopenia? | 5 |
| Mendes de Leon et al. | 2002 | Short-term change in physical function and disability: the Women's Health and Aging Study | 7 |
| Milani et al. | 2019 | Measures of lower body function as predictors of mortality among Mexican Americans aged 75 and older | 9 |
| Miller et al. | 2000 | Physical activity, functional limitations, and disability in older adults | 5 |
| Mitchell et al. | 2008 | Prediction of functional status in older adults: The ecological validity of four Delis-Kaplan Executive Function System tests | 5 |
| Montero-Odasso et al. | 2011 | Gait velocity versus the timed up and go test: Which one to use for the prediction of falls and other adverse health outcomes in primary care? | 9 |
| Mor et al. | 1989 | Risk of functional decline among well elders | 5 |
| Mossakowska et al. | 2014 | Cognitive performance and functional status are the major factors predicting survival of centenarians in Poland | 5 |
| Muller et al. | 2020 | Development and internal validation of prognostic models to predict negative health outcomes in older patients with multimorbidity and polypharmacy in general practice. | 5 |
| Munguia et al. | 2018 | Association of physical performance tests with frailty indicators and oxidative stress markers in a sample of a community-dwelling elderly population | 6 |
| Nagarkar et al. | 2017 | Predictors of functional disability with focus on activities of daily living: A community based follow-up study in older adults in India | 5 |
| Nam et al. | 2017 | A concordance of self-reported and performance-based assessments of mobility as a mortality predictor for older Mexican Americans | 5 |
| Nam et al. | 2016 | Lower body function as a predictor of mortality over 13 years of follow up: Findings from Hispanic Established Population for the Epidemiological Study of the Elderly | 7 |

| Author(s) | Year | Title | Reason for exclusion |
|---|---|---|---|
| Newman et al. | 2006 | Association of long-distance corridor walk performance with mortality, cardiovascular disease, mobility limitation, and disability | 6 |
| Ni et al. | 2017 | Clinically meaningful cutpoints of leg power in predicting disability, falls, and hospitalizations in older adults | 9 |
| Nikolaus et al. | 1996 | Prospective value of self-report and performance-based tests of functional status for 18-month outcomes in elderly patients | 2 |
| Oh-Park et al. | 2011 | Stair negotiation time in community-dwelling older adults: normative values and association with functional decline | 5 |
| Onder et al. | 2002 | Change in physical performance over time in older women: the Women's Health and Aging Study | 4 |
| Ostir et al. | 2000 | Emotional well-being predicts subsequent functional independence and survival | 5 |
| Panas et al. | 2013 | Physical performance and short-term mortality in very old Mexican Americans | 6 |
| Parker et al. | 1996 | Predictors of physical function among the oldest old: a comparison of three outcome variables in a 24-year follow-up | 5 |
| Pedone et al. | 2016 | Are Performance Measures Necessary to Predict Loss of Independence in Elderly People? | 5 |
| Peel et al. | 2013 | Gait speed as a measure in geriatric assessment in clinical settings: a systematic review | 1 |
| Penninx et al. | 2000 | Lower extremity performance in nondisabled older persons as a predictor of subsequent hospitalization | 6 |
| Perera et al. | 2016 | Gait Speed Predicts Incident Disability: A Pooled Analysis | 1 |
| Peres et al. | 2005 | The disablement process: factors associated with progression of disability and recovery in French elderly people | 5 |
| Perrault et al. | 2002 | Prognostic factors for functional independence in older adults with mild dementia: Results from the Canadian study of health and aging | 5 |
| Pinto et al. | 2017 | Global Sensory Impairment Predicts Morbidity and Mortality in Older U.S. Adults | 5 |
| Portegijs et al. | 2016 | Identification of Older People at Risk of ADL Disability Using the Life-Space Assessment: A Longitudinal Cohort Study | 5 |
| Proctor et al. | 2006 | Longitudinal changes in physical functional performance among the oldest old: insight from a study of Swedish twins | 1 |
| Pua et al. | 2019 | Physical Performance Predictor Measures in Older Adults With Falls-Related Emergency Department Visits | 7 |
| Rajan et al. | 2012 | Cognitive and physical functions as determinants of delayed age at onset and progression of disability | 7 |
| Rejeski et al. | 2015 | The MAT-sf: identifying risk for major mobility disability | 5 |
| Reuben et al. | 2004 | Refining the categorization of physical functional status: the added value of combining self-reported and performance-based measures | 4 |
| Reuben et al. | 1992 | The predictive validity of self-report and performance-based measures of function and health | 6 |
| Ribeiro et al. | 2016 | Fruit and Vegetable Intake and Physical Activity as Predictors of Disability Risk Factors in African-American Middle-Aged Individuals | 5 |
| Rivera et al. | 2008 | At the tipping point: predicting severe mobility difficulty in vulnerable older women | 4 |
| Rolland et al. | 2009 | [Sarcopenia] | 6 |
| Rolland et al. | 2006 | Physical performance measures as predictors of mortality in a cohort of community-dwelling older French women | 6 |

| Author(s) | Year | Title | Reason for exclusion |
|---|---|---|---|
| Rosano et al. | 2008 | Association between lower digit symbol substitution test score and slower gait and greater risk of mortality and of developing incident disability in well-functioning older adults | 7 |
| Rothman et al. | 2008 | Prognostic significance of potential frailty criteria | 7 |
| Russell et al. | 2015 | Predictors of long-term function in older community-dwelling people who have presented to an emergency department after a fall: a cohort study | 6 |
| Sanchez-Martinez et al. | 2016 | Transitions in functional status of community dwelling older adults: Impact of physical performance, depression and cognition | 4 |
| Santos-Eggimann et al. | 2008 | The Lausanne cohort Lc65+: a population-based prospective study of the manifestations, determinants and outcomes of frailty | 1 |
| Sarkisian et al. | 2001 | Correlates of attributing new disability to old age | 1 |
| Sarkisian et al. | 2000 | Modifiable risk factors predict functional decline among older women: a prospectively validated clinical prediction tool | 5 |
| Savino et al. | 2014 | Assessment of mobility status and risk of mobility disability in older persons | 1 |
| Scarlata et al. | 2008 | Restrictive pulmonary dysfunction at spirometry and mortality in the elderly | 6 |
| Schroll et al. | 1997 | Predictors of five-year functional ability in a longitudinal survey of men and women aged 75 to 80. The 1914-population in Glostrup, Denmark | 6 |
| Seeman et al. | 1995 | Behavioral and psychosocial predictors of physical performance: MacArthur studies of successful aging | 6 |
| Seidel et al. | 2011 | Limitations in physical functioning among older people as a predictor of subsequent disability in instrumental activities of daily living | 6 |
| Sharkey et al. | 2004 | Nutrient intake and BMI as predictors of severity of ADL disability over 1 year in homebound elders | 5 |
| Sheppard et al. | 2013 | Life-space mobility predicts nursing home admission over 6 years | 5 |
| Shi et al. | 2019 | Predicting mortality and disability: A comparison between the frailty index and general prognostic indices | 9 |
| Shimada et al. | 2010 | Predictive validity of the classification schema for functional mobility tests in instrumental activities of daily living decline among older adults | 6 |
| Shinkai et al. | 2000 | Walking speed as a good predictor for the onset of functional dependence in a Japanese rural community population | 6 |
| Simonsick et al. | 2016 | Fatigued, but Not Frail: Perceived Fatigability as a Marker of Impending Decline in Mobility-Intact Older Adults | 5 |
| Simonsick et al. | 2008 | Mobility limitation in self-described well-functioning older adults: importance of endurance walk testing | 6 |
| Sitjas Molina et al. | 2003 | Predictors factors about functional decline in community-dwelling older persons. [Spanish] | 8 |
| Siu et al. | 1993 | Using multidimensional health measures in older persons to identify risk of hospitalization and skilled nursing placement | 2 |
| Sonn et al. | 1996 | Activities of daily living studied longitudinally between 70 and 76 years of age | 5 |
| Srithumsuk et al. | 2020 | Association between physical function and long-term care in community-dwelling older and oldest people: the SONIC study | 7 |
| Strawbridge et al. | 1996 | Successful aging: predictors and associated activities | 5 |
| Sugiura et al. | 2013 | Handgrip strength as a predictor of higher-level competence decline among community-dwelling Japanese elderly in an urban area during a 4-year follow-up | 6 |

| Author(s) | Year | Title | Reason for exclusion |
|---|---|---|---|
| Sutorius et al. | 2016 | Comparison of 10 single and stepped methods to identify frail older persons in primary care: diagnostic and prognostic accuracy | 6 |
| Syddall et al. | 2015 | Self-Reported Walking Speed: A Useful Marker of Physical Performance Among Community-Dwelling Older People? | 6 |
| Szabo et al. | 2011 | Longitudinal invariance and construct validity of the abbreviated late-life function and disability instrument in healthy older adults | 5 |
| Taekema et al. | 2012 | Predicting survival in oldest old people | 6 |
| Taniguchi et al. | 2016 | Prospective Study of Trajectories of Physical Performance and Mortality Among Community-Dwelling Older Japanese | 6 |
| Terhorst et al. | 2016 | Performance-based impairment measures as predictors of activity limitations in community-dwelling older adults | 9 |
| Thakral et al. | 2014 | A stiff price to pay: does joint stiffness predict disability in an older population? | 5 |
| Tian et al. | 2017 | The relative temporal sequence of decline in mobility and cognition among initially unimpaired older adults: Results from the Baltimore longitudinal study of aging | 6 |
| Tinetti et al. | 2005 | Modifiable impairments predict progressive disability among older persons | 6 |
| Tseng et al. | 2012 | The functional performance predictors of adverse health outcomes in community-dwelling older adults: A preliminary study | 9 |
| Tsuji et al. | 2018 | The effect of body mass index, lower extremity performance, and use of a private car on incident life-space restriction: a two-year follow-up study | 6 |
| Tsutsumimoto et al. | 2018 | Aging-related anorexia and its association with disability and frailty | 5 |
| Uemura et al. | 2018 | The impact of sarcopenia on incident homebound status among community-dwelling older adults: A prospective cohort study | 5 |
| Vasunilashorn et al. | 2009 | Use of the Short Physical Performance Battery Score to predict loss of ability to walk 400 meters: analysis from the InCHIANTI study | 6 |
| Vaughan et al. | 2016 | Functional Independence in Late-Life: Maintaining Physical Functioning in Older Adulthood Predicts Daily Life Function after Age 80 | 7 |
| Veronese et al. | 2017 | A Comparison of Objective Physical Performance Tests and Future Mortality in the Elderly People | 6 |
| Verreckt et al. | 2017 | Which specific executive functions are predictors of functional decline in community-dwelling older adults? | 5 |
| Vestergaard et al. | 2009 | Characteristics of 400-meter walk test performance and subsequent mortality in older adults | 6 |
| Vestergaard et al. | 2009 | Stopping to rest during a 400-meter walk and incident mobility disability in older persons with functional limitations | 6 |
| Viccaro et al. | 2011 | Is timed up and go better than gait speed in predicting health, function, and falls in older adults? | 4 |
| Visser et al. | 2005 | Muscle mass, muscle strength, and muscle fat infiltration as predictors of incident mobility limitations in well-functioning older persons | 5 |
| Volpato et al. | 2016 | Short physical performance battery and all-cause mortality: A systematic review and meta-analysis | 1 |
| Von Bonsdorff et al. | 2013 | Coronary artery calcium and physical performance as determinants of mortality in older age: The AGES-Reykjavik Study | 6 |
| Wang et al. | 2002 | Predictors of functional change: A longitudinal study of nondemented people aged 65 and older | 5 |

| Author(s) | Year | Title | Reason for exclusion |
|---|---|---|---|
| Wang et al. | 2001 | Incidence of nursing home placement in a defined community | 5 |
| Wang et al. | 2011 | Mobility-related performance tests to predict mobility disability at 2-year follow-up in community-dwelling older adults | 6 |
| Ward et al. | 2016 | A Novel Approach to Identifying Trajectories of Mobility Change in Older Adults | 5 |
| Ward et al. | 2016 | Neuromuscular Impairments Contributing to Persistently Poor and Declining Lower-Extremity Mobility Among Older Adults: New Findings Informing Geriatric Rehabilitation | 5 |
| Weatherall et al. | 2004 | Risk factors for entry into residential care after a support-needs assessment | 5 |
| Wei et al. | 2019 | Physical Functioning Decline and Mortality in Older Adults With Multimorbidity: Joint Modeling of Longitudinal and Survival Data | 5 |
| Weiss et al. | 2012 | Incident preclinical mobility disability (PCMD) increases future risk of new difficulty walking and reduction in walking activity | 5 |
| Weiss et al. | 2007 | Exploring the hierarchy of mobility performance in high-functioning older women | 6 |
| Williams et al. | 1994 | The Timed Manual Performance test as a predictor of hospitalization and death in a community-based elderly population | 5 |
| Wolinsky et al. | 2007 | Four-year lower extremity disability trajectories among African American men and women | 3 |
| Woo et al. | 2012 | Comparison of frailty indicators based on clinical phenotype and the multiple deficit approach in predicting mortality and physical limitation | 5 |
| Woo et al. | 2018 | Predictive Ability of Individual Items of the Cardiovascular Health Study (CHS) Scale Compared With the Summative Score | 6 |
| Woods et al. | 2005 | Frailty: emergence and consequences in women aged 65 and older in the Women's Health Initiative Observational Study | 5 |
| Wu et al. | 1999 | Incidence of and predictors for chronic disability in activities of daily living among older people in Taiwan | 5 |
| Wu et al. | 2013 | The association between functional disability and acute care utilization among the elderly in Taiwan | 6 |
| Yamada et al. | 2015 | Predictive Value of Frailty Scores for Healthy Life Expectancy in Community-Dwelling Older Japanese Adults | 5 |
| Yeom et al. | 2008 | Risk Factors for Mobility Limitation in Community-Dwelling Older Adults: A Social Ecological Perspective | 1 |
| Yu et al. | 2019 | The relationship between self-reported sensory impairments and psychosocial health in older adults: a 4-year follow-up study using the English Longitudinal Study of Ageing | 5 |
| Zhang et al. | 2019 | Role of physical performance measures for identifying functional disability among Chinese older adults: Data from the China Health and Retirement longitudinal study | 1 |
| Zhang et al. | 2020 | Frailty as a predictor of future falls and disability: a four-year follow-up study of Chinese older adults | 5 |

Reasons for exclusion:
1 = No prospective longitudinal study design or no analysis of association between mobility and disability
2 = Study population: not community-dwelling
3 = Study population: mean age <60 years
4 = Study population: disability at baseline
5 = No relevant assessment of mobility capacity at baseline
6 = No relevant assessment of ADL disability at follow-up

7 = Inadequate follow-up period (<1 year or >6 years)
8 = No English or German full text available
9 = Abstract only, no full text available

### eTable 6: Reports identified through other sources and excluded after full-text assessment (n = 43)

| Author(s) | Year | Title | Reason for exclusion |
|---|---|---|---|
| Abizanda et al. | 2013 | Frailty and mortality, disability and mobility loss in a Spanish cohort of older adults: the FRADEA study | 4 |
| Aguero-Torres et al. | 2001 | Institutionalisation in the elderly: the role of chronic diseases and dementia. Cross-sectional and longitudinal data from a population-based study | 5 |
| Ailinger et al. | 1993 | Predictors of function among older Hispanic immigrants: a 5-year follow-up | 5 |
| Andel et al. | 2007 | Risk factors for nursing home placement in older adults with and without dementia | 5 |
| Branch et al. | 1984 | A prospective study of functional status among community elders | 5 |
| Branch et al. | 1982 | A prospective study of long-term care institutionalization among the aged | 5 |
| Clark et al. | 1997 | Prevalence and impact of risk factors for lower body difficulty among Mexican Americans, African Americans, and Whites | 5 |
| Cohen et al. | 1986 | Client-related risk factors of nursing home entry among elderly adults. | 5 |
| den Ouden et al. | 2013 | Association between physical performance characteristics and independence in activities of daily living in middle-aged and elderly men | 1 |
| Duchowny at al. | 2018 | Muscle Weakness and Physical Disability in Older Americans: Longitudinal Findings from the U.S. Health and Retirement Study | 5 |
| Fantin et al. | 2007 | Longitudinal body composition changes in old men and women: interrelationships with worsening disability | 4 |
| Gill et al. | 1996 | Impairments in physical performance and cognitive status as predisposing factors for functional dependence among nondisabled older persons | 5 |
| Gobbens et al. | 2014 | The prediction of ADL and IADL disability using six physical indicators of frailty: a longitudinal study in the Netherlands | 4 |
| Harris et al. | 1989 | Longitudinal study of physical ability in the oldest-old | 5 |
| Hirani et al. | 2015 | Sarcopenia is associated with incident disability, institutionalization, and mortality in community dwelling older men: the Concord Health and Ageing in Men Project | 1 |
| Ishizaki et al. | 2000 | Predictors for functional decline among nondisabled older Japanese living in a community during a 3-year follow-up | 5 |
| Jette et al. | 1990 | Musculoskeletal impairments and physical disablement among the aged | 5 |
| Jonkman et al. | 2018 | Predicting trajectories of functional decline in 60- to 70-year-old people. | 7 |
| Kaplan et al. | 1993 | Factors associated with change in physical functioning in the elderly: a six-year prospective study | 5 |
| Kempen et al. | 1998 | The impact of physical performance and cognitive status on subsequent ADL disability in low functioning older adults | 4 |
| Klein T et al. | 1994 | Determinants of nursing home admission of elderly patients and chances for prevention. A longitudinal study in Germany | 5 |
| LaCroix et al. | 1993 | Maintaining mobility in late life. II. Smoking, alcohol consumption, physical activity, and body mass index | 5 |
| Liu et al. | 1994 | Risk for entering nursing homes for long versus short stays | 5 |
| Lopez-Teros et al. | 2014 | Gait speed and handgrip strength as predictors of incident disability in Mexican older adults | 6 |
| Macklai et al. | 2013 | Prospective association of the SHARE-operationalized frailty phenotype with adverse health outcomes: evidence from 60þ community-dwelling Europeans living in 11 countries | 5 |

| Author(s) | Year | Title | Reason for exclusion |
|---|---|---|---|
| Mayhew et al. | 2020 | The association between self-reported and performance-based physical function with activities of daily living disability in the canadian longitudinal study on aging | 1 |
| McGrath et al. | 2018 | Muscle Strength and Functional Limitations: Preserving Function in Older Mexican Americans | 1 |
| Mustard et al. | 1999 | What determines the need for nursing home admission in a universally insured population? | 5 |
| Nygaard et al. | 1992 | Risk factors for admission to a nursing home: a study of elderly people receiving home nursing | 5 |
| Rantanen et al. | 2001 | Co-impairments as predictors of severe walking disability in older women | 4 |
| Shapiro et al. | 1985 | Predictors of long term care facility use among the elderly | 5 |
| Shimada et al. | 2015 | Incidence of disability in frail older persons with or without slow walking speed | 1 |
| Sonn et al. | 1995 | Instrumental activities of daily living related to impairments and functional limitations in 70-year-olds and changes between 70 and 76 years of age | 1 |
| Spiers et al. | 2005 | Diseases and impairments as risk factors for onset of disability in the older population in England and Wales: Findings from the Medical Research Council Cognitive Function and Ageing Study | 5 |
| Tanimoto et al. | 2013 | Association of sarcopenia with functional decline in community-dwelling elderly subjects in Japan | 1 |
| Tas et al. | 2007 | Incidence and risk factors of disability in the elderly: The Rotterdam Study | 5 |
| Terhorst et al. | 2017 | Performance-based impairment measures as predictors of early-stage activity limitations in community-dwelling older adults | 7 |
| Tinetti et al. | 1995 | Shared risk factors for falls, incontinence, and functional dependence: unifying the approach to geriatric syndromes | 4 |
| Wang et al. | 2001 | Incidence of nursing home placement in a defined community | 5 |
| Williams et al. | 1987 | Identifying the older person likely to require long-term care services | 5 |
| Wolinsky et al. | 1993 | Changes in functional status and the risks of subsequent nursing home placement and death | 5 |
| Woo et al. | 2000 | An estimate of long-term care needs and identification of risk factors for institutionalization among Hong Kong Chinese aged 70 years and over | 5 |

Reasons for exclusion:
1 = No prospective longitudinal study design or no analysis of association between mobility and disability
2 = Study population: not community-dwelling
3 = Study population: mean age <60 years
4 = Study population: disability at baseline
5 = No relevant assessment of mobility capacity at baseline
6 = No relevant assessment of ADL disability at follow-up
7 = Inadequate follow-up period (<1 year or >6 years)
8 = No English or German full text available
9 = Abstract only, no full text available

### eTable 7: Key characteristics of the included reports (n = 40)

| Author(s) | Year | Location (Country) | Parent study | Sample size at baseline | Sample size used for analysis (at follow-up) | Completion rate | Age in years; mean ± SD | Female (%) | Cognition (complete sample at baseline; MMSE mean ± SD) | Mobility assessment(s) | Disability outcome | Disability description/components | Disabled at follow-up (%) | Follow-up time | Type of analysis | Effect Measure(s) | Adjustment variables | NOS |
|---|---|---|---|---|---|---|---|---|---|---|---|---|---|---|---|---|---|---|
| Abe et al. | 2019 | Japan | KLSAH | 941 | 755 | 80% | 74.6 ± 5.5 | 59% | NR | Gait speed (usual) | Incident LTCI certification | Any new certification, support or care level ≥1. | 247 (32.9%) | 4.4 years (median) | Cox regression analysis and multiple regression analysis | aHR | A, G, B, C | 9 |
| Akune et al. | 2014 | Japan | ROAD study | 1773 | 1626 | 92% | 75.4 ± 5.2 | 61% | NR | Gait speed (usual), CRT | Incident LTCI certification | Any new certification, support or care level ≥1. | 169 (9.6%) | 4 years (mean) | Cox proportional hazards regression analysis | aHR | A, G, B, O | 9 |
| Artaud et al. | 2015 | France | 3C Study | 4313 | 3814 | 88% | 73.2 ± 4.6 | 61% | 27.4 ± 1.7 | Gait speed (fast) | Incident disability (hierarchical disability indicator including mobility, IADL and ADL) | Level 2 (IADL and mobility disability) and level 3 (level 2 plus ADL disability) | 628 (16.5%) | 5.1 ± 4.2 years (mean ± SD) | Logistic regression | aHR | A, G | 9 |
| Balzi et al. | 2010 | Italy | InCHIANTI | 1106 | 848 | 77% | 74.2 ± 6.9 | 56% | NR | SPPB | Incident ADL | Need for help of another person in ≥1 of 6 ADLs: bathing, dressing, eating, getting into and out of bed or chair, walking across a room, using the toilet | 72 (8.5%) | 3 years | Logistic regression | aOR | A, G, C, B, O | 8 |
| Beauchamp et al. | 2015 | USA | Boston RISE | 430 | 360 | 84% | 76.6 ± 7.0 | 68% | NR | SPPB, gait speed (usual), 400m-walk-test | LLFDI disability component (limitation and frequency domains) | NA | Mean LLFDI-DC score limitation: 68.9 ± 11.8; Mean LLFDI-DC score frequency: 52.3 ± 5.7 | 2 years | Linear regression models | amount of total variance ($R^2$) | NR | 4 |
| Bjorkman et al. | 2019 | Finland | PSNT | 428 | 262 | 61% | 82.7 ± 4.4 | 74% | 26.1 ± 2.7 | SPPB | Use of home care services | Regularly use of either municipal or private home care services | 69 (26.3%) | 4 years | Binary logistic regression analysis and linear regression analyses | aOR | A, G | 7 |
| Buchman et al. | 2021 | USA | MAP | 1026 | 1026 | 100% | 81.3 ± 7.3 | 76% | NR | Gait speed (usual) | Incident ADL | Need for assistance in ≥1 of 6 ADLs: feeding, bathing, dressing, toileting, transferring, and walking across a small room | 483 (47.1%) | 4.8 ± 3.2 years (mean ± SD) | Cox proportional hazards model | aHR | A, G, O | 8 |
| Chaudhry et al. | 2010 | USA | CHS | 5888 | 5646 | 96% | 65-69 y: 34%; 70-74 y: 32%; 75-79 y: 20%; ≥80 y: 14% | 58% | NR | Gait speed (usual) | Incident ADL | A lot of difficulty or being unable to do ≥1 of 6 ADLs: bathing, dressing, walking around the home, getting out of bed or a chair, eating, using the toilet | 869 (15.4%) | 6 years (median) | Multivariable Cox hazards regression modeling | aHR | A, G, C, B, O | 9 |
| Chu et al. | 2006 | China | Original study | 1517 | 1419 | 94% | 73.1 ± 6.2 | 50% | AMT 8.9 ± 1.1 | Gait speed (fast) | Decline in BI score over 1 year >1 SD of sample baseline value | NA | 72 (5.1%) decliners in BI score | 1 year | Multivariate logistic regression | aRR | A | 7 |

| **Author(s)** | **Year** | **Location (Country)** | **Parent study** | **Sample size at baseline** | **Sample size used for analysis (at follow-up)** | **Completion rate** | **Age in years; mean ± SD** | **Female (%)** | **Cognition (complete sample at baseline; MMSE mean ± SD)** | **Mobility assessment(s)** | **Disability outcome** | **Disability description/components** | **Disabled at follow-up (%)** | **Follow-up time** | **Type of analysis** | **Effect Measure(s)** | **Adjustment variables** | **NOS** |
|---|---|---|---|---|---|---|---|---|---|---|---|---|---|---|---|---|---|---|
| da Silva Alexandre et al. | 2012 | Brasil | SABE | 1634 | 910 | 56% | 68.6 ± 9.1 | 61% | 16.1 ± 2.8 | CRT, one-leg standing time | Incident ADL | Difficulty to perform ≥1 of 6 ADLs: walking, transferring, toileting, bathing, dressing or feeding | M: 69 (19.4%)<br>F: 181 (32.6%)<br>Total: 250 (27.5%) | 6 years | Logistic regression analysis | aOR | G, C | 8 |
| DeVore et al. | 1994 | USA | Original study | 124 | 124 | 100% | 76.0 | 78% | NR | POMA | Change in living situation: Nursing home admission or joining relative's households | NA | 13 (10.5%) | 1.5 years | C-statistics | C-statistics | NR | 3 |
| Diem et al. | 2018 | USA | SOF | 1034 | 1010 | 98% | 88.0 ± 2.6 | 100% | mMMSE <88 points: 33% | Gait speed (usual) | Incident ADL | Living in a nursing home or unable to perform ≥2 of 5 ADLs independently: eating, dressing, bathing, transferring from a bed or chair, using the toilet | 208 (20.6%) | 5 years | Multivariable logistic regression | aRR | A, C, O | 7 |
| Doi et al. | 2020 | Japan | OSHPE | 4121 | 4121 | 100% | 71.9 ± 5.0 | 53% | 26.4 ± 2.4 | Gait speed (usual) | Incident LTCI certification | Any new certification of LTCI at any level | 425 (10.3%) | 49.6 months = 4.1 years (mean) | Cox proportional hazard model; c-statistics | aHR; AUC | A, G, C, M, B, O | 9 |
| Donoghue et al. | 2014 | Ireland | TILDA | 1879 | 1664 | 89% | 72.8 ± 6.1 | 53% | 28.0 ± 1.7 | Gait speed (usual), TUG | Incident ADL | Difficulty in ≥1 of 6 ADLs: dressing, walking across a room, bathing or showering, eating, getting in or out of bed, using the toilet | 80 (4.9%) | 2 years | ROC analyses, logistic regression | AUC, probability | NR | 6 |
| Gill et al. | 2004 | USA | PEP | 754 | 754 | 100% | 78.4 ± 5.3 | 65% | 26.8 ± 2.5 | Gait speed (fast) | Incident ADL | Need for personal assistance in ≥1 of 4 ADLs: bathing, dressing, walking inside the house, transferring from a chair | 417 (55.3%) | 5 years | Kaplan-Meier method; time-dependent Cox proportional hazards method | KMC; aHR | A, G, C, M, O | 8 |
| Gill et al. (J Gerontol A Biol Sci Med Sci) | 1995 | USA | PSC | 237 | 197 | 83% | 79.7 ± 5.7 | 70% | NR | POMA (total score & each item), gait speed (fast), CRT, 360° turn, 10 foot taps, time to bend over and pick up a pen | Incident ADL | Receiving personal assistance or being completely dependent in ≥1 of 7 ADLs: bathing, dressing, transferring, walking, eating, toileting, grooming | 31 (15.7%) | 1 year | NR | cRR | NR | 4 |
| Gill et al. (JAGS) | 1995 | USA | PSC | 664 | 563 | 85% | 79.1 ± 4.7 | 74% | 27.0 ± 1.7 | Gait speed (fast), CRT, 360° turn, 10 foot taps, time to bend over and pick up a pen | Incident ADL | Receiving personal assistance or being completely dependent in ≥1 of 7 ADLs: bathing, dressing, transferring, walking, eating, toileting, grooming | 53 (9.4%) | 1 year | Binomial regression models | aRR | A, G, O | 7 |
| Guralnik et al. | 2000 | USA | EPESE | 4588 | 3414 | 74% | NR | NR | NR | Gait speed (usual), EPESE lower extremity performance battery (SPPB) | Incident ADL | Having mobility-related disability plus the inability to perform ≥1 of 4 ADLs without help from another person: moving from a bed to a chair, using the toilet, bathing, walking across a small room | Iowa (1 year): 34/1342=2.5%;<br>New Haven (1 year): 17/655=2.6%;<br>Iowa (4 years): 123/1121=11.0%; | 1 to 6 years | Multiple logistic models | aRR | A, G, C | 8 |

| Author(s) | Year | Location (Country) | Parent study | Sample size at baseline | Sample size used for analysis (at follow-up) | Completion rate | Age in years; mean ± SD | Female (%) | Cognition (complete sample at baseline; MMSE mean ± SD) | Mobility assessment(s) | Disability outcome | Disability description/components | Disabled at follow-up (%) | Follow-up time | Type of analysis | Effect Measure(s) | Adjustment variables | NOS |
|---|---|---|---|---|---|---|---|---|---|---|---|---|---|---|---|---|---|---|
| | | | | | | | | | | | | | North Carolina (4 years): 100/962=10.4; New Haven (6 years): 75/455=16.5% | | | | | |
| Guralnik et al. | 1995 | USA | EPESE | 1363 | 1122 | 82% | 77.1 | 65% | NR | SPPB | Incident ADL | Inability to perform ≥1 of 4 ADLs without the help of another person (moving from a bed to a chair, using the toilet, bathing, and walking across a small room) and mobility-related disability (inability to walk a half mile or climb stairs without help) | 112 (10.0%) | 4 years | Multiple logistic-regression analysis | aRR | A, G, C | 8 |
| Heiland et al. | 2016 | Sweden | SNAC | 3060 | 1971 | 64% | 73.7 ± 10.8 | 64% | 27.9 ± 4.2 | Gait speed (self-selected), one-leg standing time | Incident ADL | Dependence in ≥1 of 5 ADLs: bathing, dressing, toileting, transferring in and out of bed and from bed to chair, eating | 119 (6.0%) | 5.8 ± 0.3 years (mean ± SD) | Binary logistic regression | aOR | A, G, C, M, O | 8 |
| Hoshi et al. | 2012 | Japan | Tsurugaya Project | 813 | 813 | 100% | 75.6 ± 4.4 | 51% | 28.2 ± 2.2 | Gait speed (max), TUG, FRT | Incident LTCI certification | Any new certification, support or care level ≥1. | 135 (16.6%) | 4 years | Cox proportional hazards regression analysis | aHR | A, G | 9 |
| Huang et al. | 2010 | USA | Original study | 110 | 75 | 68% | 80.3 ± 6.9 | 71% | 28.0 ± 1.9 | Gait speed (usual), SPPB, BBS, TUG | Incident ADL | Difficulty in ≥1 of 7 ADLs: bathing, dressing, eating, getting in/out of bed/chairs, personal hygiene, walking, using the toilet | 24 (32.0%) | 1,5 years | Logistic regression models; c-statistics | aOR, AUC | A, G, C | 5 |
| Lee et al. | 2020 | Korea | Original study | 25031 | 25031 | 100% | 66.0 ± 0.0 | 53% | Prescreening Korean Dementia Screening Questionnaire ≥4/10 points (impaired cognition): 22% | TUG | Incident LTCI certification | Any new certification, support or care level ≥1. | 331 (1.3%) | 5.7 years (mean) | Cox proportional hazard, KMC | aHR, KMC | G, C, M, O | 9 |
| Makizako et al. | 2017 | Japan | OSHPE | 4390 | 4335 | 99% | 71.7 ± 5.4 | 52% | 26.4 ± 2.6 | CRT, TUG | Incident LTCI certification | Any new certification, support or care level ≥1. | 161 (3.7%) | 2 years | Cox proportional hazards regression models; Kaplan-Meier curves | aHR, KMC | A, G, C, M, O | 9 |
| Makizako et al. | 2015 | Japan | OSHPE | 4396 | 4341 | 99% | 71.8 ± 5.4 | 52% | 26.4 ± 2.6 | Gait speed (usual) | Incident LTCI certification | Any new certification, support or care level ≥1. | 168 (3.9%) | 2 years | Cox proportional hazard, KMC | aHR, KMC | A, G, C, M, B, O | 9 |
| Minneci et al. | 2015 | Italy | ICARe | 561 | 453 | 81% | 72.9 ± 6.4 | 58% | 26,9 ± 2.1 | Gait speed (usual), SPPB, 6-minute walk test | Incident ADL | Complete inability or need for help in ≥1 of 6 ADLs: washing hands and face, dressing and undressing self, toileting, transferring from bed to chair, maintaining continence, eating | 33 (7.3%) | 3 years | Logistic regression models | aOR | A, G, C, M, O | 8 |

| Author(s) | Year | Location (Country) | Parent study | Sample size at baseline | Sample size used for analysis (at follow-up) | Completion rate | Age in years; mean ± SD | Female (%) | Cognition (complete sample at baseline; MMSE mean ± SD) | Mobility assessment(s) | Disability outcome | Disability description/components | Disabled at follow-up (%) | Follow-up time | Type of analysis | Effect Measure(s) | Adjustment variables | NOS |
|---|---|---|---|---|---|---|---|---|---|---|---|---|---|---|---|---|---|---|
| Montero-Odasso et al. | 2005 | Argentina | EFA | 102 | 101 | 99% | 79,6 ± 4.0 | 71% | 27.6 | Gait speed (usual), TUG, POMA | Need/requirement for a caregiver; nursing home placement | NA | Need/requirement for a caregiver: 15 (14.9%); nursing home placement: 3 (3.0%) | 2 years | Multiple logistic regression | aRR | A, G, B, M, O | 9 |
| Moriya et al. | 2013 | Japan | Original study | 882 | 784 | 89% | 72.9 ± 5.1 | 58% | NR | one-leg standing time with eyes open (OLST) | Incident LTCI certification | Any new certification, support or care level ≥1. | 94 (12.0%) | 5 years | Cox proportional hazard analysis | aHR | A, C | 9 |
| Onder et al. | 2005 | USA | WHAS-I | 458 | 454 | 99% | 78.7 ± 8.0 | 100% | 26.5 ± 3.0 | Gait speed (usual), CRT, Balance test | Incident ADL (progressive and catastrophic) | A lot of difficulty or inability to perform ≥1 of 5 ADLs: bathing, dressing, eating, transferring from the bed to a chair, using the toilet | Total: 221 (48.3%); progressive: 149 (32.5%); catastrophic: 72 (15.7%) | 3 years | Cox proportional hazard models; c-statistics | aHR, AUC | A, O | 7 |
| Ostir et al. | 1998 | USA | HEPESE | 1946 | 1365 | 70% | 73.3 | 53% | NR | Gait speed (usual), CRT, standing balance, Lower body function score (mSPPB) | Incident ADL | Self-reported disability in ≥1 of 4 ADLs: bathing, using the toilet, transferring from bed to chair, walking across a small room | 58 (4.2%) | 2 years | Multiple logistic regression analysis | aOR | A, G, C | 7 |
| Rosenberg et al. | 2019 | Canada | FACTS | 380 | 380 | 100% | 88.4 ± 6.5 | 72% | MoCA: 20.6 ± 6.4 | Gait speed (usual) | Nursing home transfer | NA | 48 (12.6%) | 1.5 years | Cox proportional hazard, KMC | aHR, KMC | A, G, C, O | 7 |
| Sakamoto et al. | 2016 | Japan | TLSA | 188 | 172 | 91% | 80.2 ± 3.9 | 65% | 26.1 ± 3.6 | TUG, FRT | Incident ADL | Dependence in ≥1 of 7 ADLs: walking, ascending and descending stairs, feeding, dressing, toileting, bathing, grooming | 38 (22.1%) | 2 years | Logistic regression analysis | aOR | A, G, O | 8 |
| Shinkai et al. | 2003 | Japan | TMIG-LISA | 601 | 601 | 100% | 70.9 ± 4.9 | 56% | NR | Gait speed (usual), gait speed (fast), one leg stance | Incidence ADL | Needing help from someone else or being unable to perform ≥1 of 5 ADLs (bathing, dressing, walking, eating, continence) or institutionalization or death | 194 (23.3%) | 6 years | Cox proportional hazard | aHR | A, G, C | 8 |
| Stenholm et al. | 2014 | Italy | InCHIANTI | 901 | 819 (3-year follow-up); 727 (6-year follow-up) | 91% (3 y); 81% (6 y) | 66.7 ± 14.1 | 54% | 26.8 ± 3.0 | Gait speed (usual), SPPB | Incident ADL | Mobility disability (inability to walk 400 m or climb and descend stairs independently) and inability to perform ≥1 of 6 ADLs independently: walking across small room, bathing or showering, dressing and undressing, eating meals by oneself, using the toilet, getting in and out of bed | 74/727 (10.2%; 6-year follow-up); 43/819 (5.3%; 3-year follow-up) | 3 years; 6 years | Logistic regression analysis | aOR | A, G, C, M, B, O | 8 |
| Studenski et al. | 2003 | USA | Original Study | 487 | 453 | 93% | 74.1 ± 5.7 | 44% | 27.5 ± 2.3 | Gait speed (usual), EPESE lower extremity performance battery (SPPB) | Incident ADL | Difficulty in ≥1 of 6 ADLs: eat, dress, bathe, toilet, groom, transfer | 122 (27.4%) | 1 year | Logistic regression models | aOR | A, O | 5 |
| Tsutsumimoto et al. | 2016 | Japan | OSHPE | 4038 | 4038 | 100% | 71.9 ± 2.5 | 51% | 26.4 ± 2.5 | Gait speed (usual) | Incident LTCI certification | Any new certification, support or care level ≥1. | 220 (5.4%) | 31 months = 2.6 | Cox proportional hazard | aHR | A, G, C, M, O | 9 |

| Author(s) | Year | Location (Country) | Parent study | Sample size at baseline | Sample size used for analysis (at follow-up) | Completion rate | Age in years; mean ± SD | Female (%) | Cognition (complete sample at baseline; MMSE mean ± SD) | Mobility assessment(s) | Disability outcome | Disability description/components | Disabled at follow-up (%) | Follow-up time | Type of analysis | Effect Measure(s) | Adjustment variables | NOS |
|---|---|---|---|---|---|---|---|---|---|---|---|---|---|---|---|---|---|---|
| | | | | | | | | | | | | | | years (median) | regression model | | | |
| Vaarst et al. | 2021 | Denmark | HANC | 323 | 323 | 100% | 81.7 ± 4.2 | 59% | 28.5 (95% CI: 28.3 – 28.7) | SPPB | Incident ADL (personal care) | Need for assistance (personal care) with ADLs such as bathing and dressing | 123 (38.1%) | 4.1 years (mean) | Cox proportional hazard model | aHR | G, B, O | 8 |
| Verghese et al. | 2012 | USA | EAS | 631 | 594 | 94% | 79.9 ± 5.3 | 61% | Blessed test; 1.85 ± 1.96 | Walking While Talking test, gait speed (usual), SPPB | Incident ADL | Needing assistance or inability to perform ≥1 of 7 ADLs: bathing, walking inside home, chair rise, dressing, feeding, toileting, grooming | 88 (14.8%) | 32 ± 18 months = 2.7 ± 1.5 years (mean ± SD) | Cox proportional hazard model | aHR | A, G, C, O | 8 |
| Woo et al. | 1999 | Hong Kong/China | Original study | 1835 | 1171 | 64% | 70 – 79y: 61%; ≥80y: 39% | 49% | NR | Gait speed (usual), stride lengths | Dependency (Barthel Index <20 points); institutionalization | NA | Dependency: 327 (27.9%); institutionalization: 45 (3.8%) | 3 years | Logistic regression analysis | aOR | A | 5 |
| Zhang et al. | 2013 | Italy | InCHIANTI | 562 | 562 | 100% | 71.4 ± 5.7 | 48% | NR | CRT | Incident ADL | Need for assistance in ≥1 of 6 ADLs: bathing, dressing, eating, getting into and out of bed or chair, walking across a room, using the toilet | 15 (2.7%) | 3 years | Logistic regression | aOR | A, O | 7 |

**Abbreviations:**

ADL = activities of daily living; aHR = adjusted Hazard Ratio; AMT = Abbreviated Mental Test; aOR = adjusted Odds Ratio; aRR = adjusted Relative Risk; AUC = area under the curve; BBS = Berg Balance Scale; BI = Barthel Index; Boston RISE = Boston Rehabilitative Impairment Study of the Elderly; CHS = Cardiovascular Health Study; CRT = Chair Rise Test; EAS = Einstein Aging Study; EFA = Estudio de Evaluacion Funcional del Anciano study; EPESE = Established Populations for the Epidemiologic Study of the Elderly; FRT = Functional Reach Test; HANC = Healthy Ageing Network of Competence; HEPESE = Hispanic EPESE; IADL = instrumental activities of daily living; ICARe = Insufficienza Cardiaca negli Anziani Residenti a Dicomano Study; InCHIANTI = Aging in the Chianti area; KLSAH = Kusatsu Longitudinal Study on Aging and Health; KMC = Kaplan-Meier curves; LLFDI = Late Life Function and Disability Instrument; LLFDI-DC = Late Life Function and Disability Instrument – disability component; LTCI = Long term care insurance; MAP = Rush Memory and Aging Project; mMMSE = modified Mini-mental state examination; MMSE = Mini-mental state examination; MoCA = Montreal Cognitive Assessment; mSPPB = modified Short Physical Performance Battery; NA = not applicable; NOS = Newcastle-Ottawa scale; NR = not reported; OLST = one-leg standing time with eyes open; OSHPE = Obu Study of Health Promotion for the Elderly; PEP = Precipitating Events Project; POMA = Performance-Oriented Mobility Assessment; PSC = Project Safety Cohort; PSNT = Porvoo Sarcopenia and Nutrition Trial; ROAD = Research on Osteoarthritis/Osteoporosis Against Disability; ROC = receiver operating characteristic; SABE = “Salud Bienestar y Envejecimiento en America Latina y el Caribe / Saude, Bem-Estar e Envelhecimento [Health, Wellbeing and Aging]; SD = standard deviation; SNAC = Swedish National study on Aging and Care; SOF = Study of Osteoporotic Fractures; SPPB = Short Physical Performance Battery; TILDA = Irish Longitudinal Study on Aging; TLSA = Tosa Longitudinal Aging Study; TMIG-LISA = Tokyo Metropolitan Institute of Gerontology Longitudinal Interdisciplinary Study on Aging; TUG = Timed Up and Go test; WHAS-I = Women’s Health and Aging Study I

**Abbreviations for adjustment variables:**

A = age; G = gender; C = comorbidity; M = mental/cognition; B = BMI; O = other

### eTable 8: Additional characteristics of the included reports (n = 40)

| Authors | Study | Sample/region | Study: inclusion criteria | Study: exclusion criteria | Report: inclusion criteria | Report: exclusion criteria | Minimum age | Gender (M/F/B) | Cognition excluded* | Start date | End date | Loss to follow-up/excluded; n (%) | Description of loss to follow-up |
|---|---|---|---|---|---|---|---|---|---|---|---|---|---|
| Abe et al. 2019 | Kusatsu Longitudinal Study on Aging and Health | Data from 2003 and 2013 waves (present baseline); Kusatsu Town, Gunma Prefecture, Japan | Residnets of Kusatsu town, Gunma prefecture, or Yoita town, Niigata prefecture, Japan;<br>Aged ≥70 years;<br>complete baseline assessments;<br>cognitively intact at baseline (MMSE score > 24/30);<br>be re-evaluated using the MMSE at least once during the follow-up period | None | Age ≥70 (2003 sample) and ≥65 (2013 sample) years | Inability to walk without support at baseline;<br>already LCTI certified | ≥65 | B | Y | 2003 | Sec 2017 | 186 (20%) | Missing data (181);<br>5 Outliners (5) |
| Akune et al. 2014 | ROAD study | Residents of any one of three communities: an urban region in Itabashi, Tokyo; a mountainous region in Hidakagawa, Wakayama; and a coastal region in Taiji, Wakayama, Japan | Community-dwelling;<br>ability to walk to the survey site;<br>ability to report data;<br>ability to understand and sign an informed consent form;<br>age: urban region ≥60 years, mountainous region ≥40 years, coastal region ≥40 years | None | Aged ≥65 years;<br>not certified as need of care level elderly in the national LTCI system at baseline | None | ≥65 | B | N | 2005 | 2010 | 147 (8%) | Information on LTCI certification missing (13);<br>died (126);<br>moved away (8) |
| Artaud et al. 2015 | 3C Study | French community-dwelling older adults from the Dijon center of the 3C Study | Living in one of the 3 cities or their suburbs and registered on the electoral rolls (Bordeaux, Dijon, Montpellier);<br>aged ≥65 years;<br>not institutionalized | aged <65 years;<br>refused baseline medical interview | Aged <85 years | Disabled at baseline;<br>Participants with missing data for any of the covariates;<br>Participants with disability status unknown at all waves, participants for whom fast gait speed was never measured during follow-up | ≥65 | B | N | 1999 | 2012 | 499 (12%) | Missing data for covariates (66);<br>disability status unknown (25);<br>missing gait speed measures at all waves (394);<br>missing measures before they became disabled (60) |
| Balzi et al. 2010 | InCHIANTI | Random sample from two Italian towns located in the Chianti geographic area, located outside the urban area of Florence, Italy;<br>baseline data of 1998 wave. | Aged ≥65 years | None | Free of disability at baseline | None | ≥65 | B | N | 1998 | 2003 | 258 (23.3%) | Died (125);<br>lost to follow-up (133) |
| Beauchamp et al. 2015 | Boston RISE | Older adults were recruited through nine primary care practices located across the greater Boston, MA, USA, area.<br>. | age ≥65 years;<br>ability to speak and understand English;<br>difficulty or task modification with walking ½ mile and/or climbing one flight of stairs | significant visual impairment;<br>uncontrolled hypertension;<br>lower extremity amputation;<br>supplemental oxygen use;<br>presence of a terminal disease;<br>major surgery or myocardial infarction in past 6 months;<br>planned major surgery;<br>planned move from Boston area within 2 years;<br>major medical problems interfering with safe and successful testing;<br>MMSE score <18; | None. | None. | ≥65 | B | Y | Dec 2009 | NR | 70 (16%) | No reason reported. |

| Authors | Study | Sample/region | Study: inclusion criteria | Study: exclusion criteria | Report: inclusion criteria | Report: exclusion criteria | Minimum age | Gender (M/F/B) | Cognition excluded* | Start date | End date | Loss to follow-up/excluded; n (%) | Description of loss to follow-up |
|---|---|---|---|---|---|---|---|---|---|---|---|---|---|
| | | | | SPPB score <4 | | | | | | | | | |
| Bjorkman et al. 2019 | Porvoo Sarcopenia and Nutrition Trial | Individuals screened for or included in RCT, including community dwelling older people with sarcopenia living in Porvoo, Finland. | Not relevant, since the present study (n=428) includes individuals which have been included in the RCT (parent study, n=182) and individuals which have been screened for RCT participation (n=246) | NA | Living in Porvoo, Finland;<br>community-dwelling;<br>aged ≥75 years;<br>at risk of sarcopenia (limitations in ADLs, sedentary lifestyle, falls, exhaustion, old age, low BMI) | None | ≥75 | B | N | NR | NR | 166 (39%) | Died (88);<br>non-respondents (78) |
| Buchman et al. 2021 | Rush Memory and Aging Project (MAP) | Cohort study: longitudinal clinical-pathologic investigation of chronic conditions of old age, Chicago area, USA | lay persons from across north-eastern Illinois;<br>written informed consent;<br>anatomical Gift Act for organ donation at the time of death; | None | Valid bioimpendance and grip strength testing | missing longitudinal outcomes;<br>missing one or more demographic covariates | NR | B | N | 2005 | NR | 0 | NA |
| Chaudhry et al. 2010 | CHS | Four communities across the United States (Sacramento County, CA; Washington County, MD; Forsyth County, NC; Allegheny County, PA), with additional sample of African Americans (minority representation) | ≥65 years;<br>community-dwelling;<br>noninstitutionalized;<br>expected to remain in the area for the next three years;<br>able to give informed consent;<br>did not require a proxy respondent at baseline | wheelchair-bound in the home at baseline;<br>receiving hospice treatment, radiation therapy or chemotherapy for cancer | 1989 CHS baseline sample, incorporating the African-American cohort beginning in 1992 | None | ≥65 | B | N | 1989 (1992 African-American cohort) | 1996 (1999 African - American cohort) | 242 (4%) | Disabled at baseline (242) |
| Chu et al. 2006 | Hong Kong study by Chu et al. 2005 | Hong Kong / China | Being Chinese;<br>age ≥65 years old;<br>living at home;<br>could walk independently or with a walking aid;<br>informed consent | Being non-Chinese;<br>age <65 years old;<br>non-ambulatory;<br>unable to cooperate in the assessment | None | None | ≥65 | B | N | Mar 1998 | NR | 98 (6.5%) | NR |
| da Silva Alexandre et al. 2012 | SABE study | Probabilistic sub-sample representative of the urban population aged 60 years or more; residents of the city of Sao Paulo. | Aged 60 and over;<br>residing in one of 7 main urban centres of seven countries in Latin America and the Caribbean;<br>statement of informed consent | None | None | Difficulties in walking, transferring, toileting, bathing, dressing or feeding according to the modified Katz Index | ≥60 | B | N | 2000 | 2006 | 724 (44%) | 402 had died (25%);<br>322 (20%) were either not located, had moved to another city, had been institutionalized or refused to participate |
| DeVore et al. 1994 | Original study | Community-dwelling private patients seen in a medical practice in Hyattsville, Maryland, USA | NA | NA | ≥65 years;<br>Entering the private medical practice between 10 Feb. 1990 and 20 Dec. 1991 | None | ≥65 | B | N | Feb 1990 | NR | 0 | NA |
| Diem et al. 2018 | Study of Osteoporotic Fractures (SOF) | Participants of SOF Year 20 (Y20) examination (present baseline);<br>from population-based listings of in areas of the US (Baltimore County, MD; Minneapolis, MN; Portland, OR; Monongahela Valley, PA);<br>additional sample of African Americans | Ambulatory;<br>Female;<br>without;<br>≥65 years old | bilateral hip replacement | at least minimal information collected at Y20 visit;<br>completed examination (battery of lower extremity physical performance and neuropsychological tests);<br>community-dwelling;<br>age 85-99 years;<br>able to perform all basic ADLs at the Y20 visit;<br>5-year follow-up status available | bilateral hip replacement | ≥85 | F | N | 2006 (Y20) | NR | 24 (2%) | No reason reported. |
| Doi et al. 2020 | Obu Study of Health Promotion for the Elderly | Population-based study in a community setting, Obu city, Japan | ≥65 years;<br>reside in Obu city, Japan | History of Parkinson disease or stroke;<br>Mini-Mental State scores less than 18;<br>Participation in other similar studies;<br>hospitalized or in residential care;<br>certified at levels 3–5 to require support or care by the Long-Term Care Insurance (LTCI) | None | Any dependency for basic ADL;<br>being certified at any level by LTCI;<br>having specific medical conditions (stroke, Parkinson's disease, Alzheimer's disease);<br>having severe cognitive impairment as assessed by a Mini-Mental State | ≥65 | B | Y | 2011 | NR | 0 | NA |

| Authors | Study | Sample/region | Study: inclusion criteria | Study: exclusion criteria | Report: inclusion criteria | Report: exclusion criteria | Minimum age | Gender (M/F/B) | Cognition excluded* | Start date | End date | Loss to follow-up/excluded; n (%) | Description of loss to follow-up |
|---|---|---|---|---|---|---|---|---|---|---|---|---|---|
| | | | | | | Examination (MMSE) score < 20; censoring due to moving away or death; having missing values for any of these variables | | | | | | | |
| Donoghue et al. 2014 | TILDA | Ireland nationwide (Irish Geodirectory) | Community-dwelling; ≥50 years | nursing home residents and those resident in other institutions; cognitive impairment or dementia | TILDA first wave data; age ≥65 years; completion of either UGS or TUG tests in the health assessment; no reported difficulty in either ADL or IADL at baseline; Mini Mental State Examination (MMSE) score ≥24. | None | ≥65 | B | Y | Jan 2009 | Dec 2012 | 155 (11%) | Death (27); refusal/withdrawal (114); no contact (7); other reasons e.g. emigration (7);<br><br>Gait speed analysis: exceptionally slow gait-speed of <0.55 m/s (5); TUG analysis: very slow performance of >20.5 s (10) |
| Gill et al. 1995 (J Gerontol A Biol Sci Med Sci) | Project Safety cohort | Residents of New Haven, Connecticut | Community-living; age ≥72 years; ability to speak English, Spanish, or Italian; ability to follow simple commands; ability to walk across a room without human assistance | None | Independent (requiring no personal assistance) in seven basic ADLs: bathing, dressing, transferring, walking, eating, toileting, and grooming; mild to moderate cognitive impairment (MMSE 16-23 points) | Severe cognitive Impairment (MMSE <16 points). | ≥72 | B | Y | 1989 | NR | 40 (17%) | Deceased (14); incomplete data (26) |
| Gill et al. 1995 (JAGS) | Project Safety cohort | New Haven, Connecticut, USA | Ambulatory; speak English, Spanish, or Italian; able to follow simple commands; noninstitutionalized; aged ≥72 years; living in New Haven, Connecticut | None | Independent in seven basic ADLs; ≥24 points | None | ≥72 | B | Y | Oct 1989 | NR | 101 (15%) | Deceased (27); incomplete data at the 1-year follow-up interview (74) |
| Gill et al. 2004 | PEP | People living in the area of New Haven, Connecticut, USA | community-dwelling; aged ≥70 years; free of disability in 4 essential ADLs (bathing, dressing, walking inside the house, transferring from a chair) | Significant cognitive impairment with no available proxy; inability to speak English; diagnosis of a terminal illness with a life expectancy of less than 12 months; a plan to move out of the New Haven, Conn, area during the next 12 months. | None | None | ≥70 | B | Y | Mar 1998 | Mar 2003 | 0 | NA |
| Guralnik et al. 1995 | EPESE | Sixth annual follow-up interview data (1988) as baseline; Rural area of Iowa and Washington counties, Iowa, USA | NR | NR | ≥71 years of age; living in the community | living in institutions; unable to participate in the interview because of cognitive or physical impairment; self-reported disability in the ADLs; self-reported mobility-related disability | ≥71 | B | Y | 1988 | NR | 241 (18%) | Deceased (208); reason unknown (33) |

| Authors | Study | Sample/region | Study: inclusion criteria | Study: exclusion criteria | Report: inclusion criteria | Report: exclusion criteria | Minimum age | Gender (M/F/B) | Cognition excluded* | Start date | End date | Loss to follow-up/excluded; n (%) | Description of loss to follow-up |
|---|---|---|---|---|---|---|---|---|---|---|---|---|---|
| | | | | | | (defined as the inability to walk a half mile [0.8 km] or climb stairs without assistance); unable to complete the performance tests; missing data on disability or performance measures | | | | | | | |
| Guralnik et al. 2000 | EPESE | Participant from the Boston, Iowa, New Haven or North Carolina EPESE samples; 6-year wave (present baseline) | People living in East Boston, Massachusetts; two rural counties in Iowa; New Haven, Connecticut; and segments of five counties in the north-central Piedmont area of North Carolina; age ≥65 years | None | Participants from EPESE 6-year wave | Deceased at EPESE follow-up 6 date; living in institutions; living at home but needing a proxy respondent because of cognitive or physical impairment; living outside the area and requiring a telephone interview; participants who refused the performance tests; persons reporting ADL disability or mobility-related disability at baseline; participants reporting no disability but having scores of 0 to 3 on the summary performance scale | ≥65 | B | Y | NR | NR | 1174 (26%) | Death and loss to follow-up (1174) |
| Heiland et al. 2016 | SNAC | SNAC-K (Kungsholmen area, central Stockholm, Sweden) sample of the SNAC study. Older adults of high socioeconomic status. | People living in the study areas (Skåne, Blekinge, Kungsholmen, Nordanstig); age ≥ 60 years; participants living in their homes or in institutions | None | SNAC-Kungsholmen participants | Persons with missing data on ADL and mobility tests at baseline | ≥60 | B | N | Mar 2001 | 2010 | 1089 (35.6%) | Persons with dependency in one or more ADL activities at baseline (152); deceased (523); did not participate in further follow-ups due to refusal or loss of contact (405); missing follow-up ADL data (9) |
| Hoshi et al. 2012 | Original study (Tsurugaya Project) | The Tsurugaya Project was a Comprehensive Geriatric Assessment conducted on older residents in Sendai, Japan. | NA | NA | residents of Sendai city; aged ≥70 years; consent to use of information in the survey; consent to use of information on their certification for long-term care insurance | already been certified for long-term care insurance; physical function data could not be obtained at baseline | ≥70 | B | N | 2003 | NR | 0 | 0 |
| Huang et al. 2010 | Original study | Patients from two geriatric clinics in Pittsburgh and Baltimore, USA | NA | NA | Age ≥65 years; community-dwelling; MMSE ≥24; SPPB score of 3 to 10; No self-reported difficulties in seven ADLs | terminal health condition; nursing home resident; diagnosis of progressive dementing | ≥65 | B | Y | NR | NR | 35 (32%) | 18 months follow-up: changed MD appointment (12); missed home visit (9) deceased (7) subjects dropped (3) |

| Authors | Study | Sample/region | Study: inclusion criteria | Study: exclusion criteria | Report: inclusion criteria | Report: exclusion criteria | Minimum age | Gender (M/F/B) | Cognition excluded* | Start date | End date | Loss to follow-up/excluded; n (%) | Description of loss to follow-up |
|---|---|---|---|---|---|---|---|---|---|---|---|---|---|
| | | | | | | | | | | | | | ADL questions not answered (4) |
| Lee at al. 2020 | Original study | Study is based on the National Health Insurance Service-Senior Cohort database, including randomly sampled older people registered with the Korean National Health Insurance service. | NA | NA | Participation in the Korean National Screening Program for Transitional Ages (NSPTA) during 2007–2008 | Participants who received the LTCI service before 2010; participants with missing TUG data or other covariates; impaired ADL at baseline | 66 | B | N | Jan 2010 | Dec 2013 | 0 | NA |
| Makizako et al. 2015 | OSHPE | Obu City, Aichi Prefecture, Japan | Age of ≥65 years at examination in 2011 or 2012; being a resident of Obu, Nagoya, Japan | Previous participation in other studies; disability in basic activities of daily living (self-feeding, personal hygiene and grooming, walking, climbing stairs and bathing); inability to undergo performance-based assessments (eg, severe hypertension, balance impairment or pain) | None | History of Parkinson's disease, stroke, depression, or Alzheimer's disease; MMSE score <18 points; Missing data for frailty; the need for support or care certified by the Japanese public long-term care insurance system (LTCI; care level ≥3/5) | ≥65 | B | N | Aug 2011 | Feb 2014 | 55 (1.3%) | Died or who moved to another city during the follow-up period |
| Makizako et al. 2017 | OSHPE | Obu City, Aichi Prefecture, Japan | ≥65 years old at examination; reside in Obu City, Aichi Prefecture, Japan | Participation in another study; hospitalized or in residential care; certified care level >3 (of 5) in the Japanese LTCI system. | Participation in follow up assessments | disability in basic ADLs (self-feeding, personal hygiene and grooming, walking, stair climbing, and bathing); inability to undergo the FTSS and the TUG; history of Parkinson's disease or stroke; MMSE scores <18/30; deceased during 2 year follow up; move to another city during 2 year follow up | ≥65 | B | Y | Aug 2011 | NR | 55 (1%) | Deceased (34); moving to another city (18); unknown (3) |
| Minneci et al. 2015 | ICARe Dicomano | Dicomano, a small rural town near Florence, Italy | Living in Dicomano (recorded in the city registry office); community-dwelling; ≥65 years | living in a nursing home/long-term care facility | None | prevalent ADL disability | ≥65 | B | N | 1996 | 1999 | 108 (19%) | No reason reported. |
| Montero-Odasso et al. 2005 | EFA study | Cohort of well-functioning elderly people affiliated with a health maintenance organization (HMO) based at a University Hospital in Buenos Aires, Argentina. | Aged ≥75 years; Community-dwelling | NR | NR | Cognitive impairment; Depression; unstable chronic disease; life expectancy <12 months or terminal illness; gait disorders related to a neurological cause; use of cane or walking devices; | ≥75 | B | N | Jan 2000 | Dec 2002 | 1 (1%) | No reason reported. |

| Authors | Study | Sample/region | Study: inclusion criteria | Study: exclusion criteria | Report: inclusion criteria | Report: exclusion criteria | Minimum age | Gender (M/F/B) | Cognition excluded* | Start date | End date | Loss to follow-up/excluded; n (%) | Description of loss to follow-up |
|---|---|---|---|---|---|---|---|---|---|---|---|---|---|
| | | | | | | disability in the ADLs;<br>inability to attend appointments | | | | | | | |
| Moriya et al. 2013 | Hokkaido study by Moriya et al. 2009 | Baseline survey performed during a dental health examination sponsored by the public authorities in two rural communities (Tomamae and Iwanai) in Hokkaido prefecture, Japan | Residing in two rural communities (Tomamae and Iwanai) Hokkaido prefecture;<br>aged ≥65 years;<br>residing in the community | Persons in institutions or in need of long-term care | None | None | ≥65 | B | N | Jul 2004 (Tomamae);<br>Aug 2005 (Iwanai) | NR | 98 (11%) | Aged ≥85 years (25);<br>had missing data (10);<br>died without LTCI certification during the 5-year follow-up period (56);<br>were already certified in LTCI at baseline (7). |
| Onder et al. 2005 | WHAS-I | Medicare beneficiaries living in the geographic catchment area of Baltimore City and Baltimore country, USA | Female;<br>Medicare beneficiaries;<br>Age ≥65 years;<br>Living in the geographic catchment area of the study (Baltimore City and Baltimore country, US);<br>Community-dwelling;<br>reported difficulty in two or more of four functional domains (mobility and exercise tolerance, upper extremity function, basic selfcare, and higher functioning tasks of independent living);<br>scored >17 on the Mini Mental State Examination | Living in a nursing home;<br>MMSE <18 points;<br>unability to answer the screening interview by self-report | None | Participants who reported a specific disability at baseline or at the first 6-month follow-up;<br>missing data on one or more performance measures at baseline | ≥65 | F | Y | NR | NR | 4 (1%) | Missing data on both assessments before the onset of disability (4) |
| Ostir et al. 1998 | H-EPESE | H-EPESE 1993-1994 baseline sample;<br>Noninstitutionalized Mexican Americans aged 65-99 from five Southwestern states: Texas, California, New Mexico, Colorado, and Arizona, USA. | Community-dwelling;<br>Mexican American;<br>living in Texas, California, New Mexico, Colorado, or Arizona;<br>aged 65-99 years | None | Participants from H-EPESE 1993-1994 sample;<br>reported no baseline disability in ADLs (bathing, using the toilet, transferring from bed to chair, and walking across a small room);<br>capable of walking a half mile and climbing stairs without help | At follow-up:<br>Deceased;<br>refused to be reinterviewed;<br>moved to Mexico;<br>could not be located | ≥65 | B | N | 1993 | 1996 | 581 (30%) | NR |
| Rosenberg et al. 2019 | Original study | Frailty and Ageing Cohort Study (FACTS), including elderly community-dwelling people,receiving home-based primary in Victoria, Canada. | NA | NA | living in the community;<br>receiving home-based primary care from one interdisciplinary geriatric medical practice in Victoria, Canada, between 1 May, 2017, and 30 October, 2018;<br>age ≥70 years;<br>difficulty accessing office-based care;<br>presence of a frailty syndrome (eg, dementia, falls and chronic pain);<br>multiple comorbidities;<br>need for complex interdisciplinary medical care | None | ≥70 | B | N | May 2017 | Oct 2019 | 0 | NA |
| Sakamoto et al. 2016 | Tosa Longitudinal Aging Study (TLAS) | Unselected community-dwelling elderly population recorded in the town registry office in Tosa, a rural town in Kochi prefecture, Japan | Aged ≥65 years;<br>written informed consent;<br>resident in rural town in Kochi prefecture, Japan | Aged ≥75 years when participating in the community-based health check-ups in Tosa in 2004; | None | None | ≥75 | B | N | 2004 | 2006 | 16 (8.5%) | 2 died (1%);<br>14 did not participate in the follow-up examination in 2006 (7%) |

| Authors | Study | Sample/region | Study: inclusion criteria | Study: exclusion criteria | Report: inclusion criteria | Report: exclusion criteria | Minimum age | Gender (M/F/B) | Cognition excluded* | Start date | End date | Loss to follow-up/excluded; n (%) | Description of loss to follow-up |
|---|---|---|---|---|---|---|---|---|---|---|---|---|---|
| | | | | full score in basic ADL at the time of the check-up in 2004. | | | | | | | | | |
| Shinkai et al. 2003 | TMIG-LISA | Nangai Village sample; older adults living in Nangai Village, a rural and mainly agricultural area of Akita Prefecture in Japan. | Older individuals living in Koganei City or Nangai Village, Japan; age 65 to 84 years (Koganei) or ≥65 years (Nangai) | None | community dwelling; free from BADL and IADL disability | living in institutions; homebound due mainly to mobility difficulty; long-term absent | ≥65 | B | N | 1992 | 1998 | 0 | NA |
| Stenholm et al. 2014 | InCHIANTI | Random sample from two Italian towns located in the Chianti geographic area, located outside the urban area of Florence, Italy; baseline data of 1998 wave. | Aged ≥65 years | None | Participation in the physical performance measurements at baseline and at the 3-year follow-up; free of ADL disability at baseline (1998–2000) and 3-year follow-up (2001–2003) | None | ≥65 | B | N | 1998 | 2008 | 82 (9%) for 3-year follow-up<br><br>174 (19%) for 6-year follow-up | 3 year: Died (56); did not participate (26)<br><br>6 year: Died (136); unable to participate (38) |
| Studenski et al. 2003 | Original study | Primary care programs of a Medicare health maintenance organization (HMO) and Veterans Affairs (VA) system | NA | NA | aged ≥65 years; lived in the community within a 20-mile radius of the ambulatory clinic site; had been in the same healthcare system for at least 1 year; MMSE ≥24 or 16–23 (with caregiver) | MMSE <16; unable to walk at least 4 meters; those considered to be extremely fit or extremely fragile (gait speed >1.3 m/s or <0.2 m/s) | ≥65 | B | Y | NR | NR | 34 (7%) | Changed provider systems (20); withdrew (12); moved out of the study area (2) |
| Sugiura et al. 2013 | Original study | Japanese elderly living in Takatsuki City, japan (urban area and suburb in the northern part of Osaka Prefecture) | NA | NA | registered at or used welfare centers for the aged and community centers; community-dwelling; aged ≥65; independent in terms of BADLs (measured using a modified Katz's activities of daily living (ADL) scale, which consists of 5 items: bathing, dressing, walking, continence and feeding; independent in higher-level competence according to the baseline survey, the Tokyo Metropolitan Institute of Gerontology Index of Competence (TMIG-IC). | None | ≥65 | B | N | May and Jun 2007 | May and Jun 2011 | 58 (13%) | Hospitalized (8); had moved or could not be reached (39); died (11) |
| Tsutsumimoto et al. 2016 | OSHPE | Obu City, Aichi Prefecture, Japan | ≥65 years old at examination; reside in Obu City, Aichi Prefecture, Japan | Participation in another study; hospitalized or in residential care; certified care level >3 (of 5) in the Japanese LTCI system. | None | history of Parkinson disease, Alzheimer disease or stroke; severe cognitive impairment (MMSE <19 points); requiring support or care by the LTCI system at baseline; missing values at baseline assessment | ≥65 | B | Y | Aug 2011 | NR | 0 | NA |
| Vaarst et al. 2021 | Healthy Ageing Network of Competence (HANC) study | Community-dwelling residents from Denmark | Community-dwelling residents aged 75 years and older; able to understand written and oral Danish; consented to a preventive home visit (PHV) | Not medically stable because of serious diseases (e.g. cancer, severe heart failure) surgery within the past 6 months; fracture within the past 3 months; | Participation in both home visits of the HANC study | Participation in the RCT during the same follow-up time; currently receiving personal care, home nursing care; enrolled in a rehabilitation program at the time | ≥75 | B | Y | Mar 2013 | Nov 2017 | 0 | Censored: admitted to a nursing home (6); died prior to receiving personal care (10) |

| Authors | Study | Sample/region | Study: inclusion criteria | Study: exclusion criteria | Report: inclusion criteria | Report: exclusion criteria | Minimum age | Gender (M/F/B) | Cognition excluded* | Start date | End date | Loss to follow-up/excluded; n (%) | Description of loss to follow-up |
|---|---|---|---|---|---|---|---|---|---|---|---|---|---|
| | | | | cognitive impaired as determined by the Mini-Mental State Examination (score of 21 or less);<br>no written informed consent | | of the preventive home visit;<br>missing performance measures and/or covariate data | | | | | | | |
| Verghese et al. 2012 | EAS | Bronx County, USA | Bronx County population;<br>aged ≥70 years;<br>community residing;<br>English speaking | severe impairments in auditory (unable to follow questions asked in a loud voice) or visual function (corrected vision <20/400);<br>bed bound;<br>institutionalization | None | presence of dementia diagnosed at consensus case conferences;<br>ADL disability;<br>slow gait (1.5 standard deviations below age- and sex-specific mean values established in the EAS cohort) | ≥70 | B | Y | Dec 2004 | Apr 2011 | 37 (6%) | Major causes for attrition included awaiting study visit and loss of contact |
| Woo et al. 1999 | Original study | Hong Kong Chinese residents, selected by stratified random sampling from a registered list of all recipients of Old Age and Disability Allowances. | Hong Kong Chinese resident;<br>aged ≥70 years | None | Ability to complete the 16-foot walk | None | ≥70 | B | N | 1991 | NR | 664 (36%) | Lost to follow-up or died (664) |
| Zhang et al. 2013 | InCHIANTI | Random sample from two Italian towns located in the Chianti geographic area, located outside the urban area of Florence, Italy;<br>baseline data of 1998 wave. | Aged ≥65 years | None | Aged ≥60 years;<br>MMSE score ≥18 points;<br>able to walk 7 m at a self-selected speed without using an assistance of a walker;<br>free from falls and ADL- and IADL-related disability at baseline | None | ≥60 | B | Y | 1998 | 2003 | 0 | NA |

**Abbreviations:**

ADL = activities of daily living; B = both; BMI = body mass index; Boston RISE = Boston Rehabilitative Impairment Study of the Elderly; CHS = Cardiovascular Health Study; EAS = Einstein Aging Study; EFA = Estudio de Evaluacion Funcional del Anciano study; EPESE = Established Populations for the Epidemiologic Study of the Elderly; F = female; FTSS = Five-Times Sit-to-Stand test; HANC = Healthy Ageing Network of Competence; HEPESE = Hispanic EPESE; HMO = Medicare health maintenance organization; IADL = instrumental activities of daily living; ICARe = Insufficienza Cardiaca negli Anziani Residenti a Dicomano Study; InCHIANTI = Aging in the Chianti area; KLSAH = Kusatsu Longitudinal Study on Aging and Health; LTCI = Long term care insurance; M = male; MAP = Rush Memory and Aging Project; MD = medical doctor; MMSE = Mini-mental state examination; N = no; NA = not applicable; TUG = Timed Up and Go test; NOS = Newcastle-Ottawa scale; NR = not reported; NSPTA = Korean National Screening Program for Transitional Ages; OSHPE = Obu Study of Health Promotion for the Elderly; PEP = Precipitating Events Project; PHV = preventive home visit; PSC = Project Safety Cohort; PSNT = Porvoo Sarcopenia and Nutrition Trial; RCT = randomized controlled trial; ROAD = Research on Osteoarthritis/Osteoporosis Against Disability; SABE = “Salud Bienestar y Envejecimiento en America Latina y el Caribe / Saude, Bem-Estar e Envelhecimento [Health, Wellbeing and Aging]; SD = standard deviation; SNAC = Swedish National study on Aging and Care; SOF = Study of Osteoporotic Fractures; SPPB = Short Physical Performance Battery; TILDA = Irish Longitudinal Study on Aging; TLSA = Tosa Longitudinal Aging Study; TMIG-IC = Tokyo Metropolitan Institute of Gerontology Index of Competence; TMIG-LISA = Tokyo Metropolitan Institute of Gerontology Longitudinal Interdisciplinary Study on Aging; VA = Veterans Affairs; WHAS-I = Women’s Health and Aging Study I; Y = yes

### eTable 9: Overview of cohort studies

| Authors/report | Study: acronym | Study: full title | Sample/wave/data set (first/normal baseline and follow-up if not reported otherwise) |
|---|---|---|---|
| Abe et al. 2019 | KLSAH | Kusatsu Longitudinal Study on Aging and Health | Data from 2003 and 2013 waves |
| Akune et al. 2014 | ROAD | Research on Osteoarthritis/Osteoporosis Against Disability (ROAD) study | |
| Artaud et al. 2015 | 3C Study | Three-City Study | City of Dijon sub-sample |
| Balzi et al. 2010 | InCHIANTI | Invecchiare in Chianti (Aging in the Chianti Area) | Baseline data of 1998 wave |
| Beauchamp et al. 2015 | Boston RISE | Boston Rehabilitative Impairment Study of the Elderly | |
| Bjorkman et al. 2019 | PSNT | Porvoo Sarcopenia and Nutrition Trial | Individuals screened for or included in RCT |
| Buchman et al. 2021 | MAP | Rush Memory and Aging Project | |
| Chaudhry et al. 2010 | CHS | Cardiovascular Health Study | Baseline: Initial 1989 assessment, incorporating the African-American cohort beginning in 1992 |
| Chu et al. 2006 | n.a. | Hong Kong study by Chu 2005 | |
| da Silva Alexandre et al. 2012 | SABE | Saúde, Bem-Estar e Envelhecimento (Health, Wellbeing and Aging) | |
| DeVore et al. 1994 | n.a. | No parent study | |
| Diem et al. 2018 | SOF | Study of Osteoporotic Fractures | Participants of SOF Year 20 (Y20) examination |
| Doi et al. 2020 | OSHPE | Obu Study of Health Promotion for the Elderly | |
| Donoghue et al. 2014 | TILDA | The Irish Longitudinal Study on Ageing | Data from first wave (January 2009 to July 2011; baseline); follow-up data March2012 to December 2012 |
| Gill et al. 1995 (J Gerontol) | n.a. | Project Safety cohort | |
| Gill et al. 1995 (JAGS) | n.a. | Project Safety cohort | |
| Gill et al. 2004 | PEP | Precipitating Events Project | |
| Guralnik et al. 1995 | EPESE | Established Populations for the Epidemiologic Study of the Elderly | |
| Guralnik et al. 2000 | EPESE | Established Populations for the Epidemiologic Study of the Elderly | |
| Heiland et al. 2016 | SNAC | Swedish National study on Aging and Care in Kungsholmen | SNAC-K sub-sample (Kungsholmen area, central Stockholm, Sweden) |
| Hoshi et al. 2012 | n.a. | Tsurugaya Project | |
| Huang et al. 2010 | n.a. | Studenski et al. 2004 Study | |
| Lee at al. 2020 | n.a. | Korean National Health Insurance Service-Senior Cohort database | |
| Makizako et al. 2015 | OSHPE | Obu Study of Health Promotion for the Elderly | |
| Makizako et al. 2017 | OSHPE | Obu Study of Health Promotion for the Elderly | |

| Authors/report | Study: acronym | Study: full title | Sample/wave/data set (first/normal baseline and follow-up if not reported otherwise) |
|---|---|---|---|
| Minneci et al. 2015 | ICARe Dicomano | Insufficienza Cardiaca negli Anziani Residenti a Dicomano Study | |
| Montero-Odasso et al. 2005 | EFA study | Estudio de Evaluacion Funcional del Anciano' study | |
| Moriya et al. 2013 | n.a. | Hokkaido study by Moriya et al. 2009 | |
| Onder et al. 2005 | WHAS-I | Women's Health and Aging Study I | |
| Ostir et al. 1998 | H-EPESE | Hispanic Established Populations for the Epidemiologic Studies of the Elderly | H-EPESE 1993-1994 baseline sample |
| Rosenberg et al. 2019 | FACTS | The Frailty and Ageing Cohort Study | |
| Sakamoto et al. 2016 | TLAS | Tosa Longitudinal Aging Study | |
| Shinkai et al. 2003 | TMIG-LISA | Tokyo Metropolitan Institute of Gerontology Longitudinal Interdisciplinary Study on Aging | Nangai Village sub-sample |
| Stenholm et al. 2014 | InCHIANTI | Invecchiare in Chianti (Aging in the Chianti Area) | Baseline data of 1998 wave |
| Studenski et al. 2003 | n.a. | No parent study | |
| Tsutsumimoto et al. 2016 | OSHPE | Obu Study of Health Promotion for the Elderly | |
| Vaarst et al. 2021 | HANC | Healthy Ageing Network of Competence study | |
| Verghese et al. 2012 | EAS | Einstein Aging Study | |
| Woo et al. 1999 | n.a. | No parent study | |
| Zhang et al. 2013 | InCHIANTI | Invecchiare in Chianti (Aging in the Chianti Area) | Baseline data of 1998 wave. |

**Abbreviations:**

Boston RISE = Boston Rehabilitative Impairment Study of the Elderly; CHS = Cardiovascular Health Study; EAS = Einstein Aging Study; EFA = Estudio de Evaluacion Funcional del Anciano study; EPESE = Established Populations for the Epidemiologic Study of the Elderly; HANC = Healthy Ageing Network of Competence; HEPESE = Hispanic EPESE; ICARe = Insufficienza Cardiaca negli Anziani Residenti a Dicomano Study; InCHIANTI = Aging in the Chianti area; KLSAH = Kusatsu Longitudinal Study on Aging and Health; MAP = Rush Memory and Aging Project; na = not applicable; OSHPE = Obu Study of Health Promotion for the Elderly; PEP = Precipitating Events Project; PSC = Project Safety Cohort; PSNT = Porvoo Sarcopenia and Nutrition Trial; RCT = randomized controlled trial; ROAD = Research on Osteoarthritis/Osteoporosis Against Disability; SABE = "Salud Bienestar y Envejecimiento en America Latina y el Caribe / Saude, Bem-Estar e Envelhecimento [Health, Wellbeing and Aging]; SNAC = Swedish National study on Aging and Care; SOF = Study of Osteoporotic Fractures; TILDA = Irish Longitudinal Study on Aging; TLSA = Tosa Longitudinal Aging Study; TMIG-LISA = Tokyo Metropolitan Institute of Gerontology Longitudinal Interdisciplinary Study on Aging; WHAS-I = Women's Health and Aging Study I

### eTable 10: Detailed description of gait speed assessment conduction in the included reports

| Author(s)/report | Part of SPPB (with reference) | Instruction on speed | Distance | Walking start vs. standing start | Deceleration | Computerized walkway vs. over ground (OG if NR) | Stopwatch or other method | Counting | Setting | Person measuring | Feasibility notes |
|---|---|---|---|---|---|---|---|---|---|---|---|
| Abe et al. 2019 | no | usual | 5 m | 3 m | 3 m | Over ground (straight walkway) | NR | NR | Local public health center or gymnasium | Well-trained staff | NR |
| Akune et al. 2014 | no | usual | 6 m | NR | NR | over ground (hallway) | NR | NR | Clinic | NR | NR |
| Artaud et al. 2015 | no | fast | 6 m | started walking 3 m before the start line | stopped walking after the end line = deceleration time not included in walking speed | over ground | two photoelectric cells connected to a chronometer. | NR | Study center & home | NR | wave 6 participants: Gait speed was measured at home over 6 m in most instances (85%); shorter distances (3.5–5.9 m) were used if there was not enough space at home. |
| Beauchamp et al. 2015 | yes; Guralnik et al. 1994 | usual | 4 m | SPPB protocol | SPPB protocol | SPPB protocol | SPPB protocol | SPPB protocol | Rehabilitation Hospital | Nurse | NR |
| Buchman et al. 2021 | no | usual | 2.4 m | NR | NR | over ground | NR | on a marked 8 ft course back and forth twice without stopping; counting NR | Residential facilities or the participants' homes | NR | NR |
| Chaudhry et al. 2010 | no | usual | 15 ft = 4.6 m | NR | NR | over ground | NR | NR | Clinic | NR | NR |
| Chu et al. 2006 | no | fast | 5 m | NR | NR | over ground | NR | NR | Home visit | NR | NR |
| Diem et al. 2018 | no | usual | 6 m | NR | NR | over ground | NR | NR | Clinic | NR | NR |
| Doi et al. 2020 | no | usual | 2.4 m | 2 m | 2 m | over ground (straight and flat pathway) | stopwatch | mean gait speed over five trials | Clinic | Well-trained staff who had nursing, allied health, or similar qualifications | NR |
| Donoghue et al. 2014 | no | usual | 4.88 m | 2.5 m | 2.0 m | computerized walkway | computerized | 2 walks, which were combined | Health center | Research nurses | This test was performed in the health center but was not available during home assessments |
| Gill et al. 1995 (593) | no | fast | 20 ft = 6.1 m (back and forth over a 10-foot course) | NR | NR | over ground | NR | NR | Home | Trained research nurse | NR |

| Author(s)/report | Part of SPPB (with reference) | Instruction on speed | Distance | Walking start vs. standing start | Deceleration | Computerized walkway vs. over ground (OG if NR) | Stopwatch or other method | Counting | Setting | Person measuring | Feasibility notes |
|---|---|---|---|---|---|---|---|---|---|---|---|
| Gill et al. 2004 | no | fast | 20 ft = 6.1 m (back and forth over a 10-foot course) | NR | NR | over ground | NR | NR | Home | Trained research nurse | NR |
| Gill. et al. 1995 (9579) | no | fast | 20 ft = 6.1 m (back and forth over a 10-foot course) | NR | NR | over ground | NR | NR | Home | Trained research nurse | NR |
| Guralnik et al. 2000 | yes; Guralnik et al. 1994 | usual | 8 ft = 2.4 m | standing start | SPPB protocol | SPPB protocol | SPPB protocol | Time on the faster of two walks was used | Home | Specially trained interviewers | NR |
| Heiland et al. 2016 | no | self-selected | 2.4 m or 6 m | NR | NR | over ground | NR | NR | Clinical centers (most); according to participants' living situation: living in their homes or in institutions | Nurse | Subjects who were unable to walk without personal support received the lowest possible score, i.e. 0 m/s. |
| Hoshi et al. 2012 | no | fast | 10 m | 3 m acceleration | 3 m deceleration | flat floor | NR | Better results of two trials | NR | NR | |
| Huang et al. 2010 | yes; Guralnik et al. 1994 | usual | 4 m | SPPB protocol | SPPB protocol | SPPB protocol | SPPB protocol | The better of two measures. | Home | Trained assessors | usually completed in 2 minutes using a stopwatch and a 4-m tape; adverse events not reported |
| Makizako et al. 2015 | no | usual | 2.4 m | 2 m | 2 m | over ground (flat and straight surface) | stopwatch | NR | Clinic | Trained physical therapists | NR |
| Minneci et al. 2015 | yes; Guralnik et al. 1995 | usual | 4 m | SPPB protocol | SPPB protocol | SPPB protocol | SPPB protocol | fastest speed in two attempts | Home | Study physicians | NR |
| Montero-Odasso et al. 2005 | no | usual | 8 m | 1 m | 1 m | over ground | timed by a chronometer | one nontimed practice trial; final score was the time of the quicker of two timed trials. | Ambulatory clinics | Trained geriatricians | NR |
| Onder et al. 2005 | yes; Guralnik et al. 1994 | usual | 4 m | SPPB protocol | SPPB protocol | SPPB protocol | SPPB protocol | faster of two walks | Home | Trained examiners | NR |
| Ostir et al. 1998 | yes; Guralnik et al. 1994; Guralnik et al. 1995 | usual | 8 ft = 2.4 m | SPPB protocol | SPPB protocol | SPPB protocol | SPPB protocol | faster of two walks | Home | NR | NR |

| Author(s)/report | Part of SPPB (with reference) | Instruction on speed | Distance | Walking start vs. standing start | Deceleration | Computerized walkway vs. over ground (OG if NR) | Stopwatch or other method | Counting | Setting | Person measuring | Feasibility notes |
|---|---|---|---|---|---|---|---|---|---|---|---|
| Rosenberg et al. 2019 | no | usual | 3 m | NR | NR | over ground | NR | NR | Home | Practice nurses | NR |
| Shinkai et al. 2003 | no | fast + usual | 5 m | 3 m acceleration | 3 m deceleration | over ground | NR | NR | Municipal community halls | NR | NR |
| Stenholm et al. 2014 | yes; Guralnik et al. 1994 | usual | 4 m | SPPB protocol | SPPB protocol | SPPB protocol | SPPB protocol | faster of two walks | Clinic | Trained geriatricians and physical therapists | NR |
| Studenski et al. 2003 | no | usual | 4 m | 1 m | NR | over ground | NR | NR | Average of the clinic and home observations | NR | NR |
| Tsutsumimoto et al. 2016 | no | usual | 2.4 m | 2 m | 2 m | over ground | NR | NR | NR | Well-trained staff who had nursing, allied health, or similar qualifications | NR |
| Verghese et a. 2012 | no | usual | 15 ft and 20 ft | NR | NR | computerized walkway | computerized walkway | 2 trials on 15 ft walkway; 1 trial on 20 ft walkway | Research center | NR | NR |
| Woo et al. 1999 | no | usual | 16 feet (2 times 8 ft) | NR | NR | over ground | NR | average of two readings | Home | NR | NR |

**Abbreviations:**
SPPB = Short Physical Performance Battery, NR = not reported
Guralnik et al. 1994: Guralnik JM et al. (1994): A short physical performance battery assessing lower extremity function: association with self-reported disability and prediction of mortality and nursing home admission. J Gerontol 49 (2), S. 85-94.
Guralnik et al. 1995: Guralnik JM et al. (1995): Lower-extremity function in persons over the age of 70 years as a predictor of subsequent disability. N. Engl. J. Med 332 (9), S. 556-561.

### eTable 11: Types of ADL components measured in the included reports using incident ADL disability as an outcome (n = 23)

| Author(s)/report | Criteria for ADL disability | Number of criteria to indicate disability | Number of criteria assessed | Bath | Dress | Toilet | Transfer | Eating | Walk | Other criteria for ADL disability | Additional criteria |
|---|---|---|---|---|---|---|---|---|---|---|---|
| Balzi et al. 2010 | Need for help of another person | ≥1 | 6 | X | X | X | X | X | X | | |
| Buchman et al. 2021 | Need for help of another person | ≥1 | 6 | X | X | X | X | X | X | | |
| Chaudhry et al. 2010 | A lot of difficulty or being unable | ≥1 | 6 | X | X | X | X | X | X | | |
| da Silva Alexandre et al. 2012 | Difficulty | ≥1 | 6 | X | X | X | X | X | X | | |
| Diem et al. 2018 | Unable to perform | ≥2 | 5 | X | X | X | X | X | | | |
| Donoghue et al. 2014 | Difficulty | ≥1 | 6 | X | X | X | X | X | X | | |
| Gill et al. 1995 (J Gerontol) | Receiving personal assistance or being completely dependent | ≥1 | 7 | X | X | X | X | X | X | Groom | |
| Gill et al. 1995 (JAGS) | Receiving personal assistance or being completely dependent | ≥1 | 7 | X | X | X | X | X | X | Groom | |
| Gill et al. 2004 | Need for personal assistance | ≥1 | 4 | X | X | | X | | X | | |
| Guralnik et al. 1995 | Inability to perform without help from another person | ≥1 | 4 | X | | X | X | | X | | Mobility-related disability |
| Guralnik et al. 2000 | Inability to perform without help from another person | ≥1 | 4 | X | | X | X | | X | | Mobility-related disability |
| Heiland et al. 2016 | Dependence | ≥1 | 5 | X | X | X | X | X | | | |
| Huang et al. 2010 | Difficulty | ≥1 | 7 | X | X | X | X | X | X | Personal hygiene | |
| Minneci et al. 2015 | Complete inability or need for help | ≥1 | 6 | | X | X | X | X | | Continence; washing hands and face | |
| Onder et al. 2005 | A lot of difficulty or inability to perform | ≥1 | 5 | X | X | X | X | X | | | |
| Ostir et al. 1998 | Self-reported disability | ≥1 | 4 | X | | X | X | | X | | |
| Sakamoto et al. 2016 | Difficulty in carrying out basic ADL independently | ≥1 | 7 | X | X | X | | X | X | Ascending and descending stairs; groom | |
| Shinkai et al. 2003 | Needing help from someone else or being unable to perform | ≥1 | 5 | X | X | | | X | X | Continence | |
| Stenholm et al. 2014 | Inability to perform independently | ≥1 | 6 | X | X | X | X | X | X | | Mobility-related disability |
| Studenski et al. 2003 | Difficulty | ≥1 | 6 | X | X | X | X | X | | Groom | |
| Vaarst et al. 2021 | Need for assistance (personal care) with ADLs such as bathing and dressing | ≥1 | unclear | | | | | | | | |
| Verghese et al. 2012 | Needing assistance or inability to perform | ≥1 | 7 | X | X | X | X | X | X | Groom | |
| Zhang et al. 2013 | Need for assistance | ≥1 | 6 | X | X | X | X | X | X | | |
| | | | | 21 | 19 | 20 | 20 | 18 | 17 | | |

### eTable 12: Quality assessment of included reports using the Newcastle–Ottawa Scale (NOS)

| Author(s) | Selection | | | | Comparability | | Outcome | | | Total score |
|---|---|---|---|---|---|---|---|---|---|---|
| | S1 | S2 | S3 | S4 | C1a | C1b | O1 | O2 | O3 | |
| Abe et al. 2019 | 1 | 1 | 1 | 1 | 1 | 1 | 1 | 1 | 1 | 9 |
| Akune et al. 2014 | 1 | 1 | 1 | 1 | 1 | 1 | 1 | 1 | 1 | 9 |
| Artaud et al. 2015 | 1 | 1 | 1 | 1 | 1 | 1 | 1 | 1 | 1 | 9 |
| Balzi et al. 2010 | 1 | 1 | 1 | 1 | 1 | 1 | 0 | 1 | 1 | 8 |
| Beauchamp et al. 2015 | 0 | 1 | 1 | 0 | 0 | 0 | 0 | 1 | 1 | 4 |
| Bjorkman et al. 2019 | 0 | 1 | 1 | 1 | 1 | 1 | 0 | 1 | 1 | 7 |
| Buchman et al. 2021 | 1 | 1 | 1 | 1 | 1 | 1 | 0 | 1 | 1 | 8 |
| Chaudhry et al. 2010 | 1 | 1 | 1 | 1 | 1 | 1 | 1 | 1 | 1 | 9 |
| Chu et al. 2006 | 1 | 1 | 1 | 1 | 0 | 1 | 1 | 0 | 1 | 7 |
| da Silva Alexandre et al. 2012 | 1 | 1 | 1 | 1 | 1 | 1 | 0 | 1 | 1 | 8 |
| DeVore et al. 1994 | 0 | 1 | 1 | 0 | 0 | 0 | 0 | 0 | 1 | 3 |
| Diem et al. 2018 | 0 | 1 | 1 | 1 | 1 | 1 | 0 | 1 | 1 | 7 |
| Doi et al. 2020 | 1 | 1 | 1 | 1 | 1 | 1 | 1 | 1 | 1 | 9 |
| Donoghue et al. 2014 | 1 | 1 | 1 | 1 | 0 | 0 | 0 | 1 | 1 | 6 |
| Gill et al. 2004 | 1 | 1 | 1 | 1 | 1 | 1 | 0 | 1 | 1 | 8 |
| Gill et al. 1995 (J Gerontol) | 0 | 1 | 1 | 1 | 0 | 0 | 0 | 0 | 1 | 4 |
| Gill et al. 1995 (JAGS) | 1 | 1 | 1 | 1 | 1 | 1 | 0 | 0 | 1 | 7 |
| Guralnik et al. 2000 | 1 | 1 | 1 | 1 | 1 | 1 | 0 | 1 | 1 | 8 |
| Guralnik et al. 1995 | 1 | 1 | 1 | 1 | 1 | 1 | 0 | 1 | 1 | 8 |
| Heiland et al. 2016 | 1 | 1 | 1 | 1 | 0 | 1 | 1 | 1 | 1 | 8 |
| Hoshi et al. 2012 | 1 | 1 | 1 | 1 | 1 | 1 | 1 | 1 | 1 | 9 |
| Huang et al. 2010 | 0 | 1 | 1 | 1 | 1 | 1 | 0 | 0 | 0 | 5 |
| Lee et al. 2020 | 1 | 1 | 1 | 1 | 1 | 1 | 1 | 1 | 1 | 9 |
| Makizako et al. 2015 | 1 | 1 | 1 | 1 | 1 | 1 | 1 | 1 | 1 | 9 |
| Makizako et al. 2017 | 1 | 1 | 1 | 1 | 1 | 1 | 1 | 1 | 1 | 9 |
| Minneci et al. 2015 | 1 | 1 | 1 | 1 | 1 | 1 | 0 | 1 | 1 | 8 |
| Montero-Odasso et al. 2005 | 1 | 1 | 1 | 1 | 1 | 1 | 1 | 1 | 1 | 9 |
| Moriya et al. 2013 | 1 | 1 | 1 | 1 | 1 | 1 | 1 | 1 | 1 | 9 |
| Onder et al. 2005 | 0 | 1 | 1 | 1 | 1 | 1 | 0 | 1 | 1 | 7 |
| Ostir et al. 1998 | 1 | 1 | 1 | 1 | 1 | 1 | 0 | 1 | 0 | 7 |
| Rosenberg et al. 2019 | 0 | 1 | 1 | 1 | 1 | 1 | 1 | 0 | 1 | 7 |
| Sakamoto et al. 2016 | 1 | 1 | 1 | 1 | 1 | 1 | 0 | 1 | 1 | 8 |
| Shinkai et al. 2003 | 1 | 1 | 1 | 1 | 1 | 1 | 0 | 1 | 1 | 8 |
| Stenholm et al. 2014 | 1 | 1 | 1 | 1 | 1 | 1 | 0 | 1 | 1 | 8 |
| Studenski et al. 2003 | 0 | 1 | 1 | 1 | 0 | 1 | 0 | 0 | 1 | 5 |
| Tsutsumimoto et al. 2016 | 1 | 1 | 1 | 1 | 1 | 1 | 1 | 1 | 1 | 9 |
| Vaarst et al. 2021 | 0 | 1 | 1 | 1 | 1 | 1 | 1 | 1 | 1 | 8 |
| Verghese et al. 2012 | 1 | 1 | 1 | 1 | 1 | 1 | 0 | 1 | 1 | 8 |
| Woo et al. 1999 | 0 | 1 | 1 | 0 | 1 | 1 | 0 | 1 | 0 | 5 |
| Zhang et al. 2013 | 1 | 1 | 1 | 1 | 0 | 1 | 0 | 1 | 1 | 7 |

### eTable 13: Grading of Recommendations, Assessment, Development and Evaluations (GRADE) for each risk factor

| Prognostic factor (clinical outcome assessment of mobility capacity) | Risk Ratio [95% CI] per unit | Number of reports | Number of participants | Study design | Risk of bias | Inconsistency | Imprecision | Indirectness | Publication bias | Other considerations | Certainty of evidence |
|---|---|---|---|---|---|---|---|---|---|---|---|
| Gait speed (usual) | 1.23 [1.18–1.28] per -0.1 m/s | 19 | 26,638 | Observational (longitudinal cohort studies) | Serious (-1) | Serious (-1) | Low (0) | Low (0) | NA | Dose-response gradient (+1) | Moderate |
| Gait speed (fast) | 1.28 [1.19–1.38] per -0.1 m/s | 7 | 8,161 | Observational (longitudinal cohort studies) | Serious (-1) | Serious (-1) | Serious (-1) | Low (0) | NA | Dose-response gradient (+1) | Low |
| Short Physical Performance Battery (SPPB) | 1.30 [1.23–1.38] per 1-point decrease | 11 | 9,183 | Observational (longitudinal cohort studies) | Serious (-1) | Low (0) | Serious (-1) | Low (0) | NA | Dose-response gradient (+1) | Moderate |
| Chair Rise Test (CRT) | 1.07 [1.04–1.10] per 1 additional sec | 7 | 9,450 | Observational (longitudinal cohort studies) | Serious (-1) | Serious (-1) | Low (0) | Low (0) | NA | Dose-response gradient (+1) | Moderate |
| Timed Up and Go test (TUG) | 1.15 [1.09–1.21] per 1 additional sec | 5 | 30,426 | Observational (longitudinal cohort studies) | Low (0) | Low (0) | Serious (-1) | Low (0) | NA | Dose-response gradient (+1) | High |

Study design: A body of observational studies begins at high certainty of evidence.
Risk of bias: Low = All/most reports with an overall low risk of bias; or sensitivity analysis shows that results are similar in the reports at lower and higher risk of bias. Serious = Some reports with overall moderate/high risk of bias. Very serious = Most/all reports with an overall moderate/high risk of bias.
Inconsistency: Rating down based on variability in point estimates and extent of overlap in confidence intervals.
Imprecision: Rating down based on the width of the 95% confidence interval around the pooled estimate (≤ 10% RR).
Indirectness: Rating down based on the generalizability of population and outcome.
Other considerations: GRADE criteria for rating up confidence of evidence: “Large effect” criterion not applicable due to the analyses for different units of measurement perform in this study. “Dose-response gradient” criterion applied to all prognostic factors. “Nature of plausible biases” criterion not applicable as reported by Iorio et al. 2015.

Certainty of evidence: High indicates confidence that the true prognosis (risk ratio) lies close to that of the estimate. Moderate indicates confidence that the true prognosis (risk ratio) is likely to be close to the estimate, but there is a possibility that it is substantially different. Low indicates limited confidence in the estimate, true prognosis (risk ratio) may be substantially different from the estimate. Very low indicates very little confidence in the estimate, the true prognosis (risk ratio) is likely to be substantially different from the estimate.

### eTable 14: Associations between baseline mobility capacity and disability at follow-up

| Author(s)/report | Predictor (mobility assessment) | Continuous | Categorial | High mobility (reference) | Low mobility | Disability outcome | Disability categories | Association | 95% CI | p-value |
|---|---|---|---|---|---|---|---|---|---|---|
| Abe et al. 2019 | Gait speed (usual) | X | | One unit was defined as 0.1 m/s for gait speed | NA | Incident LTCI certification | No LTCI certification vs. LTCI certification | HR: 0.86 | 0.81 – 0.92 | p < 0.001 |
| Akune et al. 2014 | CRT (x5) | X | | Per +1 s increase | NA | Incident LTCI certification | No LTCI certification vs. LTCI certification or dead | HR: 1.06 | 1.03 – 1.10 | NR |
| Akune et al. 2014 | Gait speed (usual) | X | | Per +0.1 m/s increase | NA | Incident LTCI certification | No LTCI certification vs. LTCI certification or dead | HR: 0.84 | 0.79 – 0.90 | NR |
| Artaud et al. 2015 | Gait speed (fast) | X | | Increase per standard deviation (0.22 m/s) | NA | Incident disability | Independent vs. dependent | HR: 1.77 | 1.60 – 1.94 | NR |
| Balzi et al. 2010 | SPPB | | X | ≥10 points | 6 – 9 points | Incident ADL | Independent vs. dependent | OR: 2.28 | 1.05 – 4.94 | NR |
| Balzi et al. 2010 | SPPB | | X | ≥10 points | <6 points | Incident ADL | Independent vs. dependent | OR: 17.0 | 6.46 – 44.9 | NR |
| Beauchamp et al. 2015 | 400m-walk-test | X | | continues | NA | LLFDI disability limitation | Continues scale | $R^2$: 0.07 | NR | NR |
| Beauchamp et al. 2015 | 400m-walk-test | X | | continues | NA | LLFDI disability frequency | Continues scale | $R^2$: 0.13 | NR | NR |
| Beauchamp et al. 2015 | gait speed (usual) | X | | continues | NA | LLFDI disability limitation | Continues scale | $R^2$: 0.10 | NR | NR |
| Beauchamp et al. 2015 | Gait speed (usual) | X | | continues | NA | LLFDI disability frequency | Continues scale | $R^2$: 0.15 | NR | NR |
| Beauchamp et al. 2015 | SPPB | X | | continues | NA | LLFDI disability limitation | Continues scale | $R^2$: 0.13 | NR | NR |
| Beauchamp et al. 2015 | SPPB | X | | continues | NA | LLFDI disability frequency | Continues scale | $R^2$: 0.16 | NR | NR |
| Bjorkman et al. 2019 | SPPB | X | | per unit increase | | Use of home care services | No use vs. use of home care services | OR: 0.79 | 0.70 – 0.88 | <0.001 |
| Buchman et al. 2021 | Gait speed (usual) | X | | per 0.2 m/s | NA | Incident ADL | Independent vs. dependent | HR: 0.65 | 0.58 – 0.74 | ≤0.001 |
| Chaudhry et al. 2010 | Gait speed (usual) | | X | The highest four quintiles for sex and height in m/s (no figures reported) | The lowest quintile for sex and height in m/s (no figures reported) | Incident ADL | Independent vs. dependent | HR: 3.03 | 2.56 – 3.59 | <0.001 |
| Chu et al. 2006 | Gait speed (fast) | | X | ≥ 0.56 m/s | < 0.56 m/s | Decline in BI score | Decliners versus non-decliners | RR: 3.83 | 2.20 – 6.65 | < 0.001 |
| da Silva Alexandre et al. 2012 | CRT (x5) | X | | Per +1 s increase | NA | Incident ADL | Independent vs. dependent | M: OR: 1.11<br>F: OR: 1.03 | M: 1.03 – 1.20<br>F: 0.98 – 1.08 | M: 0.008<br>F: 0.137 |
| da Silva Alexandre et al. 2012 | One-leg standing time | X | | Per +1 s increase | NA | Incident ADL | Independent vs. dependent | M: OR: 0.95<br>F: OR: NR | M: 0.84 – 1.08 | M: 0.432<br>F: NR |
| DeVore et al. 1994 | POMA | | X | High mobility: >18 points; n=112 (91%) | Low mobility: <19 points; n=11 (9%) | Change in living situation | Unchanged vs. Nursing home admission or joining relative's households | Sen: 77%<br>Spe: 99% | NR | NR |
| Diem et al. 2018 | Gait speed (usual) | | X | Fast: ≥0.9 m/s | Slow: >0.6 to <0.9 m/s | ADL dependency | Independent vs. dependent or dead | RR: 0.78 | 0.66 – 0.89 | NR |
| Diem et al. 2018 | Gait speed (usual) | | X | Fast: ≥0.9 m/s | Slow: ≤0.6 m/s | ADL dependency | Independent vs. dependent or dead | RR: 0.40 | 0.29 – 0.52 | NR |
| Diem et al. 2018 | Gait speed (usual) | | X | Fast: ≥0.9 m/s | Slow: >0.6 to <0.9 m/s | ADL dependency | Independent vs. dependent among those alive | RR: 0.85 | 0.77 – 0.94 | NR |
| Diem et al. 2018 | Gait speed (usual) | | X | Fast: ≥0.9 m/s | Slow: ≤0.6 m/s | ADL dependency | Independent vs. dependent among those alive | RR: 0.51 | 0.39 – 0.63 | NR |

| Author(s)/report | Predictor (mobility assessment) | Continuous | Categorial | High mobility (reference) | Low mobility | Disability outcome | Disability categories | Association | 95% CI | p-value |
|---|---|---|---|---|---|---|---|---|---|---|
| Doi et al. 2020 | Gait speed (usual) | X | | per 0.1 m/s | NA | Incident LTCI certification | No LTCI certification vs. LTCI certification | HR: 0.83 | 0.79 – 0.87 | <0.001 |
| Doi et al. 2020 | Gait speed (usual) | | X | >1.10 m/s | ≤1.10 m/s | Incident LTCI certification | No LTCI certification vs. LTCI certification | AUC: 0.74 | 0.71 – 0.76 | <0.001 |
| Doi et al. 2020 | Gait speed (usual) | | X | >1.10 m/s | ≤1.10 m/s | Incident LTCI certification | No LTCI certification vs. LTCI certification | HR: 2.06 | 1.65 – 2.57 | <0.001 |
| Donoghue et al. 2014 | Gait speed (usual) | X | | continues | NA | Incident ADL | Independent vs. dependent | AUC: 0.70 | 0.62 – 0.78 | NR |
| Donoghue et al. 2014 | Gait speed (usual) | X | | in 10 cm/s intervals | NA | Incident ADL | Independent vs. dependent | P per score | NR | NR |
| Donoghue et al. 2014 | TUG | X | | continues | NA | Incident ADL | Independent vs. dependent | AUC: 0.67 | 0.58 – 0.75 | NR |
| Donoghue et al. 2014 | TUG | X | | in 1s intervals | NA | Incident ADL | Independent vs. dependent | P per score | NR | NR |
| Gill et al. 1995 (J Gerontol) | 10-foot taps | | X | Normal: ≤7 sec | Abnormal: >7 sec | Incident ADL | Independent vs. dependent | RR: 2.1 | 1.1 – 3.9 | NR |
| Gill et al. 1995 (J Gerontol) | 360° turn | | X | Normal: ≤4 sec | Abnormal: >4 sec | Incident ADL | Independent vs. dependent | RR: 2.7 | 1.4 – 5.1 | NR |
| Gill et al. 1995 (J Gerontol) | CRT (3x) | | X | Normal: ≤10 sec | Abnormal: >10 sec | Incident ADL | Independent vs. dependent | RR: 4.4 | 2.3 – 8.3 | NR |
| Gill et al. 1995 (J Gerontol) | Gait speed (fast) | | X | Normal: ≤11 sec for 20 feet (10 feet back and forth = 6.1m): ≥0.55 m/s | Abnormal: >11 sec for 20 feet (10 feet back and forth = 6.1m): <0.55 m/s | Incident ADL | Independent vs. dependent | RR: 6.4 | 3.0 – 14.0 | NR |
| Gill et al. 1995 (J Gerontol) | time to bend over and pick up a pen | | X | Normal: ≤3 sec | Abnormal: >3 sec | Incident ADL | Independent vs. dependent | RR: 2.6 | 1.3 – 5.2 | NR |
| Gill et al. 2004 | Gait speed (fast) | | X | Fast: ≤10 sec for 20 feet (10 feet back and forth = 6.1m): ≥0.61 m/s | Slow: >10 sec for 20 feet (10 feet back and forth = 6.1m): <0.61 m/s | Incident ADL | Independent vs. dependent | HR: 2.09 | 1.67 – 2.62 | NR |
| Gill et al. 2004 | Gait speed (fast) | | X | Fast: ≤10 sec for 20 feet (10 feet back and forth = 6.1m): ≥0.61 m/s | Slow: >10 sec for 20 feet (10 feet back and forth = 6.1m): <0.61 m/s | Incident ADL disability, incident persistent ADL disability, ADL disability with admission to a nursing home | Independent vs. dependent | KMC | NA | NA |
| Gill. et al. 1995 (JAGS) | 10-foot taps | | X | 2.1 – 5.8 sec | 5.9 – 30.0 sec or unable | Incident ADL | Independent vs. dependent | RR: 1.2 | 0.7 – 2.0 | NR |
| Gill. et al. 1995 (JAGS) | 360° turn | | X | 1.1 – 3.8 sec | 3.9 – 23.0 sec or unable | Incident ADL | Independent vs. dependent | RR: 2.1 | 1.2 – 3.6 | NR |
| Gill. et al. 1995 (JAGS) | CRT (x3) | | X | 2.2 – 8.7 sec | 8.8 – 30.0 sec or unable | Incident ADL | Independent vs. dependent | RR: 2.1 | 1.2 – 3.5 | NR |
| Gill. et al. 1995 (JAGS) | Gait speed (fast) | | X | Normal: 4.1 – 9.4 sec for 20 feet/6.1m: ≥0.65 m/s | Abnormal: 9.5 – 60.0 sec for 20 feet/6.1m or unable: <0.65 m/s | Incident ADL | Independent vs. dependent | RR: 2.4 | 1.4 – 4.2 | NR |
| Gill. et al. 1995 (JAGS) | Time to bend over and pick up a pen | | X | 1.0 – 3.0 sec | 3.1 – 24.6 sec or unable | Incident ADL | Independent vs. dependent | RR: 1.7 | 1.0 – 2.9 | NR |
| Guralnik et al. 1995 | SPPB | | X | 10 – 12 points | 4 – 6 points | Incident ADL | Independent vs. dependent | RR: 4.2 | 2.3 – 7.7 | NR |
| Guralnik et al. 1995 | SPPB | | X | 10 – 12 points | 7 – 9 points | Incident ADL | Independent vs. dependent | RR: 1.6 | 1.0 – 2.6 | NR |
| Guralnik et al. 2000 | Gait speed (usual) | X | | Gait speed was categorized as deciles for these analyses | NA | Incident ADL (1 y) | Independent vs. dependent | AUC: 0.70 | NR | NR |
| Guralnik et al. 2000 | Gait speed (usual) | X | | Gait speed was categorized as deciles for these analyses | NA | Incident ADL (4 y) | Independent vs. dependent | AUC: 0.67 | NR | NR |
| Guralnik et al. 2000 | SPPB | | X | 10 – 12 points | 4 – 6 points | Incident ADL (1, Iowa cohort) | Independent vs. dependent | RR: 5.1 | 1.8 – 14.3 | NR |
| Guralnik et al. 2000 | SPPB | | X | 10 – 12 points | 7 – 9 points | Incident ADL (1 y, Iowa cohort) | Independent vs. dependent | RR: 2.0 | 0.7 – 5.3 | NR |
| Guralnik et al. 2000 | SPPB | | X | 10 – 12 points | 4 – 6 points | Incident ADL (1 y, New Haven cohort) | Independent vs. dependent | RR: 7.4 | 1.8 – 30.5 | NR |

| Author(s)/report | Predictor (mobility assessment) | Continuous | Categorial | High mobility (reference) | Low mobility | Disability outcome | Disability categories | Association | 95% CI | p-value |
|---|---|---|---|---|---|---|---|---|---|---|
| Guralnik et al. 2000 | SPPB | | X | 10 – 12 points | 7 – 9 points | Incident ADL (1 y, New Haven cohort) | Independent vs. dependent | RR: 1.5 | 0.3 – 6.3 | NR |
| Guralnik et al. 2000 | SPPB | | X | 10 – 12 points | 4 – 6 points | Incident ADL (4 y, Iowa cohort) | Independent vs. dependent | RR: 4.5 | 2.6 – 8.0 | NR |
| Guralnik et al. 2000 | SPPB | | X | 10 – 12 points | 7 – 9 points | Incident ADL (4 y, Iowa cohort) | Independent vs. dependent | RR: 1.5 | 0.9 – 2.5 | NR |
| Guralnik et al. 2000 | SPPB | | X | 10 – 12 points | 4 – 6 points | Incident ADL (4 y, North Carolina cohort) | Independent vs. dependent | RR: 4.0 | 2.2 – 7.2 | NR |
| Guralnik et al. 2000 | SPPB | | X | 10 – 12 points | 7 – 9 points | Incident ADL (4 y, North Carolina cohort) | Independent vs. dependent | RR: 1.2 | 0.6 – 2.1 | NR |
| Guralnik et al. 2000 | SPPB | | X | 10 – 12 points | 4 – 6 points | Incident ADL (6 y, New Haven cohort) | Independent vs. dependent | RR: 3.4 | 1.7 – 7.1 | NR |
| Guralnik et al. 2000 | SPPB | | X | 10 – 12 points | 7 – 9 points | Incident ADL (6 y, New Haven cohort) | Independent vs. dependent | RR: 1.2 | 0.7 – 2.2 | NR |
| Heiland et al. 2016 | Gait speed (self-selected) | | X | ≥0.8 m/s | <0.8 m/s | Incident ADL | Independent vs. dependent | OR: 7.2 | 4.4 – 11.6 | ≤0.001 |
| Heiland et al. 2016 | Gait speed (self-selected) | | X | >1.0 m/s (upper tertile) | <0.8 m/s (lower tertile) | Incident ADL | Independent vs. dependent | OR: 15.8 | 7.7 – 32.6 | ≤0.001 |
| Heiland et al. 2016 | Gait speed (self-selected) | | X | >1.0 m/s (upper tertile) | 0.8 – 1.0 m/s (middle tertile) | Incident ADL | Independent vs. dependent | OR: 3.7 | 1.7 – 7.8 | ≤0.001 |
| Heiland et al. 2016 | One-leg standing time | | X | ≥5 sec | <5 sec | Incident ADL | Independent vs. dependent | OR: 3.4 | 2.0 – 5.6 | ≤0.001 |
| Heiland et al. 2016 | One-leg standing time | | X | 60 sec (upper tertile) | <10 sec (lower tertile) | Incident ADL | Independent vs. dependent | OR: 7.1 | 2.0 – 24.6 | ≤ 0.05 |
| Heiland et al. 2016 | One-leg standing time | | X | 60 sec (upper tertile) | 10–59 (middle tertile) | Incident ADL | Independent vs. dependent | OR: 3.0 | 0.8 – 10.6 | >0.05 |
| Hoshi et al. 2012 | FRT | | X | M: ≥34.6 cm; F: ≥31.9 cm | M: 31.0 – 34.5 cm; F: 28.3 – 31.8 cm | Incident LTCI certification | No LTCI certification vs. LTCI certification | HR: 1.16 | 0.61 – 2.18 | NR |
| Hoshi et al. 2012 | FRT | | X | M: ≥34.6 cm; F: ≥31.9 cm | M: 31.0 – 34.5 cm; F: 28.3 – 31.8 cm | Incident LTCI certification | No LTCI certification vs. LTCI certification or dead | HR: 1.21 | 0.68 – 2.13 | NR |
| Hoshi et al. 2012 | FRT | | X | M: ≥34.6 cm; F: ≥31.9 cm | M: 26.2 – 30.9 cm; F: 24.9 – 28.2 cm | Incident LTCI certification | No LTCI certification vs. LTCI certification | HR: 1.57 | 0.87 – 2.83 | NR |
| Hoshi et al. 2012 | FRT | | X | M: ≥34.6 cm; F: ≥31.9 cm | M: 26.2 – 30.9 cm; F: 24.9 – 28.2 cm | Incident LTCI certification | No LTCI certification vs. LTCI certification or dead | HR: 1.73 | 1.02 – 2.92 | NR |
| Hoshi et al. 2012 | FRT | | X | M: ≥34.6 cm; F: ≥31.9 cm | M: ≤26.1 cm; F: ≤24.8 cm | Incident LTCI certification | No LTCI certification vs. LTCI certification | HR: 2.45 | 1.39 – 4.34 | NR |
| Hoshi et al. 2012 | FRT | | X | M: ≥34.6 cm; F: ≥31.9 cm | M: ≤26.1 cm; F: ≤24.8 cm | Incident LTCI certification | No LTCI certification vs. LTCI certification or dead | HR: 2.46 | 1.47 – 4.13 | NR |
| Hoshi et al. 2012 | Gait speed (max) | | X | Fast: M: ≥2.08 m/s; F: ≥1.85 m/s | Slow: M: 1.88 – 2.07 m/s; F: 1.66 – 1.84 | Incident LTCI certification | No LTCI certification vs. LTCI certification | HR: 1.47 | 0.75 – 2.87 | NR |
| Hoshi et al. 2012 | Gait speed (max) | | X | Fast: M: ≥2.08 m/s; F: ≥1.85 m/s | Slow: M: 1.88 – 2.07 m/s; F: 1.66 – 1.84 | Incident LTCI certification | No LTCI certification vs. LTCI certification or dead | HR: 1.52 | 0.87 – 2.67 | NR |
| Hoshi et al. 2012 | Gait speed (max) | | X | Fast: M: ≥2.08 m/s; F: ≥1.85 m/s | Slow: M: 1.66 – 1.87 m/s; F: 1.48 – 1.65 | Incident LTCI certification | No LTCI certification vs. LTCI certification | HR: 2.25 | 1.20 – 4.21 | NR |
| Hoshi et al. 2012 | Gait speed (max) | | X | Fast: M: ≥2.08 m/s; F: ≥1.85 m/s | Slow: M: 1.66 – 1.87 m/s; F: 1.48 – 1.65 | Incident LTCI certification | No LTCI certification vs. LTCI certification or dead | HR: 1.88 | 1.10 – 3.23 | NR |
| Hoshi et al. 2012 | Gait speed (max) | | X | Fast: M: ≥2.08 m/s; F: ≥1.85 m/s | Slow: M: 0 – 1.65 m/s; F: 0 – 1.47 m/s | Incident LTCI certification | No LTCI certification vs. LTCI certification | HR: 3.66 | 2.00 – 6.69 | NR |
| Hoshi et al. 2012 | Gait speed (max) | | X | Fast: M: ≥2.08 m/s; F: ≥1.85 m/s | Slow: M: 0 – 1.65 m/s; F: 0 – 1.47 m/s | Incident LTCI certification | No LTCI certification vs. LTCI certification or dead | HR: 3.02 | 1.79 – 5.08 | NR |
| Hoshi et al. 2012 | TUG | | X | Fast: M: ≤7.73 sec; F: ≤8.09 sec | Slow: M: 7.74 – 8.64 sec; F: 8.10 – 9.13 sec | Incident LTCI certification | No LTCI certification vs. LTCI certification | HR: 1.31 | 0.68 – 2.52 | NR |
| Hoshi et al. 2012 | TUG | | X | Fast: M: ≤7.73 sec; F: ≤8.09 sec | Slow: M: 7.74 – 8.64 sec; F: 8.10 – 9.13 sec | Incident LTCI certification | No LTCI certification vs. LTCI certification or dead | HR: 1.41 | 0.80 – 2.48 | NR |
| Hoshi et al. 2012 | TUG | | X | Fast: M: ≤7.73 sec; F: ≤8.09 sec | M: 8.65 – 9.60 sec; F: 9.14 – 10.35 sec | Incident LTCI certification | No LTCI certification vs. LTCI certification | HR: 1.86 | 1.01 – 3.42 | NR |
| Hoshi et al. 2012 | TUG | | X | Fast: M: ≤7.73 sec; F: ≤8.09 sec | M: 8.65 – 9.60 sec; F: 9.14 – 10.35 sec | Incident LTCI certification | No LTCI certification vs. LTCI certification or dead | HR: 1.73 | 1.01 – 2.97 | NR |
| Hoshi et al. 2012 | TUG | | X | Fast: M: ≤7.73 sec; F: ≤8.09 sec | M: ≥9.61 sec; F: ≥10.36 sec | Incident LTCI certification | No LTCI certification vs. LTCI certification | HR: 2.94 | 1.63 – 5.31 | NR |
| Hoshi et al. 2012 | TUG | | X | Fast: M: ≤7.73 sec; F: ≤8.09 sec | M: ≥9.61 sec; F: ≥10.36 sec | Incident LTCI certification | No LTCI certification vs. LTCI certification or dead | HR: 2.65 | 1.58 – 4.46 | NR |

| Author(s)/report | Predictor (mobility assessment) | Continuous | Categorial | High mobility (reference) | Low mobility | Disability outcome | Disability categories | Association | 95% CI | p-value |
|---|---|---|---|---|---|---|---|---|---|---|
| Huang et al. 2010 | BBS | | X | ≥50 points | <50 points | Incident ADL | Independent vs. dependent | OR: 0.851 | 0.74 – 0.99 | NR |
| Huang et al. 2010 | BBS | | X | ≥50 points | <50 points | Incident ADL | Independent vs. dependent | AUC: 0.821 | 0.71 – 0.93 | NR |
| Huang et al. 2010 | Gait speed (usual) | | X | Fast: ≥0.67 m/s | Slow: <0.67 m/s | Incident ADL | Independent vs. dependent | OR: 0.286 | 0.01 – 8.79 | NR |
| Huang et al. 2010 | Gait speed (usual) | | X | Fast: ≥0.67 m/s | Slow: <0.67 m/s | Incident ADL | Independent vs. dependent | AUC: 0.771 | 0.66 – 0.88 | NR |
| Huang et al. 2010 | SPPB | | X | ≥7.6 points | <7.6 points | Incident ADL | Independent vs. dependent | OR: 0.798 | 0.57 – 1.11 | NR |
| Huang et al. 2010 | SPPB | | X | ≥7.6 points | <7.6 points | Incident ADL | Independent vs. dependent | AUC: 0.792 | 0.68 – 0.90 | NR |
| Huang et al. 2010 | TUG | | X | <12.5 sec | ≥12.5 sec | Incident ADL | Independent vs. dependent | OR: 1.142 | 0.97 – 1.35 | NR |
| Huang et al. 2010 | TUG | | X | <12.5 sec | ≥12.5 sec | Incident ADL | Independent vs. dependent | AUC: 0.792 | 0.68 – 0.91 | NR |
| Lee at al. 2020 | TUG | | X | <10 sec | ≥10 sec | Incident LTCI certification | No LTCI certification vs. LTCI certification | HR: 1.65 | 1.33 – 2.04 | NR |
| Makizako et al. 2015 | Gait speed (usual) | | X | ≥1.0 m/s | <1.0 m/s | Incident LTCI certification | No LTCI certification vs. LTCI certification | HR: 2.32 | 1.62 – 3.33 | <0.001 |
| Makizako et al. 2017 | CRT (x5) | | X | <10 sec | ≥10 sec | Incident care need by LTCI certification system (care level ≥4/5) | No certification or LTCI care level ≤3 vs LTCI care level ≥4 | HR: 1.88 | 1.11 – 3.20 | 0.0190 |
| Makizako et al. 2017 | CRT (x5) | | X | <10 sec | ≥10 sec | Incident care need by LTCI certification system (care level ≥4/5) | No certification or LTCI care level ≤3 vs LTCI care level ≥4 | AUC: 0.68 | 0.64 – 0.72 | <0.0001 |
| Makizako et al. 2017 | CRT (x5) | | X | <10 sec | ≥10 sec | Incident care need by LTCI certification system (care level ≥4/5) | No certification or LTCI care level ≤3 vs LTCI care level ≥4 | KMC | NA | NA |
| Makizako et al. 2017 | TUG | | X | <9 sec | ≥9 sec | Incident care need by LTCI certification system (care level ≥4/5) | No certification or LTCI care level ≤3 vs LTCI care level ≥4 | HR: 2.24 | 1.42 – 3.53 | 0.0005 |
| Makizako et al. 2017 | TUG | | X | <9 sec | ≥9 sec | Incident care need by LTCI certification system (care level ≥4/5) | No certification or LTCI care level ≤3 vs LTCI care level ≥4 | AUC: 0.74 | 0.70 – 0.78 | <0.0001 |
| Makizako et al. 2017 | TUG | | X | <9 sec | ≥9 sec | Incident care need by LTCI certification system (care level ≥4/5) | No certification or LTCI care level ≤3 vs LTCI care level ≥4 | KMC | NA | NA |
| Minneci et al. 2015 | 6-min | X | | per unit increase | NA | Incident ADL | Independent vs. dependent | OR: 0.993 | 0.988 – 0.997 | NR |
| Minneci et al. 2015 | 6-min | X | | continues | NA | Incident ADL | Independent vs. dependent | AUC: 0.74 | NR | NR |
| Minneci et al. 2015 | Gait speed (usual) | X | | continues | NA | Incident ADL | Independent vs. dependent | AUC: 0.73 | NR | NR |
| Minneci et al. 2015 | Gait speed (usual) | X | | per unit increase | NA | Incident ADL | Independent vs. dependent | OR: 0.08 | 0.02 – 0.36 | NR |
| Minneci et al. 2015 | SPPB | X | | per unit increase | NA | Incident ADL | Independent vs. dependent | OR: 0.74 | 0.61 – 0.89 | NR |
| Minneci et al. 2015 | SPPB | X | | continues | NA | Incident ADL | Independent vs. dependent | AUC: 0.71 | NR | NR |
| Montero-Odasso et al. 2005 | Gait speed (usual) | | X | High: ≥1.1 m/s | Intermediate: 0.7 – 1.0 m/s | Need/ Requirement for a caregiver | No caregiver vs. caregiver | RR: 1.64 | 0.7 – 4.1 | NR |
| Montero-Odasso et al. 2005 | Gait speed (usual) | | X | High: ≥1.1 m/s | Low: <0.7 m/s | Need/ Requirement for a caregiver | No caregiver vs. caregiver | RR: 9.5 | 1.3 – 72.5 | NR |
| Montero-Odasso et al. 2005 | Gait speed (usual) | | X | High: ≥1.1 m/s | Intermediate: 0.7 – 1.0 m/s; Low: <0.7 m/s | Nursing home placement | Community-dwelling vs. nursing home | NA (only 3 events) | NA | NA |
| Moriya et al. 2013 | one-leg standing time with eyes open (OLST) | | X | Female: good balance: 30.0 – 120.0 sec | Female: low balance: 14.0 – 29.9 sec | Incident LTCI certification | No LTCI certification vs. LTCI certification | HR: 3.4 | 1.1 – 10.4 | 0.030 |
| Moriya et al. 2013 | one-leg standing time with eyes open (OLST) | | X | Female: good balance: 30.0 – 120.0 sec | Female: low balance: 6.0 – 13.9 sec | Incident LTCI certification | No LTCI certification vs. LTCI certification | HR: 4.9 | 1.6 – 14.5 | 0.004 |
| Moriya et al. 2013 | one-leg standing time with eyes open (OLST) | | X | Female: good balance: 30.0 – 120.0 sec | Female: low balance: 0.0 – 5.9 sec | Incident LTCI certification | No LTCI certification vs. LTCI certification | HR: 8.1 | 2.9 – 23.0 | <0.001 |

| Author(s)/report | Predictor (mobility assessment) | Continuous | Categorial | High mobility (reference) | Low mobility | Disability outcome | Disability categories | Association | 95% CI | p-value |
|---|---|---|---|---|---|---|---|---|---|---|
| Moriya et al. 2013 | one-leg standing time with eyes open (OLST) | | X | Male: good balance: 40.0 – 120.0 sec | Male: low balance: 18.0 – 39.9 sec | Incident LTCI certification | No LTCI certification vs. LTCI certification | HR: 5.1 | 0.6 – 43.9 | 0.100 |
| Moriya et al. 2013 | one-leg standing time with eyes open (OLST) | | X | Male: good balance: 40.0 – 120.0 sec | Male: low balance: 6.0 – 17.9 | Incident LTCI certification | No LTCI certification vs. LTCI certification | HR: 7.2 | 0.9 – 58.8 | 0.060 |
| Moriya et al. 2013 | one-leg standing time with eyes open (OLST) | | X | Male: good balance: 40.0 – 120.0 sec | Male: low balance: 0.0 – 5.9 | Incident LTCI certification | No LTCI certification vs. LTCI certification | HR: 19.7 | 2.6 – 148.3 | 0.004 |
| Onder et al. 2005 | Balance Test (SPPB) | X | | per 10.2 s increase | NA | Incident ADL disability (progressive) | Disabled vs. non-disabled | RR: 0.81 | 0.66 – 0.89 | <0.05 |
| Onder et al. 2005 | Balance Test (SPPB) | X | | per 10.2 s increase | NA | Incident ADL disability (catastrophic) | Disabled vs. non-disabled | RR: 0.93 | 0.67 – 1.29 | ns (>0.05) |
| Onder et al. 2005 | Balance Test (SPPB) | X | | per 10.2 s increase | NA | Incident ADL disability (progressive) | Disabled vs. non-disabled | AUC: 0.63 | NR | <0.05 |
| Onder et al. 2005 | Balance Test (SPPB) | X | | per 10.2 s increase | NA | Incident ADL disability (catastrophic) | Disabled vs. non-disabled | AUC: 0.58 | NR | ns (>0.05) |
| Onder et al. 2005 | CRT (x5) | X | | per 8.4 s increase | NA | Incident ADL disability (progressive) | Disabled vs. non-disabled | RR: 1.54 | 1.29 – 1.83 | <0.05 |
| Onder et al. 2005 | CRT (x5) | X | | per 8.4 s increase | NA | Incident ADL disability (catastrophic) | Disabled vs. non-disabled | RR: 1.14 | 0.51 – 1.58 | ns (>0.05) |
| Onder et al. 2005 | CRT (x5) | X | | per 8.4 s increase | NA | Incident ADL disability (progressive) | Disabled vs. non-disabled | AUC: 0.67 | NR | <0.05 |
| Onder et al. 2005 | CRT (x5) | X | | per 8.4 s increase | NA | Incident ADL disability (catastrophic) | Disabled vs. non-disabled | AUC: 0.54 | NR | ns (>0.05) |
| Onder et al. 2005 | Gait speed (usual) | X | | per 0.31 m/s increase | NA | Incident ADL disability (progressive) | Disabled vs. non-disabled | RR: 0.65 | 0.52 – 0.82 | <0.05 |
| Onder et al. 2005 | Gait speed (usual) | X | | per 0.31 m/s increase | NA | Incident ADL disability (catastrophic) | Disabled vs. non-disabled | RR: 0.72 | 0.53 – 0.99 | <0.05 |
| Onder et al. 2005 | Gait speed (usual) | X | | per 0.31 m/s increase | NA | Incident ADL disability (progressive) | Disabled vs. non-disabled | AUC: 0.66 | NR | <0.05 |
| Onder et al. 2005 | Gait speed (usual) | X | | per 0.31 m/s increase | NA | Incident ADL disability (catastrophic) | Disabled vs. non-disabled | AUC: 0.60 | NR | <0.05 |
| Ostir et al. 1998 | CRT (x5) | | X | ≤ 10.9 sec | 11.0 – 13.5 sec | Incident ADL | Independent vs. dependent | OR: 1.0 | 0.4 – 2.6 | NR |
| Ostir et al. 1998 | CRT (x5) | | X | ≤ 10.9 sec | 13.6 – 16.4 sec | Incident ADL | Independent vs. dependent | OR: 1.6 | 0.7 – 3.9 | NR |
| Ostir et al. 1998 | CRT (x5) | | X | ≤ 10.9 sec | ≥16.5 sec | Incident ADL | Independent vs. dependent | OR: 2.8 | 1.2 – 6.4) | NR |
| Ostir et al. 1998 | Gait speed (usual) | | X | ≤3 sec | 4 – 5 sec | Incident ADL | Independent vs. dependent | OR: 3.6 | 0.8 – 15.7 | NR |
| Ostir et al. 1998 | Gait speed (usual) | | X | ≤3 sec | 6 – 8 sec | Incident ADL | Independent vs. dependent | OR: 4.3 | 1.0 – 19.0 | NR |
| Ostir et al. 1998 | Gait speed (usual) | | X | ≤3 sec | ≥9 sec | Incident ADL | Independent vs. dependent | OR: 5.4 | 1.2 – 23.6 | NR |
| Ostir et al. 1998 | Lower body function score (mSPPB) | | X | 9 – 11 points | 1 – 4 points | Incident ADL | Independent vs. dependent | OR: 6.2 | 2.4 – 16.0 | NR |
| Ostir et al. 1998 | Lower body function score (mSPPB) | | X | 9 – 11 points | 5 – 8 points | Incident ADL | Independent vs. dependent | OR: 2.0 | 0.9 – 4.2 | NR |
| Ostir et al. 1998 | Standing balance test | | X | 3 points | 1 point | Incident ADL | Independent vs. dependent | OR: 2.4 | 1.0 – 5.4 | NR |
| Ostir et al. 1998 | Standing balance test | | X | 3 points | 2 points | Incident ADL | Independent vs. dependent | OR: 2.5 | (1.2 – 4.8) | NR |
| Rosenberg et al. 2019 | Gait speed (usual) | X | | per 0.1 m/s increment | NA | Nursing home transfer | Community-dwelling vs. nursing home | HR: 0.13 | 0.03 – 0.57 | 0.01 |
| Sakamoto et al. 2016 | TUG | | X | <15 sec | ≥15 sec | Incident ADL | Independent vs. dependent | OR: 2.74 | 1.14 – 6.58 | 0.015 |
| Sakamoto et al. 2016 | FRT | | X | ≥20 cm | <20 cm | Incident ADL | Independent vs. dependent | OR: 2.92 | 1.09 – 7.83 | 0.033 |
| Shinkai et al. 2003 | Gait speed (fast) | X | | decrease by a quartile | NA | Incident ADL | Independent vs. dependent | HR: 1.40 | 1.22 – 1.61 | NR |
| Shinkai et al. 2003 | Gait speed (usual) | X | | decrease by a quartile | NA | Incident ADL | Independent vs. dependent | HR: 1.31 | 1.14 – 1.50 | NR |

| Author(s)/report | Predictor (mobility assessment) | Continuous | Categorial | High mobility (reference) | Low mobility | Disability outcome | Disability categories | Association | 95% CI | p-value |
|---|---|---|---|---|---|---|---|---|---|---|
| Shinkai et al. 2003 | One-leg standing time | X | | decrease by a quartile | NA | Incident ADL | Independent vs. dependent | HR: 1.41 | 1.22 – 1.62 | NR |
| Stenholm et al. 2014 | Gait speed (usual) | X | | Per 0.1 m/s increment | NA | Incident ADL (3-year follow-up) | Independent vs. dependent | OR: 0.63 | 0.52–0.76 | NR |
| Stenholm et al. 2014 | Gait speed (usual) | X | | Per 0.1 m/s increment | NA | Incident ADL (6-year follow-up) | Independent vs. dependent | OR: 0.69 | 0.59–0.81 | NR |
| Stenholm et al. 2014 | SPPB | X | | Per 1-point increment | NA | Incident ADL (3-year follow-up) | Independent vs. dependent | OR: 0.57 | 0.48–0.68 | NR |
| Stenholm et al. 2014 | SPPB | X | | Per 1-point increment | NA | Incident ADL (6-year follow-up) | Independent vs. dependent | OR: 0.63 | 0.54–0.73 | NR |
| Studenski et al. 2003 | Gait speed (usual) | X | | Unit: 0.2 m/sec | NA | Incident difficulty in personal care | No difficulty in personal care vs. difficulty in personal care | OR: 0.62 | NR | 0.0003 |
| Studenski et al. 2003 | LEPB | X | | per unit increase | NA | Incident difficulty in personal care | No difficulty in personal care vs. difficulty in personal care | NR | NR | NR |
| Tsutsumimoto et al. 2016 | Gait speed (usual) | | X | ≥1.0 m/s | <1.0 m/s | Incident LTCI certification | No LTCI certification vs. LTCI certification | HR: 2.44 | 1.71 – 3.47 | <0.001 |
| Vaarst et al. 2021 | SPPB | | X | ≥10 points | ≤9 points | Incident ADL (personal care) | Independent vs. dependent | HR: 1.90 | 1.29 – 2.81 | <0.01 |
| Verghese et a. 2012 | Gait speed (usual) | X | | Per 10 cm/sec change (decrease) | NA | Incident ADL | Independent vs. dependent | HR: 1.15 | 1.01 – 1.32 | 0.040 |
| Verghese et a. 2012 | SPPB | X | | Per 1-point chane (decrease) | NA | Incident ADL | Independent vs. dependent | HR: 1.23 | 1.08 – 1.40 | 0.002 |
| Verghese et a. 2012 | WTTT | X | | Per 10 cm/sec change (decrease) | NA | Incident ADL | Independent vs. dependent | HR: 1.13 | 1.03 – 1.23 | 0.008 |
| Verghese et a. 2012 | WTTT | | X | >70 cm/s | ≤70 cm/s | Incident ADL | Independent vs. dependent | HR: 1.82 | 1.15 – 2.86 | 0.010 |
| Verghese et a. 2012 | WTTT | | X | >60 cm/s | ≤60 cm/s | Incident ADL | Independent vs. dependent | HR: 1.35 | 0.88 – 2.01 | 0.172 |
| Verghese et a. 2012 | WTTT | | X | >50 cm/s | ≤50 cm/s | Incident ADL | Independent vs. dependent | HR: 1.51 | 0.96 – 2.37 | 0.072 |
| Woo et al. 1999 | Gait speed (usual) | NR | NR | NR | NR | Dependency (Barthel Index) | Barthel Index score 20 of 20 (max) vs. Barthel Index score <20 | M: OR: 1.190<br>F: OR: 1.163 | M: 1.13 – 1.26<br>F: 1.12 – 1.21 | M+F: <0.001 |
| Woo et al. 1999 | Gait speed (usual) | NR | NR | NR | NR | Institutionalization | Community-dwelling vs. institutionalized | M: OR: 1.087<br>F: OR: 1.031 | M: 0.99 – 1.19<br>F: 1.00 – 1.06 | M: 0.066<br>F: 0.038 |
| Woo et al. 1999 | Stride lengths | NR | NR | NR | NR | Dependency (Barthel Index) | Barthel Index score 20 of 20 (max) vs. Barthel Index score <20 | M: OR: 0.082<br>F: OR: 0.021 | M: 0.04 – 0.18<br>F: 0.01 – 0.05 | M+F:<0.001 |
| Woo et al. 1999 | Stride lengths | NR | NR | NR | NR | Institutionalization | Community-dwelling vs. institutionalized | M: OR: 0.094<br>F: OR: 0.096 | M: 0.01 – 0.62<br>F: 0.02 – 0.42) | M: 0.014<br>F: 0.002 |
| Zhang et al. 2013 | CRT (x5) | | X | <11.2 s (group 1) | 11.2 – 13.6 s (group 2) | Incident ADL | Independent vs. dependent | OR: 0.88 | 0.22 – 3.46 | 0.86 |
| Zhang et al. 2013 | CRT (x5) | | X | <11.2 s (group 1) | 13.7 – 16.6 s (group 3) | Incident ADL | Independent vs. dependent | OR: 0.50 | 0.05 – 4.89 | 0.55 |
| Zhang et al. 2013 | CRT (x5) | | X | <11.2 s (group 1) | >16.6 s (group 4) | Incident ADL | Independent vs. dependent | OR: 0.85 | 0.09 – 8.52 | 0.89 |
| Zhang et al. 2013 | CRT (x5) | | X | <11.2 s (group 1) | inability to complete (group 5) | Incident ADL | Independent vs. dependent | OR: 24.70 | 2.83 – 215.44 | <0.01 |

**Abbreviations:**

6-min = 6-minute walk test; ADL = activities of daily living; AUC = area under the curve; BBS = Berg Balance Scale; BI = Barthel Index; CI = confidence interval; CRT = Chair Rise Test; F = female; FRT = Functional Reach Test; HR = Hazard Ratio; KMC = Kaplan-Meier curves; LLFDI = Late Life Function and Disability Instrument; LTCI = Long term care insurance; M = male; mSPPB = modified Short Physical Performance Battery; NA = not applicable; NR = not reported; NS = not significant; OLST = one-leg standing time with eyes open; OR = Odds Ratio; POMA = Performance-Oriented Mobility Assessment; RR = Relative Risk; Sen = Sensitivity; Spe = Specificity; SPPB = Short Physical Performance Battery; TUG = Timed Up and Go test; WTTT = Walking While Talking test

### eFigure 1: Flow chart

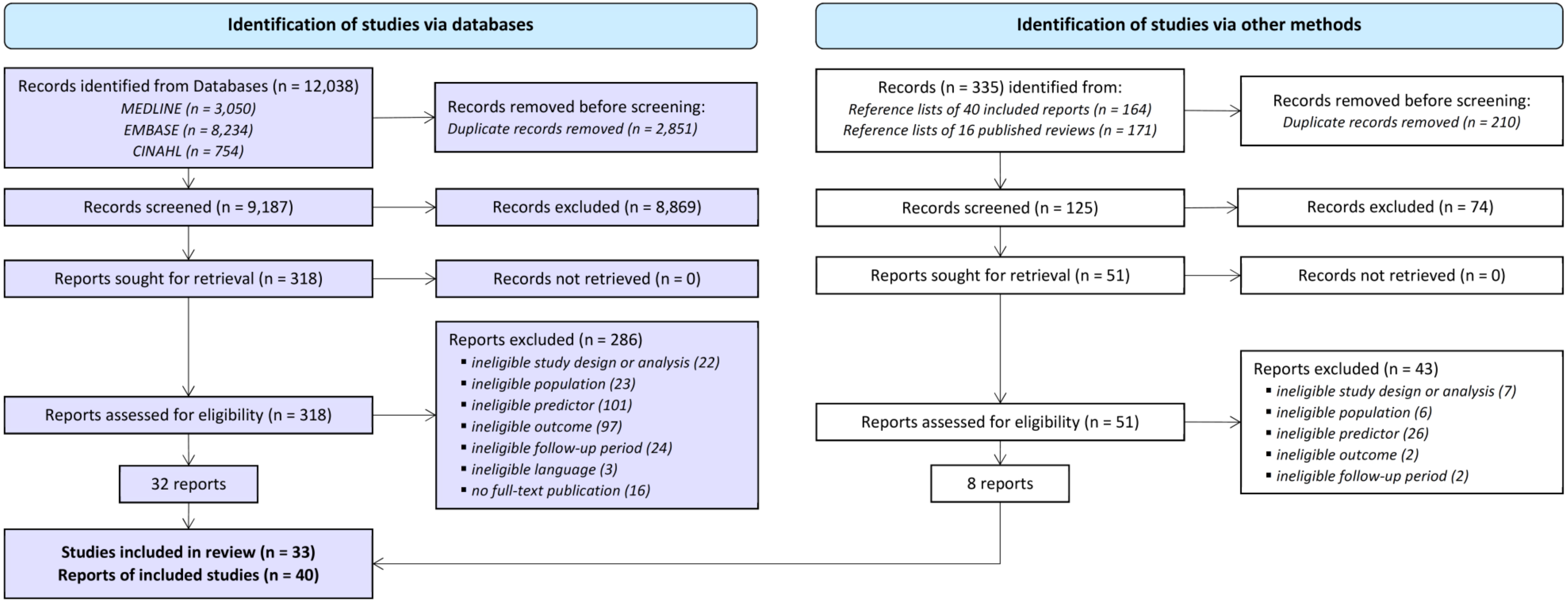

**eFigure 2: Risk of bias summary**

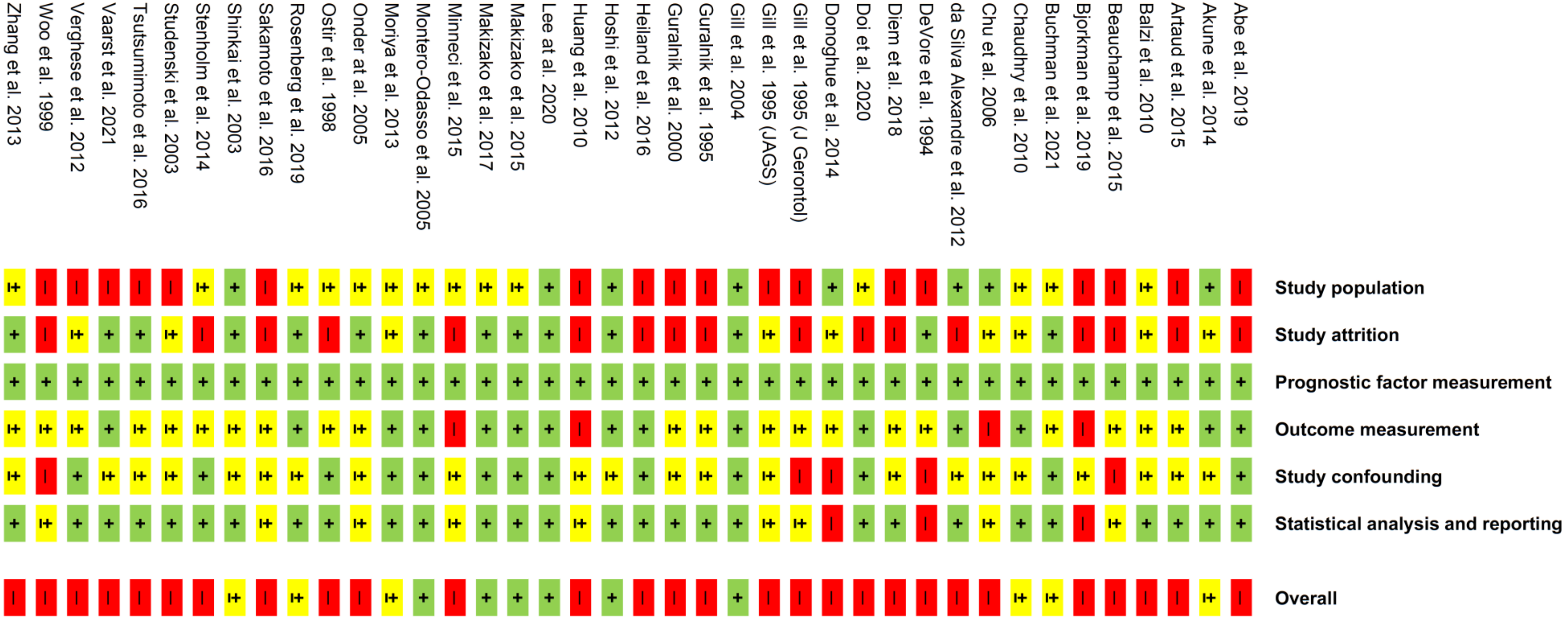

**eFigure 3: Risk of bias graph**

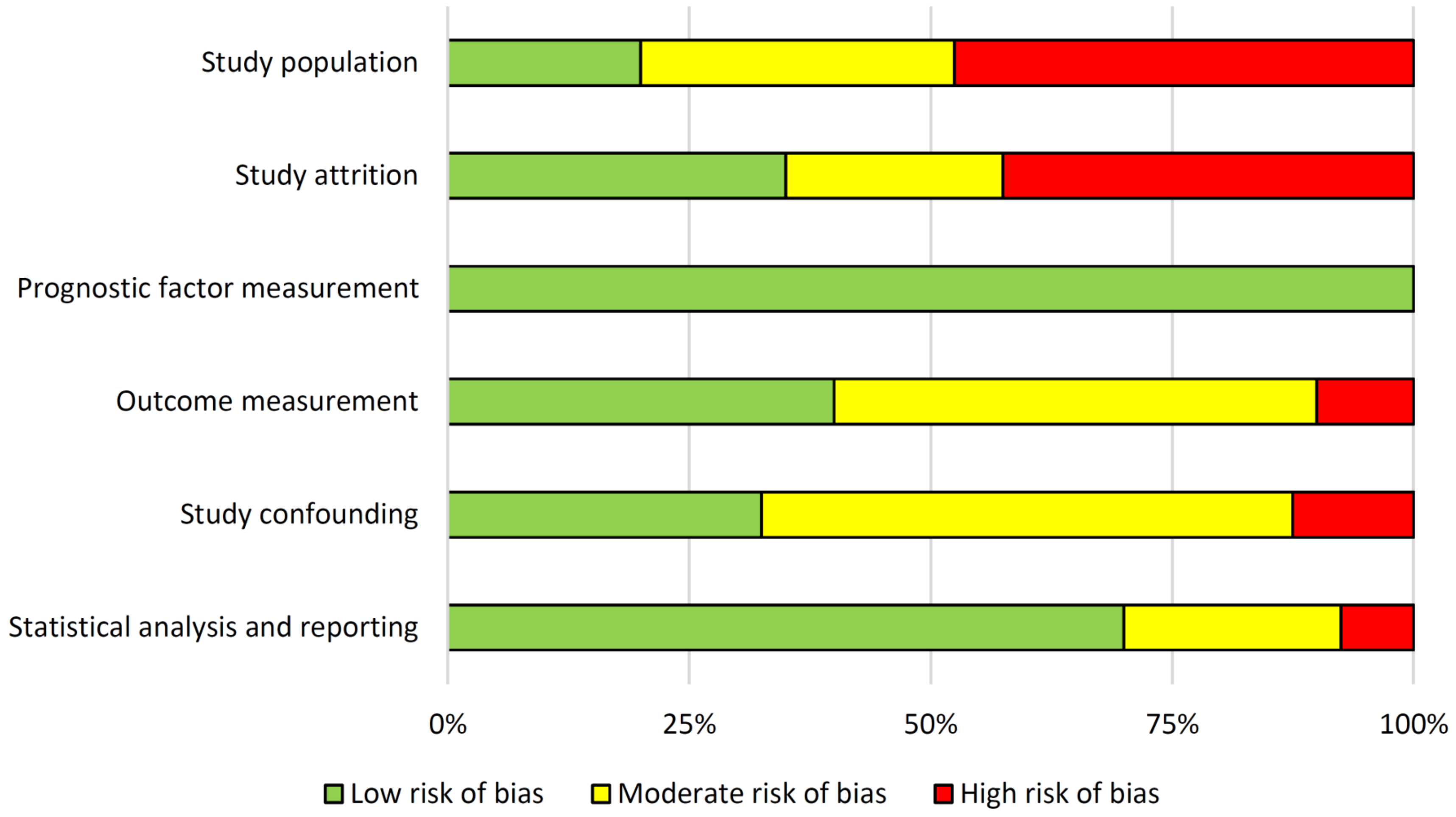

**eFigure 4: Funnel plot for reports on usual gait speed**

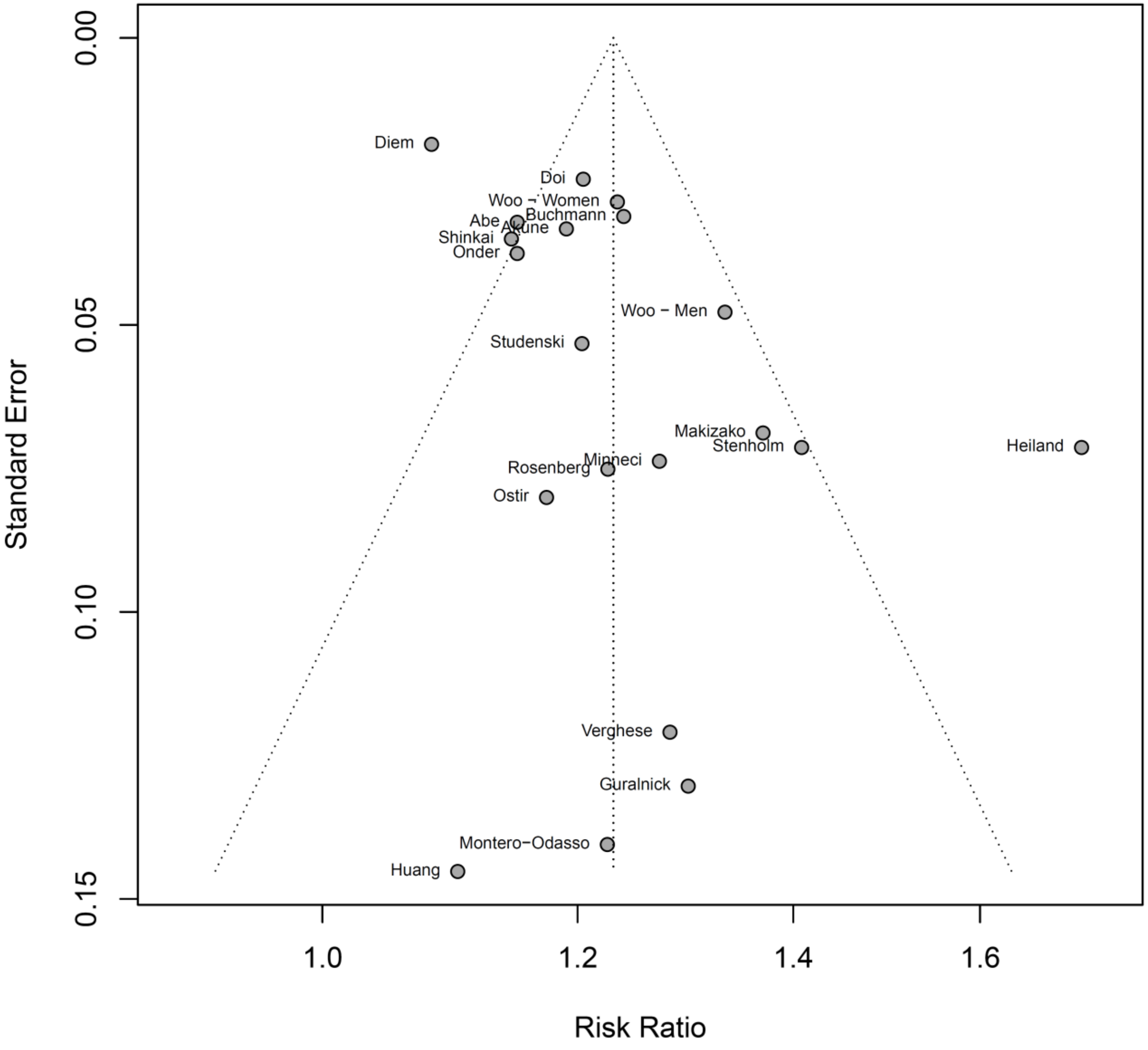

**eFigure 5: Funnel plot for reports on the Short Physical Performance Battery (SPPB)**

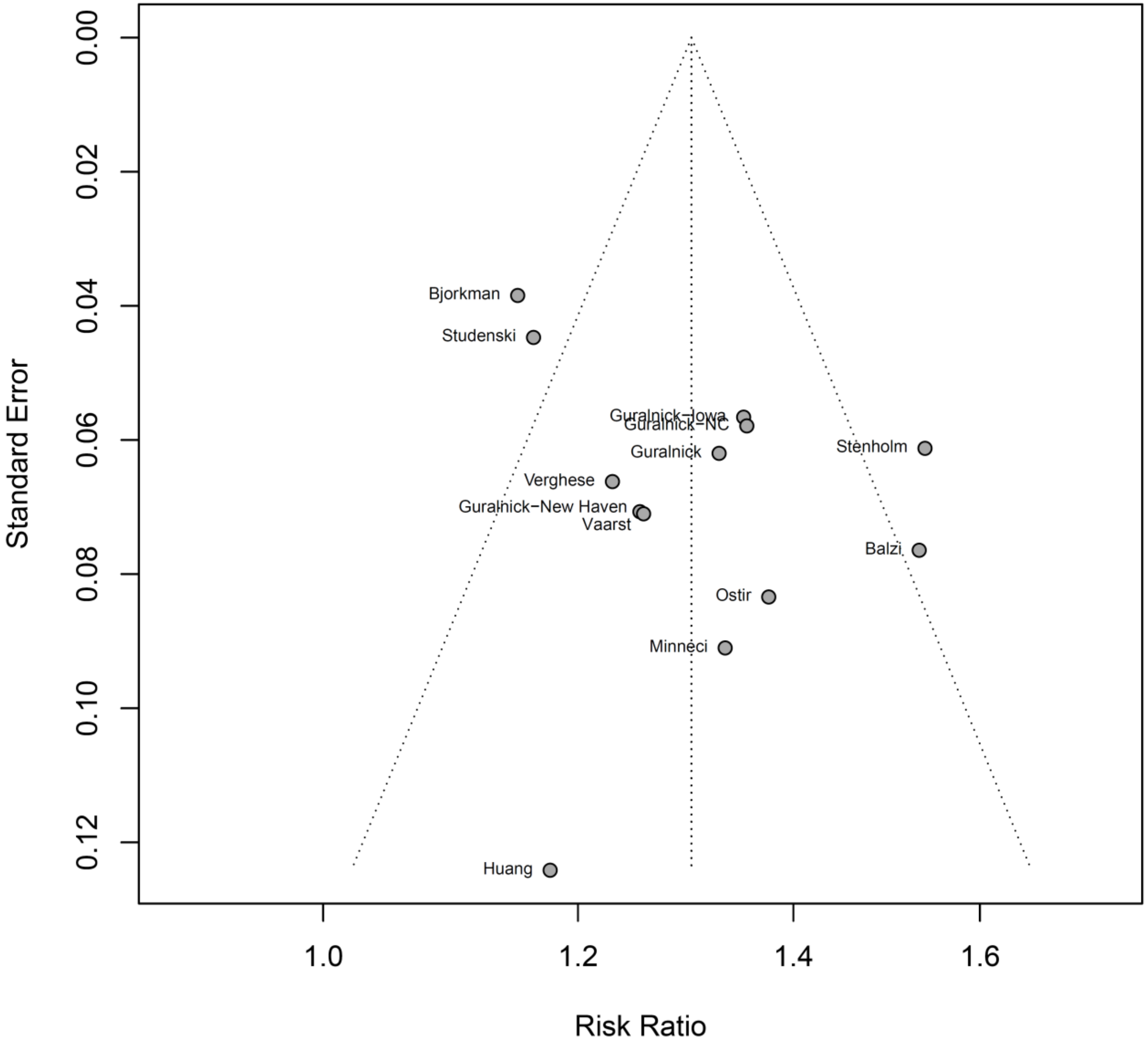

### eText 1: Eligibility criteria

Reports were eligible for inclusion if they met the following criteria:

(1) Design: Published reports of longitudinal cohort studies that estimated a direct association between baseline mobility capacity and subsequent development of disability over a follow-up period of 1 to 6 years.
(2) Population: The complete sample of study participants consisted of community-dwelling older adults; the mean age of study participants was ≥ 60 years; study participants were non-disabled at baseline. Reports were also included if study participants were living with only minimal (initial) impairment, with minimal functional inability, in a pre-frail condition, with support in IADLs or other similar conditions, as long as the participants were living independently.
(3) Prognostic factor: Studies that assessed at least one aspect of mobility capacity at baseline (according to the WHO definition of mobility)(World Health Organization 2001) with a standardized outcome assessment of mobility capacity that fulfilled the following criteria: performance-based or observer-based; feasible in an individual's home environment or general practitioner's facilities; feasible with older, potentially frail and cognitively impaired individuals; applicable within 10 minutes without special equipment; cost per measure are achievable (< 5 USD/EURO).
(4) Outcome: Studies that included at least one measure of ADL disability at follow-up. Outcomes such as 'nursing home admission' or 'care dependency' were considered as indicators of ADL disability.
(5) Association: Studies that reported at least one effect estimate of the association between baseline mobility capacity and the longitudinal development of disability, or on the predictive value of mobility capacity measures for the development of disability, such as C-statistics (sensitivity, specificity, AUC), odds ratios (OR), relative risk (RR), hazard ratio (HR) or provide data that allow for calculation of such estimates.

We excluded the following reports:

(1) Reports of studies with a cross-sectional design, study protocols, incomplete/ongoing studies, reviews/posters/commentary articles, reports with no available full-text version, and reports that reported on pooled analyses based on ≥ 2 samples from different cohort studies.
(2) Reports not written in English or German.
(3) Reports solely focusing on individuals with clinical disorders (such as Parkinson's disease, stroke, hip fracture, dementia) or individuals with any other disease-specific impairments.
(4) Reports including individuals from institutionalized setting (such as hospital inpatient clinics, aged care residencies, or nursing home environments).
(5) Reports of studies that included only prognostic factors irrelevant for this review (such as self-reported mobility capacity, unfeasible assessments, or change scores from mobility assessments).
(6) Reports of studies that included only mobility disability or IADL disability or other (disability) outcomes irrelevant for this review, because this does not reflect the concept of ADL disability.

### eText 2: Data extraction

A standardized data extraction sheet (based on the Cochrane template),[24] was developed, pilot-tested on five reports, and refined accordingly. Information was extracted by one reviewer and checked for accuracy by a second reviewer. Disagreements were resolved by discussion. We contacted the authors of seven reports up to three times over a month, of which two provided relevant data.

The following data were extracted from included reports: authors; country of origin; year of publication; study methodology, including study design, setting, inclusion and exclusion criteria, statistics; recruitment and study completion rates; population description and baseline characteristics (including age, sex, cognition); follow-up period; type of mobility capacity measure (predictor); type of disability measure (outcome); adjustment for any covariates; outcome data (eg, estimates of the association between mobility and disability); and references to other relevant studies. If possible, we calculated and reported the pooled mean values for the total baseline sample (eg, age, MMSE score) if study authors reported only the mean values for subgroups (eg, by gender). If not described elsewhere, effect sizes were extracted from text, tables, or figures.

Any measure of incident ADL disability (outcome domain) was eligible for inclusion, including nursing home admission, use of home care services or care dependency, assessed within 1 to 6 years after baseline. Data were based on the measurement method, unit, and cut-offs applied in analyses. If multiple follow-ups were reported, we extracted data on the longest follow-up. If results from multiple multivariable models were presented, we used the most fully adjusted model.

### eText 3: Additional information on the use of the Newcastle-Ottawa scale (NOS) in this review

Quality assessment was performed based on the following NOS categories: selection, comparability, and outcome. The NOS requires review authors to specify some items.

**Selection:**

For the area “Selection”, we defined the “average” population as “older adults free of disability living in the community”.

**Comparability:**

For the area “Comparability”, we defined that a positive rating was given if the study controlled for sex (a: most important factor) and if the study controls for least one of the following relevant factors: age, height, BMI, muscle mass, grip strength, nutritional status, depressive symptoms, cognition, comorbidities (a: additional factors).

For reports which reported associations clustered by gender, and for reports which included only male or female participants (e.g. only women), a positive rating was given for “study controlled for sex”.

**Outcome:**

For the area “Outcome”, we gave a positive rating if the report included (a) a complete follow up or (b) if subjects lost to follow up were ≤ 25%, or if a description provided of those lost was given. We gave a negative rating if (c) the follow up rate was > 25% and no description of those lost was given or (d) if no statement on follow-up rate was made by the study authors.
